# Genome and epigenome wide studies of plasma protein biomarkers for Alzheimer’s disease implicate TBCA and TREM2 in disease risk

**DOI:** 10.1101/2021.06.07.21258457

**Authors:** Robert F. Hillary, Danni A. Gadd, Daniel L. McCartney, Liu Shi, Archie Campbell, Rosie M. Walker, Craig W. Ritchie, Ian J. Deary, Kathryn L. Evans, Alejo J. Nevado-Holgado, Caroline Hayward, David J. Porteous, Andrew M. McIntosh, Simon Lovestone, Matthew R. Robinson, Riccardo E. Marioni

## Abstract

The levels of many blood proteins are associated with Alzheimer’s disease or its pathological hallmarks. Elucidating the molecular factors that control circulating levels of these proteins may help to identify proteins causally associated with the disease. Here, genome-wide and epigenome-wide studies (n_individuals_≤1,064) were performed on plasma levels of 281 Alzheimer’s disease-associated proteins, identified by a systematic review of the literature. We quantified the contributions of genetic and epigenetic variation towards inter-individual variability in plasma protein levels. Sixty-one independent genetic and 32 epigenetic loci were associated with expression levels of 49 proteins; eight and 24 of these respective findings are previously unreported. Novel findings included an association between plasma TREM2 levels and a polymorphism and CpG site within the *MS4A4A* locus. Through Mendelian randomisation analyses, causal associations were observed between higher plasma TBCA and TREM2 levels and lower Alzheimer’s disease risk. Our data inform the regulation of biomarker levels and their relationships with Alzheimer’s disease.

## Background

Alzheimer’s disease (AD) is one of the leading causes of disease burden and death globally (1, 2). Blood-based methods for assessing disease risk are potentially more cost-effective and less-invasive than neuroimaging methods or lumbar punctures for collecting cerebrospinal fluid (CSF). Further, blood-based measures that reflect brain pathology, including amyloid-beta and phosphorylated tau, are highly promising markers for diagnosis, and for recruitment and patient stratification in preventative trials (3-6). Approaches that use genomics and untargeted proteomics have suggested that there are signals in blood that might supplement targeted assays, and contribute to the understanding and prediction of AD (7-9). However, the relevance of many candidate protein markers identified by untargeted approaches to AD remains unclear (7, 10-12). Understanding the molecular factors that regulate the levels of AD-associated proteins may identify proteins with likely causal roles in disease risk (13-16).

Unlike genetic factors which remain largely stable over the life-course, differential DNA methylation (DNAm) profiles at individual CpG sites are influenced by genetic and non-genetic factors. These include dietary and lifestyle behaviours (17). DNAm data may capture independent information beyond genetic factors in explaining inter-individual variation in circulating protein levels. Several genome-wide association studies (GWAS) have catalogued polymorphisms associated with plasma protein levels and identified causal relationships between proteins and disease states including AD (13, 15, 18-24). Further, Zaghlool *et al*. (2020) performed the only large-scale epigenome-wide association study (EWAS) to date of plasma protein levels (>1,000 proteins) (25). Few studies have integrated GWAS and EWAS data to quantify the independent and combined contributions of genetic and epigenetic factors towards differential protein biomarker levels (26-28).

We performed a systematic review of studies that report associations between plasma proteins and AD diagnosis or related traits such as amyloid burden and cortical atrophy (29-40). We focused on studies that measured plasma protein levels using the SOMAscan® affinity proteomics platform (Somalogic® Inc.) as this matches the protocol used in our study, Generation Scotland. We identified 281 proteins that were also measured in our sample. Our first aim was to quantify the degree to which genome-wide genetic and DNA methylation factors explain inter-individual differences in plasma levels of 281 AD-associated proteins. Using these data, our second aim was to investigate whether plasma proteins have likely causal relationships with AD.

For our first aim, we performed a combined GWAS/EWAS on circulating levels of 281 proteins in up to 1,064 participants of the family-based Generation Scotland study. Using Bayesian penalised regression (through the BayesR+ software), we estimated the proportion of inter-individual variability in plasma protein levels that can be accounted for by variation in genetic and DNA methylation factors. BayesR+ implicitly adjusts for probe intercorrelations and data structure, including relatedness (41). Results were then integrated with publicly-available methylation and expression quantitative trait loci (mQTL/eQTL) data to probe the molecular mechanisms that might regulate protein abundances in plasma. For our second aim, Mendelian randomisation (MR) and colocalisation analyses tested for possibly causal relationships between plasma protein levels and AD phenotypes.

## Results

### Systematic identification of plasma proteins associated with Alzheimer’s disease

Following a systematic search of MEDLINE (Ovid), Embase (Ovid), Web of Science (Thomson Reuters) and preprint servers, twelve studies were identified that reported associations between SOMAscan® plasma proteins and AD or related traits (Figure 1, see Methods). In total, 359 unique proteins were identified and 22 (6.1%) were reported in more than one study (Additional File 1: Table S1-S3). In the Generation Scotland dataset, there were 309 SOMAmers (**S**low **O**ff-rate **M**odified **A**pta**mers**) that targeted 281/359 proteins of interest (Additional File 1: Table S4 and Additional File 2: Figure S1). The 281 unique proteins were brought forward for analyses (UniProt IDs and Seq-ids are shown in Additional 1: Table S5).

**Figure 1.**
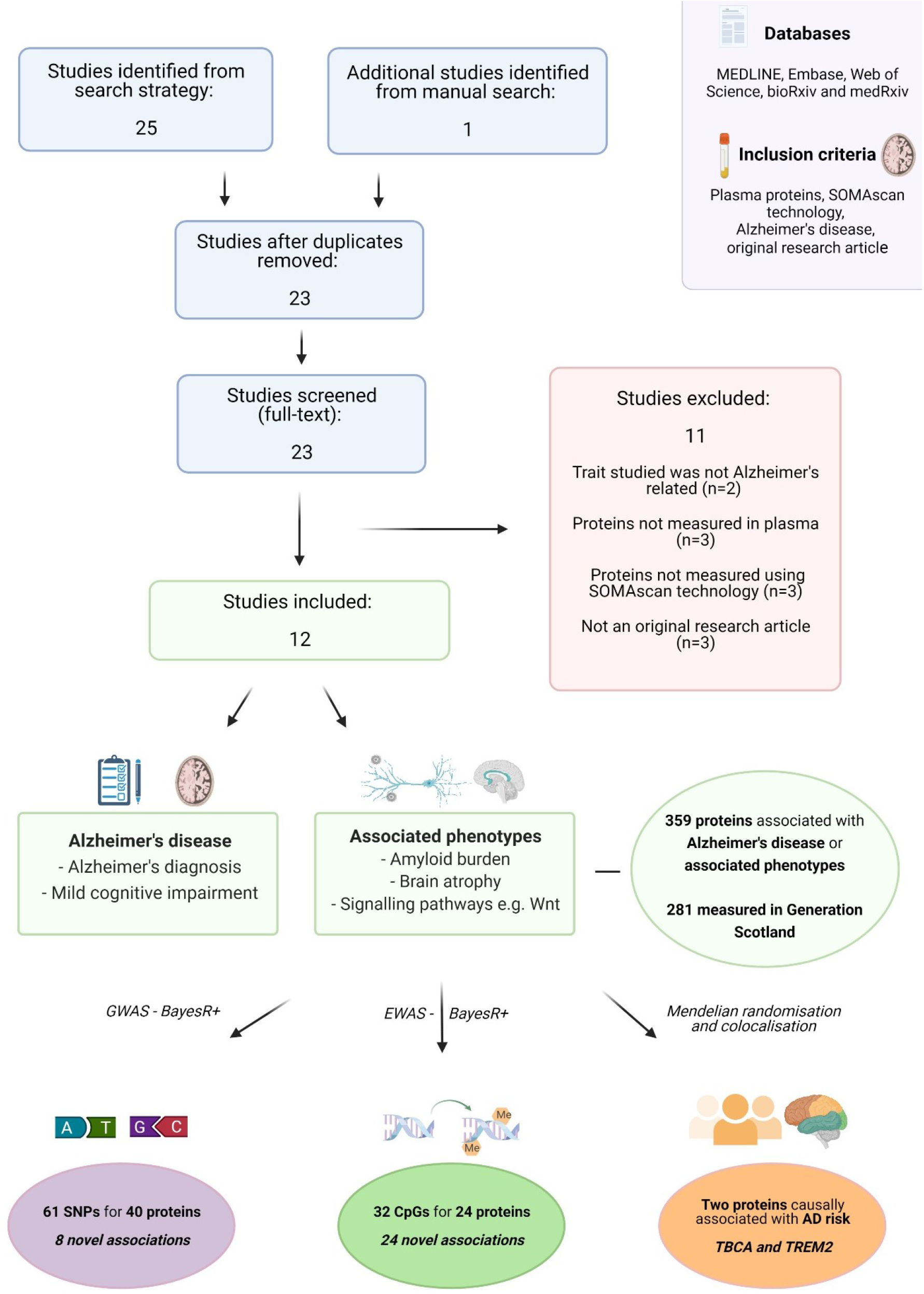
Systematic review of SOMAscan® plasma proteins that were associated with Alzheimer’s disease in the literature, and assessment of their molecular architectures and relationships with Alzheimer’s disease in the present study. The MEDLINE, Embase, Web of Science databases and preprint servers were queried to identify studies that reported associations between SOMAscan®-measured plasma proteins and Alzheimer’s disease. GWAS, EWAS and causal inference analyses were performed to identify molecular correlates of 281 AD-associated plasma protein levels and to probe their causal relationships with Alzheimer’s disease and related traits. AD, Alzheimer’s disease; EWAS, epigenome-wide association study; GWAS, genome-wide association study. Image created using Biorender.com.

### Genome-wide studies of plasma protein correlates of Alzheimer’s disease

There were 1,064 individuals with genotype and proteomic data in Generation Scotland. The mean age of the sample was 59.9 (SD=5.9) years and 59.1% of the sample were female. In the BayesR+ GWAS, 61 independent variants (or protein quantitative trait Loci, pQTLs) were associated with 42 SOMAmers that mapped to 40 unique protein targets (posterior inclusion probability (PIP) ≥95%; Additional File 1: Table S6). The phenotypic correlation structure of these 42 SOMAmers is presented in Additional File 2: Figure S2. The median correlation coefficient between SOMAmer levels was 0.17. Thirty-four pQTLs represented *cis* associations (pQTLs within 10 Mb of transcription start site (TSS) for a given gene) and 27 pQTLs were *trans-*chromosomal effects (Figure 2). The majority of variants were located in intronic regions using annotations from the ENSEMBL variant effect predictor (46.4%, Additional File 2: Figure S3) (42).

**Figure 2.**
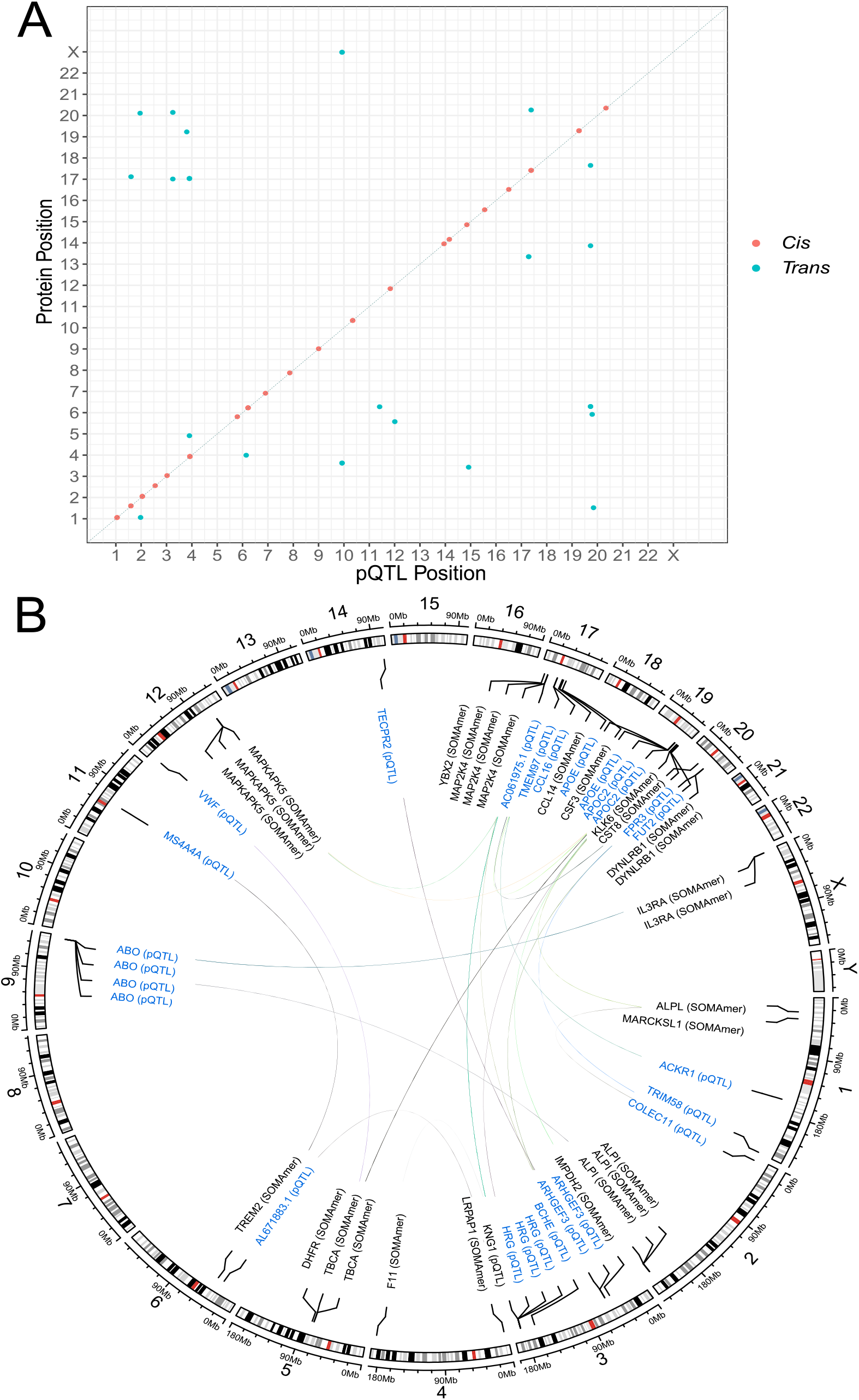
Genome-wide association studies of plasma protein levels previously associated with Alzheimer’s disease and disease-related phenotypes. (A) Chromosomal locations of pQTLs identified through Bayesian penalised regression GWAS. The *x*-axis shows the chromosomal location of pQTLs associated with the levels of SOMAmers that correlate with Alzheimer’s disease status or related pathways. The *y*-axis represents the position of the gene encoding the target protein. *Cis* (red circles); *trans* (blue circles). (B) A circos plot for the 27 *trans*-associated pQTLs from (A). Lines indicate an association between a pQTL and SOMAmer. GWAS, genome-wide association study; pQTL, protein quantitative trait locus.

Fifty-three pQTLs were previously reported in GWAS of blood protein levels (Additional File 1: Table S7) (13-15, 18-20, 22, 23, 27, 28, 43-50). Variants either directly replicated known associations or showed high linkage disequilibrium (LD, *r*^2^>0.75) with known pQTLs for queried proteins. Relative effect sizes reported in the literature correlated strongly with those in our study (*r*=0.78, 95% confidence interval (CI) = [0.68, 0.85]). We identified eight novel pQTLs associated with eight unique proteins. Three pQTLs were in *cis* (GM2A, MATN3 and IL1RAP). Five pQTLs represented *trans-*chromosomal effects: rs1126680 (*BCHE* for KLK6), rs1354034 (*ARHGEF3* for YBX2), rs17695224 (*FPR3* for MARCKSL1), rs3820897 (*COLEC11* for ALPL) and rs1582763 (*MS4A4A* for TREM2).

Thirty-one pQTLs were associated with at least one trait in the GWAS Catalog at P<5×10^−8^ (range=1-78 associations, Additional File 1: Table S8) (51). In relation to AD traits, the *trans* pQTL in *MS4A4A* (rs1582763) for TREM2 levels was associated with AD in *APOE* ε4 carriers and family history of AD (8, 52). Further, the *trans* pQTL in *APOE* (rs769449) for TBCA levels was associated with 15 AD-related traits including genetic predisposition to AD and CSF biomarkers of the disease (53, 54).

BayesR+ was used to estimate the proportions of inter-individual variation in individual plasma protein levels that were attributable to common SNPs (minor allele frequency>1%). Estimates ranged from 5.4% (PRL; 95% credible interval (CrI) = [0%, 24.9%]) to 73.0% (IL1RAP; 95% CrI = [56.1%, 82.7%]), with a median estimate of 13.0% across all 309 SOMAmers (Additional File 1: Table S9).

### Colocalisation of protein QTLs with expression QTLs

The 34 *cis* pQTLs identified in BayesR+ were annotated to 23 unique proteins. For 11/23 proteins, at least one pQTL was previously reported to be an expression QTL for the respective gene in blood tissue (eQTL consortium database) (55). The R package *coloc* (56) was used to test the hypothesis that one causal variant might underpin differences in gene expression (eQTL) and protein levels (pQTL) for each gene of interest. For two proteins (PCSK7 and GM2A), there was strong evidence (posterior probability (PP)>95%) for a shared causal variant underlying gene expression and protein levels (Additional File 1: Table S10). MR analyses provided evidence for reciprocal associations between changes in gene expression and circulating levels of these proteins (Additional File 1: Table S11). One protein had weak evidence for colocalisation (PDCD1LG2, PP=60%) and eight proteins had strong evidence for separate causal variants underlying gene expression and protein levels.

### Epigenome-wide studies of plasma protein correlates of Alzheimer’s disease

There were 778 individuals with DNA methylation and proteomic data in the Generation Scotland sample. The mean age of the sample was 60.2 (SD=8.8) years and 56.4% of the sample were female. Thirty-two CpGs were associated with the levels of 24 unique proteins (PIP>95%, Additional File 1: Table S12 and Additional File 2: Figure S4). The median correlation coefficient between measured protein levels was 0.15. The associations consisted of 11 *cis* CpG sites and 21 *trans* CpG loci (Figure 3). The cg07839457 probe in the *NLRC5* locus was associated with IL18BP, CSF1R and B2M levels, and the smoking-associated probe cg05575921 in *AHRR* (57-59) was associated with TREM2, PIGR, WFDC2 and RSPO4 levels.

**Figure 3.**
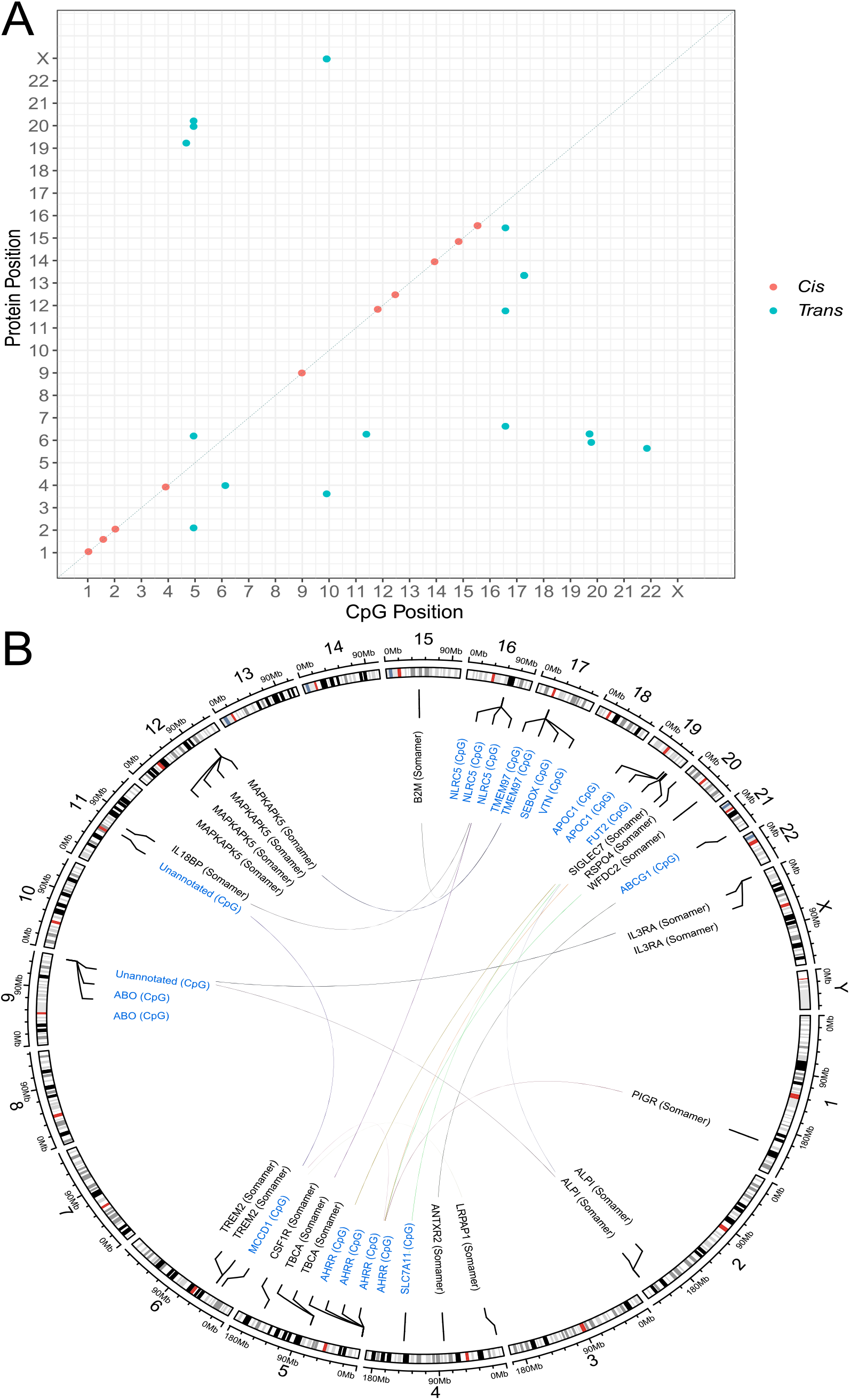
Epigenome-wide association studies of plasma protein levels previously associated with Alzheimer’s disease and disease-related phenotypes. (A) Chromosomal locations of CpGs identified through Bayesian penalised regression EWAS. The *x*-axis shows the chromosomal location of CpG sites and the *y*-axis represents the position of the gene encoding the target protein. *Cis* (red circles); *trans* (blue circles). (B) A circos plot for the 21 *trans*-associated CpGs from (A). Lines indicate an association between a CpG site and SOMAmer. EWAS, epigenome-wide association study.

We used linear mixed-effects models that accounted for relatedness to perform sensitivity analyses for the 32 CpG associations identified in BayesR+ (Additional File 1: Table S13) (60). Effect sizes were highly correlated with those from BayesR+ and showed full directional concordance (*r*=0.94, 95% CI = [0.88, 0.97], Additional File 2: Figure S5). Twenty-six associations were replicated at a genome-wide significance threshold of P < 3.6 × 10^−8^ (61) and the remaining six associations were replicated with P < 1.3 × 10^−4^. Further, 8/32 CpG associations were previously reported in the literature and effect sizes correlated strongly with those in our study (*r*=0.98, 95% CI = [0.87, 1.0]). The 24 novel CpG sites were associated with levels of 17 unique proteins.

Using methylation data, pathway enrichment was assessed among KEGG and GO terms (Additional File 1: Table S14 and Table S15, respectively, see Additional File 3 for Methods). Genes associated with the levels of six separate proteins in our EWAS were overrepresented in biological pathways using KEGG and GO terms. Genes associated with IL18BP levels were overrepresented in prolactin signalling and inflammation pathways. Gene sets identified in our EWAS of PIGR, RSPO4 and ANTXR2 levels were each associated with cancer pathways. Genes associated with B2M levels were enriched in viral response and antigen processing pathways. LRPAP1 levels were associated with immune cell-mediated cytotoxicity and antigen processing pathways. LRPAP1 harboured a *trans* CpG signal in the *MCCD1* locus within the major histocompatibility complex III region.

In BayesR+, estimates for the proportions of variability in SOMAmer levels that could be accounted for by DNA methylation measured on the EPIC BeadChip array ranged from 6.0% (ENO2; 95% CrI = [0%, 27.4%]) to 49.2% (MAPKAPK5; 95% CrI = [36.3%, 61.0%]), with a median estimate of 14.0% (Additional File 1: Table S16).

Estimates for variance in SOMAmer levels accounted for by genetic and methylation data together, while conditioned on each other, ranged from 21.8% for ENTPD1 (95% CrI = [0.0%, 59.1%]) to 93.3% for GHR (95% CrI = 80.1%, 100%]) (Additional File 1: Table S17 and Additional File 4). The mean and median estimates were 48.7% and 46.8%, respectively.

### Colocalisation of protein QTLs with methylation QTLs

Fifteen proteins had both genome-wide significant pQTL and CpG associations in our study. There were 45 possible SNP-CpG pairs across these proteins. For each pair, we used linear regression to test whether the SNP was associated with CpG methylation at P<5×10^−8^, thereby representing an mQTL effect (Additional File 1: Table S18, see Methods). We also performed look-up analyses of mQTL databases including GoDMC and phenoscanner (62-64). In instances where an mQTL effect was identified in more than one database, coefficients from the study with the largest sample size were brought forward for colocalisation analyses. Further, in instances where two or more mQTLs were associated with the same CpG site in a given locus, only the most significant mQTL was brought forward for colocalisation analyses (n = 20 mQTLs, 14 proteins, Additional File 1: Table S19).

For seven proteins, we observed strong evidence in *coloc* that a single causal variant may underpin differential DNA methylation levels and protein abundances (PP>95%, Additional File 1: Table S20). For five proteins (MATN3, ANXA2, F7, PLA2G2A and PCSK7), these associations involved *cis* pQTLs and mQTLs. A single *trans* variant in *APOE* was associated with *APOE* methylation and plasma TBCA levels. A *trans* variant in *FUT2* likely influenced *FUT2* methylation and measured ALPI levels. MR analyses provided evidence that relationships between methylation and protein levels were bidirectional (Additional File 1: Table S21).

For PCSK7, protein levels colocalised with DNA methylation and gene expression. Using *moloc*, there was strong evidence that these three traits colocalise with a common variant in the *PCSK7* locus (n_snps_= 543, PP>95%) (65).

### Causal associations between plasma proteins and Alzheimer’s disease risk

Bidirectional MR was applied to test for relationships between the 42 SOMAmers with pQTL associations in BayesR+ and 20 AD-related traits (Additional File 1: Table S22). A Bonferroni-corrected threshold of P<5.95 × 10^−5^ (<0.05/42 x 20) was set. Plasma levels of three proteins had a unidirectional association with AD risk: TREM2 (Table 1, Wald ratio test, *trans* pQTL in *MS4A4A*, beta=-0.13, se=0.05, P=8.43 × 10^−17^), CSF3 (Wald ratio test, 1 *trans* pQTL in *APOC2*, beta=0.10, se=0.01, P=7.62 × 10^−14^) and TBCA (inverse variance-weighted method, 2 *trans* pQTLs in *APOE*, beta=-0.50, se=0.12, P=1.20 × 10^−5^). Conversely, AD risk was not associated with plasma levels of these proteins. Colocalisation analyses provided evidence for one causal variant underlying TREM2 or TBCA levels and AD risk, and two separate causal variants underlying CSF3 levels and AD risk (Additional File 1: Table S23).

**Table 1.**
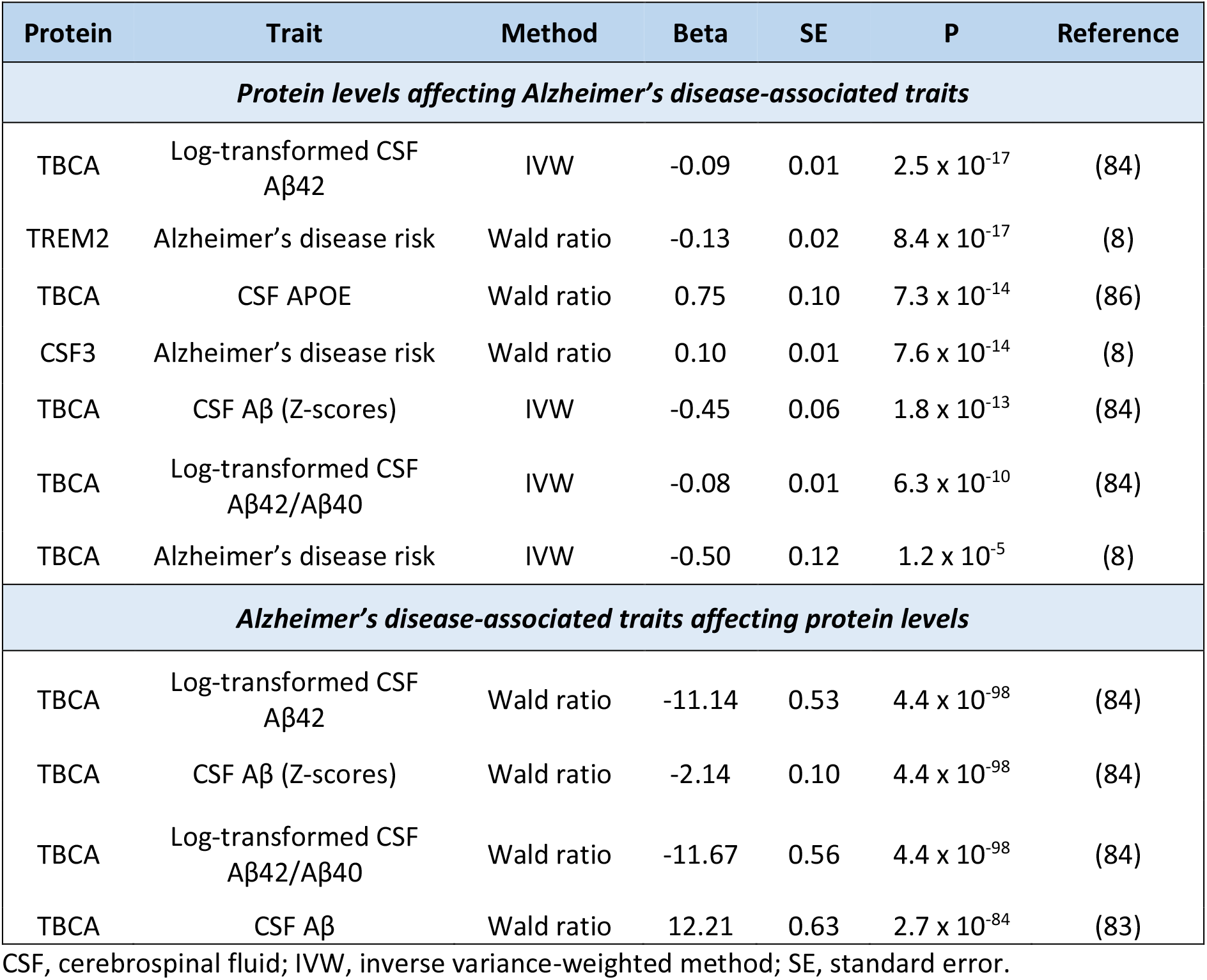
Mendelian randomisation analyses of plasma protein levels and Alzheimer’s disease-associated traits (Bonferroni-corrected P<5.95 × 10^−5^).

## Discussion

In this study, we identified eight novel protein QTLs and 24 novel CpG sites that associated with the levels of 23 AD-related plasma proteins. Using BayesR+, we provided estimates for associations between common genetic and DNAm variation and inter-individual differences in plasma levels of 281 AD-related proteins. We integrated our data with publicly-available gene expression and methylation QTL databases thereby highlighting molecular mechanisms that might causally link pQTLs to differential levels of eight proteins. This included a *trans* pQTL in *APOE* underpinning both DNAm variation in the *APOE* locus and TBCA levels. Using Mendelian randomisation and colocalisation analyses, we observed strong evidence for relationships between plasma levels of TREM2 or TBCA and AD risk. These associations were driven by *trans* pQTLs in *MS4A4A* and *APOE*, respectively.

For the first time, we show that the *trans* pQTL (rs1582763) in the *MS4A4A* locus associates with higher plasma TREM2 levels. This variant has been associated with higher CSF TREM2 levels and lower AD risk (8, 66). Furthermore, it is in moderate LD (*r*^2^ = 0.55) with a variant in the 3’UTR region of *MS4A6A* (rs610932) that was associated with serum TREM2 levels in a sample of 5,457 Icelanders (mean age = 75 years) (24). In a strategy that used human macrophages as proxies for microglia, this polymorphism was shown to alter *MS4A4A* expression and subsequently modulate TREM2 concentration (67). We also identified a novel blood CpG correlate of plasma TREM2 levels (cg02521229) located near *MS4A4A* that previously associated with dementia risk in Generation Scotland participants (68). Our data suggest that risk mechanisms arising from *MS4A4A* polymorphisms and TREM2 levels can be captured in plasma assays and that these mechanisms may involve differential methylation in the *MS4A4A* locus. We identified four other novel *trans* pQTLs. First, FPR (formyl peptide receptor) proteins regulate the phosphorylation of downstream MARCKS (myristoylated alanine-rich C-kinase substrate) proteins (69, 70). Second, the T allele of rs3820897 in *COLEC11* previously was associated with higher circulating inflammatory protein levels (22). In our study, this allele associated with lower levels of the anti-inflammatory protein ALPL (non-specific alkaline phosphatase isozyme) (22). Third, a *trans* variant in *BCHE* for KLK6 may represent a technical artefact as it is in moderate LD (*r*^2^ ∼ 0.5) with variants that increase the DNA-binding affinity of BCHE and therefore induce interactions with other SOMAmers (20). Fourth, we observed a novel *trans* association between rs1354034 (*ARHGEF3*) and YBX2 levels. The *ARHGEF3* locus influences platelet counts (71) and the rs1354034 variant was associated with >1,000 SOMAmers in a recent pQTL study (20). Individuals with this variant might have higher platelet counts or reactivity thereby resulting in the secretion of larger amounts of proteins from platelets during sample preparation.

We observed associations between plasma levels of three proteins (CSF3, MAPKAPK5 and TBCA) and *trans* pQTLs in the *TOMM40-APOE-APOC2* locus. Further, we identified two pQTLs and three CpG correlates of plasma MAPKAPK5 levels in the *TMEM97* locus. MAPKAPK5 correlated with cognitive decline in the Twins UK cohort, however its relationship with neuropathology is unknown (37). TMEM97 acts a synaptic receptor for amyloid-beta and mediates its cellular update via APOE-dependent and APOE-independent mechanisms (72, 73). Given that *TMEM97* polymorphisms may influence MAPKAPK5 levels, our data prioritise MAPKAPK5 for follow-up studies as a potential downstream effector or correlate of TMEM97 in amyloid clearance. TBCA correlates with amyloid-beta burden (29). TBCA levels are higher in individuals with the protective *APOE* ε2/ε2 genotype and lower in carriers of the risk ε4 polymorphism (74). These data are consistent with our GWAS and MR analyses. We supplement these data by identifying two novel CpG correlates of TBCA levels in *APOC1* and show the associations are likely mediated via mQTL effects. Future studies should examine whether TBCA dysregulation is a cause or consequence of disease risk mechanisms in carriers of *APOE* ε4 polymorphisms.

Our study has a number of limitations. First, our systematic review does not reflect an exhaustive list of potential AD-associated traits. Furthermore, there is heterogeneity across studies in terms of diagnostic criteria and phenotype definitions. Second, by focussing on the SOMAscan® platform alone, we do not capture all blood protein correlates of AD that are reported in the literature. Third, an insufficient number of variants were available to test for horizontal pleiotropy in Mendelian randomisation analyses. Fourth, some loci may have multiple SNPs that are in high LD. In this case, BayesR+ will interchangeably include and exclude closely-linked SNPs across iterations i.e. an individual SNP is selected in some iterations and inclusion probabilities will be shared across closely-linked SNPs. In regions where a SNP shows a high PIP (>95%), there is likely no other variant in LD with a causal variant. Therefore, if pQTLs with PIP>95% overlap with mQTLs for example then there is strong evidence for a single causal variant underlying both traits. Fifth, it is important to note that variants may alter SOMAmer reactivity with protein targets, or reflect technical artefacts such as sample handling and cross-reactive events (20). The replication of pQTL and CpG associations across proteomic platforms will help to disentangle biological from non-specific signals (19). Sixth, our sample consisted of Scottish individuals with a relatively homogenous genetic background thereby limiting generalisability of findings to individuals from other ethnic backgrounds.

## Conclusion

Our strategy of integrating multiple omics measures has determined the degree to which molecular factors can explain inter-individual differences in blood levels of possible biomarkers for AD and advanced understanding of mechanisms underlying AD risk.

## Methods

### Study Cohort

Analyses were performed using blood samples from participants of the **ST**ratifying **R**esilience **a**nd **D**epression **L**ongitudinally (STRADL) cohort, which comprises 1,188 individuals from the larger, family-structured Generation Scotland: the Scottish Family Health Study (GS). GS consists of 24,084 individuals from across Scotland, some of whom were members of the Walker Cohort in Dundee (75) and the Aberdeen Children of the 1950s study (76). Recruitment for GS took place between 2006 and 2011. Members of the STRADL cohort partook in follow-up data collection 4-13 years after baseline (77, 78). Of the original GS members, 5,649 were invited to take part in the STRADL cohort. There were 1,188 positive respondents. Participants were tested across two sites (n=582 and 606 from Aberdeen and Dundee, respectively).

### Search strategy and inclusion criteria

We searched MEDLINE (Ovid interface, Ovid MEDLINE in-process and other non-indexed citations and Ovid MEDLINE 1946 onwards), Embase (Ovid interface, 1980 onwards), Web of Science (core collection, Thomson Reuters) and medRxiv/bioRxiv to identify relevant articles indexed as of 28 May 2021. Search terms are outlined in Additional File 3. Twenty-five articles were identified and one further article was identified through a supplemental manual literature search. After removal of duplicates, 23 articles were assessed for inclusion criteria: (i) original research article, (ii) proteins were measured in plasma, (iii) proteins were measured using SOMAscan® technology and (iv) proteins were associated with Alzheimer’s disease or related phenotypes. Twelve articles met inclusion criteria.

### Protein measurements in Generation Scotland

The 5k SOMAscan® v4 array was used to quantify the levels of plasma proteins in Generation Scotland participants (n=1,065). This highly multiplexed platform uses chemically modified aptamers termed SOMAmers (**S**low **O**ff-rate **M**odified **A**pta**mers**) that recognise epitopes on their cognate protein targets with high specificity and high affinity in the nanomolar-to-picomolar range. The recognition signal is measured as relative fluorescence units (RFUs) on microarrays (79).

Plasma samples were collected in 150 µl aliquots and stored at -80°C. Samples were run in 96-well plates and reagents were spread across three dilution factors (0.005%, 0.5%, and 20%) to create distinct sets for high, medium, and low abundance proteins, respectively. Raw microarray data were normalised through a number of quality control steps, which are detailed in Additional File 3 (80). After quality control and the exclusion of non‐human proteins, deprecated markers and spuriomers, 4,235 SOMAmers were retained for proteomic analyses.

Normalised RFUs (from Somalogic®) were first log-transformed and regressed onto the following covariates: age, sex, study site (Aberdeen/Dundee), time between sample being collected and sample being processed for proteomics (factor, 4 levels), estimated glomerular filtration rate and 20 genetic principal components (PCs) of ancestry from multi-dimensional scaling (to control for population structure). Relationships between covariates and SOMAmers are shown in Additional File 1: Table S24. Residualised RFUs were transformed by rank-based inverse normalisation. We refer to these as protein levels; however, they reflect RFUs which have undergone a number of quality control, transformation and pre-correction steps.

### Genome-wide association studies

Generation Scotland samples were genotyped using the Illumina Human OmniExpressExome-8v1.0 Bead Chip and processed using the Illumina Genome Studio software v2011 (Illumina, San Diego, CA, USA) (81). Quality control steps are outlined in Additional File 3. After quality control, 561,125 SNPs remained for 1,064 individuals. In total, 1,064 individuals had both genotype and proteomic data available for analyses.

Bayesian penalised regression GWAS were performed using BayesR+ software in C++ (41). BayesR+ utilises a mixture of prior Gaussian distributions to allow for markers with effect sizes of different magnitudes. It also includes a discrete spike at zero that enables the exclusion of markers with non-identifiable effects on the trait of interest. Guided by data from our previous studies, mixture variances for the stand-alone GWAS were set to 0.01 and 0.1 to allow for markers that account for 1% or 10% of variation in circulating protein levels, respectively (27, 28). In the combined GWAS/EWAS analysis, genotype and DNAm data must have had the same number of prior variances (n=3 each). Therefore, mixture variances for SNP data were set to 0.01, 0.1 and 0.2 in the combined analyses. Input data were scaled to mean zero and unit variance. Gibbs sampling was used to sample over the posterior distribution conditional on input data and 10,000 samples were used. The first 5,000 samples of burn-in were removed and a thinning of 5 samples was applied to reduce autocorrelation. SNPs which exhibited a posterior inclusion probability ≥95% were deemed significant.

### Epigenome-wide association studies

Blood DNAm in Generation Scotland participants was assessed using the Illumina HumanMethylationEPIC BeadChip Array. Blood DNAm was assessed in two separate sets. After quality control, 793,706 and 773,860 CpG remained in sets 1 and 2, respectively. In total, 772,619 CpG sites were shared across sets. Each set was truncated to these overlapping probes.

In the stand-alone EWAS and combined GWAS/EWAS, mixture variances were set to 0.001, 0.01 and 0.1 (n=778). Missing DNAm data were mean imputed separately within each set as BayesR+ cannot accept missing values. Both sets were then combined and adjusted for DNAm batch, set, age and sex. Each CpG site was scaled to mean zero and unit variance. Houseman-estimated white blood cell proportions were included as fixed effect covariates in EWAS models (82). CpG sites that had a posterior inclusion probability ≥95% were deemed significant.

Linear mixed-effects models were performed in sensitivity EWAS analyses using the lmekin function from the *coxme* package in R (version 2.2-16) (60). DNAm data were pre-corrected for age, sex, batch and set. Houseman-estimated white blood cell proportions were incorporated as fixed effect covariates and a kinship matrix was fitted to account for relatedness among individuals in the family-based STRADL cohort.

### Colocalisation analyses

Formal Bayesian tests of colocalisation were used to determine whether a shared causal variant likely underpinned two traits of interest (56). For each protein, a 200 kilobase region (upstream and downstream—recommended default setting) surrounding the variant was extracted from our GWAS summary statistics.

Expression QTL data were extracted from eQTLGen summary statistics. Methylation QTL summary statistics were extract from phenoscanner, GoDMC or our own mQTL analyses. Methylation QTL analyses were performed using additive linear regression models and by regressing CpG sites (beta values) on SNPs (0, 1, 2) while adjusting for age, sex, DNAm batch, set, Houseman-estimated white blood cell proportions and 20 genetic PCs (n=778). In instances where an mQTL effect was identified in more than one database, summary statistics from the study with the largest sample size were used in *coloc* (55, 62, 63). For AD-related traits, summary statistics were extracted from the relevant GWAS (8, 83, 84). Default priors were applied. Summary statistics for all SNPs (±200 kilobases from the queried SNP) were used to estimate the posterior probability for five separate hypotheses: a single causal variant for both traits, separate causal variants for both traits, a causal variant for just one trait (encompassing two hypotheses), or no causal variant for either trait. Posterior probabilities ≥95% provided strong evidence for a given hypothesis.

### Mendelian randomisation

Bidirectional Mendelian randomisation was used to test for possibly causal relationships between (i) gene expression and plasma protein levels, (ii) DNAm and plasma protein levels and (iii) plasma protein levels and AD risk or related biomarkers. Pruned variants (*r*^2^<0.1) were used as instrumental variables (IVs) in MR analyses. In tests where only one independent variant remained after pruning, effect size estimates were assessed using the Wald ratio test. In tests where two SNPs remained, analyses were performed using the inverse variance-weighted method. Analyses were conducted using MR-base (85). Two-sample MR was used with the exception of 3 mQTL-pQTL associations derived from our own summary statistics (F7, PLA2G2A, PCSK7) and these relationships were assessed using the Wald ratio test. Further information on IVs used are provided in Additional File 3.

### Ethics approval and consent to participate

All components of the Generation Scotland study received ethical approval from the NHS Tayside Committee on Medical Research Ethics (REC Reference Numbers: 05/S1401/89 and 10/S1402/20). All participants provided broad and enduring written informed consent for biomedical research. Generation Scotland has also been granted Research Tissue Bank status by the East of Scotland Research Ethics Service (REC Reference Number: 20-ES-0021). This study was performed in accordance with the Helsinki declaration.

## Supporting information

Additional File 4 - Combined Variance Components BayesR

## Data Availability

According to the terms of consent for Generation Scotland participants, access to data must be reviewed by the Generation Scotland Access Committee. Applications should be made to. All code is available at the following Github repository: https://github.com/robertfhillary/gwas_ewas_AD_plasma_biomarkers.

## Acknowledgments

We are grateful to the families who took part in this study, the general practitioners and the Scottish School of Primary Care for their help in recruiting them and the wider Generation Scotland team. Generation Scotland received core support from the Chief Scientist Office of the Scottish Government Health Directorates [CZD/16/6] and the Scottish Funding Council [HR03006]. Genotyping and DNA methylation profiling of the Generation Scotland samples was carried out by the Genetics Core Laboratory at the Wellcome Trust Clinical Research Facility, Edinburgh, Scotland and was funded by the Medical Research Council UK and the Wellcome Trust (Wellcome Trust Strategic Award “STratifying Resilience and Depression Longitudinally” ((STRADL) Reference [104036/Z/14/Z])). A.M.M is supported by Wellcome [104036/Z/14/Z, 216767/Z/19/Z, 220857/Z/20/Z], UKRI MRC [MC_PC_17209, MR/S035818/1] and the European Union H2020 [SEP-210574971]. R.F.H and D.A.G are supported by funding from the Wellcome 4-year PhD in Translational Neuroscience – training the next generation of basic neuroscientists to embrace clinical research [108890/Z/15/Z]. D.L.McC and R.E.M are supported by Alzheimer’s Research UK major project grant ARUK-PG2017B-10. Proteomic analyses in STRADL were supported by Dementias Platform UK (DPUK). DPUK funded this work through core grant support from the Medical Research Council [MR/L023784/2]. C.H. is supported by an MRC University Unit Programme Grant MC_UU_00007/10 (QTL in Health and Disease). L.S. is funded by DPUK through MRC [MR/L023784/2] and the UK Medical Research Council Award to the University of Oxford [MC_PC_17215]. L.S. received support from the NIHR Biomedical Research Centre at Oxford Health NHS Foundation Trust.

## Data availability

According to the terms of consent for Generation Scotland participants, access to data must be reviewed by the Generation Scotland Access Committee. Applications should be made to.

## Code availability

All code is available at the following Github repository: https://github.com/robertfhillary/gwas_ewas_AD_plasma_biomarkers.

## References

1. 2020 GHE. Deaths by Cause, Age, Sex, by Country and by Region, 2000-2019. Geneva: World Health Organization. 2020.

2. DALYs GBD, Collaborators H. Global, regional, and national disability-adjusted life-years (DALYs) for 333 diseases and injuries and healthy life expectancy (HALE) for 195 countries and territories, 1990-2016: a systematic analysis for the Global Burden of Disease Study 2016. Lancet (London, England). 2017;390(10100):1260–344.

3. Nakamura A, Kaneko N, Villemagne VL, Kato T, Doecke J, Doré V, et al. High performance plasma amyloid-β biomarkers for Alzheimer’s disease. Nature. 2018;554(7691):249–54.

4. Kiddle SJ, Voyle N, Dobson RJB. A Blood Test for Alzheimer’s Disease: Progress, Challenges, and Recommendations. Journal of Alzheimer’s disease: JAD. 2018;64(1):S289–S97.

5. Mielke MM, Hagen CE, Xu J, Chai X, Vemuri P, Lowe VJ, et al. Plasma phospho-tau181 increases with Alzheimer’s disease clinical severity and is associated with tau- and amyloid-positron emission tomography. Alzheimer’s & dementia: the journal of the Alzheimer’s Association. 2018;14(8):989–97.

6. Palmqvist S, Insel PS, Stomrud E, Janelidze S, Zetterberg H, Brix B, et al. Cerebrospinal fluid and plasma biomarker trajectories with increasing amyloid deposition in Alzheimer’s disease. EMBO molecular medicine. 2019;11(12):e11170.

7. Shi L, Baird AL, Westwood S, Hye A, Dobson R, Thambisetty M, et al. A Decade of Blood Biomarkers for Alzheimer’s Disease Research: An Evolving Field, Improving Study Designs, and the Challenge of Replication. Journal of Alzheimer’s disease: JAD. 2018;62(3):1181–98.

8. Jansen IE, Savage JE, Watanabe K, Bryois J, Williams DM, Steinberg S, et al. Genome-wide meta-analysis identifies new loci and functional pathways influencing Alzheimer’s disease risk. Nature genetics. 2019;51(3):404–13.

9. Wightman DP, Jansen IE, Savage JE, Shadrin AA, Bahrami S, Rongve A, et al. Largest GWAS (N=1,126,563) of Alzheimer’s Disease Implicates Microglia and Immune Cells. medRxiv. 2020:2020.11.20.20235275.

10. Olsson B, Lautner R, Andreasson U, Öhrfelt A, Portelius E, Bjerke M, et al. CSF and blood biomarkers for the diagnosis of Alzheimer’s disease: a systematic review and meta-analysis. The Lancet Neurology. 2016;15(7):673–84.

11. Lista S, Faltraco F, Prvulovic D, Hampel H. Blood and plasma-based proteomic biomarker research in Alzheimer’s disease. Prog Neurobiol. 2013;101-102:1–17.

12. Zetterberg H, Burnham SC. Blood-based molecular biomarkers for Alzheimer’s disease. Molecular Brain. 2019;12(1):26.

13. Sun BB, Maranville JC, Peters JE, Stacey D, Staley JR, Blackshaw J, et al. Genomic atlas of the human plasma proteome. Nature. 2018;558(7708):73–9.

14. Yao C, Chen G, Song C, Keefe J, Mendelson M, Huan T, et al. Genome‐wide mapping of plasma protein QTLs identifies putatively causal genes and pathways for cardiovascular disease. Nature communications. 2018;9(1):3268.

15. Emilsson V, Ilkov M, Lamb JR, Finkel N, Gudmundsson EF, Pitts R, et al. Co-regulatory networks of human serum proteins link genetics to disease. Science (New York, NY). 2018;361(6404):769–73.

16. Kibinge NK, Relton CL, Gaunt TR, Richardson TG. Characterizing the Causal Pathway for Genetic Variants Associated with Neurological Phenotypes Using Human Brain-Derived Proteome Data. The American Journal of Human Genetics. 2020;106(6):885–92.

17. Fraga MF, Ballestar E, Paz MF, Ropero S, Setien F, Ballestar ML, et al. Epigenetic differences arise during the lifetime of monozygotic twins. Proc Natl Acad Sci U S A. 2005;102(30):10604–9.

18. Lourdusamy A, Newhouse S, Lunnon K, Proitsi P, Powell J, Hodges A, et al. Identification of cis-regulatory variation influencing protein abundance levels in human plasma. Human molecular genetics. 2012;21(16):3719–26.

19. Pietzner M, Wheeler E, Carrasco-Zanini J, Kerrison ND, Oerton E, Koprulu M, et al. Cross-platform proteomics to advance genetic prioritisation strategies. bioRxiv. 2021:2021.03.18.435919.

20. Pietzner M, Wheeler E, Carrasco-Zanini J, Raffler J, Kerrison ND, Oerton E, et al. Genetic architecture of host proteins involved in SARS-CoV-2 infection. Nature communications. 2020;11(1):6397.

21. Di Narzo AF, Telesco SE, Brodmerkel C, Argmann C, Peters LA, Li K, et al. High-Throughput Characterization of Blood Serum Proteomics of IBD Patients with Respect to Aging and Genetic Factors. PLoS genetics. 2017;13(1):e1006565.

22. Suhre K, Arnold M, Bhagwat AM, Cotton RJ, Engelke R, Raffler J, et al. Connecting genetic risk to disease end points through the human blood plasma proteome. Nature communications. 2017;8:14357.

23. Carayol J, Chabert C, Di Cara A, Armenise C, Lefebvre G, Langin D, et al. Protein quantitative trait locus study in obesity during weight-loss identifies a leptin regulator. Nature communications. 2017;8(1):2084.

24. Emilsson V, Gudmundsdottir V, Ilkov M, Staley JR, Gudjonsson A, Gudmundsson EF, et al. Human serum proteome profoundly overlaps with genetic signatures of disease. bioRxiv. 2020:2020.05.06.080440.

25. Zaghlool SB, Kühnel B, Elhadad MA, Kader S, Halama A, Thareja G, et al. Epigenetics meets proteomics in an epigenome-wide association study with circulating blood plasma protein traits. Nature communications. 2020;11(1):15.

26. Ahsan M, Ek WE, Rask-Andersen M, Karlsson T, Lind-Thomsen A, Enroth S, et al. The relative contribution of DNA methylation and genetic variants on protein biomarkers for human diseases. PLoS genetics. 2017;13(9):e1007005–e.

27. Hillary RF, Trejo-Banos D, Kousathanas A, McCartney DL, Harris SE, Stevenson AJ, et al. Multi-method genome- and epigenome-wide studies of inflammatory protein levels in healthy older adults. Genome medicine. 2020;12(1):60.

28. Hillary RF, McCartney DL, Harris SE, Stevenson AJ, Seeboth A, Zhang Q, et al. Genome and epigenome wide studies of neurological protein biomarkers in the Lothian Birth Cohort 1936. Nature communications. 2019;10(1):3160.

29. Shi L, Westwood S, Baird AL, Winchester L, Dobricic V, Kilpert F, et al. Discovery and validation of plasma proteomic biomarkers relating to brain amyloid burden by SOMAscan assay. Alzheimer’s & dementia: the journal of the Alzheimer’s Association. 2019;15(11):1478–88.

30. Shi L, Winchester LM, Liu BY, Killick R, Ribe EM, Westwood S, et al. Dickkopf-1 Overexpression in vitro Nominates Candidate Blood Biomarkers Relating to Alzheimer’s Disease Pathology. Journal of Alzheimer’s disease: JAD. 2020;77(3):1353–68.

31. Tanaka T, Lavery R, Varma V, Fantoni G, Colpo M, Thambisetty M, et al. Plasma proteomic signatures predict dementia and cognitive impairment. Alzheimer’s & Dementia: Translational Research & Clinical Interventions. 2020;6(1):e12018.

32. Sattlecker M, Kiddle SJ, Newhouse S, Proitsi P, Nelson S, Williams S, et al. Alzheimer’s disease biomarker discovery using SOMAscan multiplexed protein technology. Alzheimer’s & dementia: the journal of the Alzheimer’s Association. 2014;10(6):724–34.

33. Kiddle SJ, Sattlecker M, Proitsi P, Simmons A, Westman E, Bazenet C, et al. Candidate blood proteome markers of Alzheimer’s disease onset and progression: a systematic review and replication study. Journal of Alzheimer’s disease: JAD. 2014;38(3):515–31.

34. Sattlecker M, Khondoker M, Proitsi P, Williams S, Soininen H, Kłoszewska I, et al. Longitudinal Protein Changes in Blood Plasma Associated with the Rate of Cognitive Decline in Alzheimer’s Disease. Journal of Alzheimer’s disease: JAD. 2016;49(4):1105–14.

35. Begic E, Hadzidedic S, Kulaglic A, Ramic-Brkic B, Begic Z, Causevic M. SOMAscan-based proteomic measurements of plasma brain natriuretic peptide are decreased in mild cognitive impairment and in Alzheimer’s dementia patients. PloS one. 2019;14(2):e0212261.

36. Lindbohm JV, Mars N, Walker KA, Singh-Manoux A, Livingston G, Brunner EJ, et al. Association of plasma proteins with rate of cognitive decline and dementia: 20-year follow-up of the Whitehall II and ARIC cohort studies. medRxiv. 2020:2020.11.18.20234070.

37. Kiddle SJ, Steves CJ, Mehta M, Simmons A, Xu X, Newhouse S, et al. Plasma protein biomarkers of Alzheimer’s disease endophenotypes in asymptomatic older twins: early cognitive decline and regional brain volumes. Translational psychiatry. 2015;5(6):e584.

38. Begic E, Hadzidedic S, Obradovic S, Begic Z, Causevic M. Increased Levels of Coagulation Factor XI in Plasma Are Related to Alzheimer’s Disease Diagnosis. Journal of Alzheimer’s disease: JAD. 2020;77(1):375–86.

39. Shi L, Winchester LM, Westwood S, Baird AL, Anand SN, Buckley NJ, et al. Replication study of plasma proteins relating to Alzheimer’s pathology. Alzheimer’s & dementia: the journal of the Alzheimer’s Association. 2021.

40. Walker KA, Chen J, Zhang J, Fornage M, Yang Y, Zhou L, et al. Large-scale plasma proteomic analysis identifies proteins and pathways associated with dementia risk. Nature Aging. 2021;1(5):473–89.

41. Trejo Banos D, McCartney DL, Patxot M, Anchieri L, Battram T, Christiansen C, et al. Bayesian reassessment of the epigenetic architecture of complex traits. Nature communications. 2020;11(1):2865.

42. McLaren W, Gil L, Hunt SE, Riat HS, Ritchie GRS, Thormann A, et al. The Ensembl Variant Effect Predictor. Genome biology. 2016;17(1):122.

43. Sun W, Kechris K, Jacobson S, Drummond MB, Hawkins GA, Yang J, et al. Common Genetic Polymorphisms Influence Blood Biomarker Measurements in COPD. PLoS genetics. 2016;12(8):e1006011.

44. Ruffieux H, Carayol J, Popescu R, Harper M-E, Dent R, Saris WHM, et al. A fully joint Bayesian quantitative trait locus mapping of human protein abundance in plasma. PLoS Comput Biol. 2020;16(6):e1007882–e.

45. Nath AP, Ritchie SC, Grinberg NF, Tang HH, Huang QQ, Teo SM, et al. Multivariate Genome-wide Association Analysis of a Cytokine Network Reveals Variants with Widespread Immune, Haematological, and Cardiometabolic Pleiotropy. Am J Hum Genet. 2019;105(6):1076–90.

46. Folkersen L, Fauman E, Sabater-Lleal M, Strawbridge RJ, Frånberg M, Sennblad B, et al. Mapping of 79 loci for 83 plasma protein biomarkers in cardiovascular disease. PLoS genetics. 2017;13(4):e1006706–e.

47. Innocenti F, Jiang C, Sibley AB, Etheridge AS, Hatch AJ, Denning S, et al. Genetic variation determines VEGF-A plasma levels in cancer patients. Scientific reports. 2018;8(1):16332.

48. Hoglund J, Rafati N, Rask-Andersen M, Enroth S, Karlsson T, Ek WE, et al. Improved power and precision with whole genome sequencing data in genome-wide association studies of inflammatory biomarkers. Scientific reports. 2019;9(1):16844.

49. Ho JE, Mahajan A, Chen M-H, Larson MG, McCabe EL, Ghorbani A, et al. Clinical and genetic correlates of growth differentiation factor 15 in the community. Clinical chemistry. 2012;58(11):1582–91.

50. Sliz E, Kalaoja M, Ahola-Olli A, Raitakari O, Perola M, Salomaa V, et al. Genome-wide association study identifies seven novel loci associating with circulating cytokines and cell adhesion molecules in Finns. Journal of medical genetics. 2019;56(9):607–16.

51. Buniello A, MacArthur JAL, Cerezo M, Harris LW, Hayhurst J, Malangone C, et al. The NHGRI-EBI GWAS Catalog of published genome-wide association studies, targeted arrays and summary statistics 2019. Nucleic Acids Res. 2019;47(D1):D1005–d12.

52. Jun G, Ibrahim-Verbaas CA, Vronskaya M, Lambert JC, Chung J, Naj AC, et al. A novel Alzheimer disease locus located near the gene encoding tau protein. Molecular psychiatry. 2016;21(1):108–17.

53. Cruchaga C, Kauwe JS, Harari O, Jin SC, Cai Y, Karch CM, et al. GWAS of cerebrospinal fluid tau levels identifies risk variants for Alzheimer’s disease. Neuron. 2013;78(2):256–68.

54. Nazarian A, Yashin AI, Kulminski AM. Genome-wide analysis of genetic predisposition to Alzheimer’s disease and related sex disparities. Alzheimers Res Ther. 2019;11(1):5.

55. Võsa U, Claringbould A, Westra H-J, Bonder MJ, Deelen P, Zeng B, et al. Unraveling the polygenic architecture of complex traits using blood eQTL metaanalysis. bioRxiv. 2018:447367.

56. Giambartolomei C, Vukcevic D, Schadt EE, Franke L, Hingorani AD, Wallace C, et al. Bayesian test for colocalisation between pairs of genetic association studies using summary statistics. PLoS genetics. 2014;10(5):e1004383.

57. Joehanes R, Just AC, Marioni RE, Pilling LC, Reynolds LM, Mandaviya PR, et al. Epigenetic Signatures of Cigarette Smoking. Circ Cardiovasc Genet. 2016;9(5):436–47.

58. Zeilinger S, Kühnel B, Klopp N, Baurecht H, Kleinschmidt A, Gieger C, et al. Tobacco smoking leads to extensive genome-wide changes in DNA methylation. PloS one. 2013;8(5):e63812.

59. Philibert RA, Beach SR, Brody GH. Demethylation of the aryl hydrocarbon receptor repressor as a biomarker for nascent smokers. Epigenetics. 2012;7(11):1331–8.

60. Therneau TM. coxme: Mixed Effects Cox Models. R package version 2.2-16. 2020.

61. Saffari A, Silver MJ, Zavattari P, Moi L, Columbano A, Meaburn EL, et al. Estimation of a significance threshold for epigenome-wide association studies. Genet Epidemiol. 2018;42(1):20–33.

62. Staley JR, Blackshaw J, Kamat MA, Ellis S, Surendran P, Sun BB, et al. PhenoScanner: a database of human genotype-phenotype associations. Bioinformatics (Oxford, England). 2016;32(20):3207–9.

63. Min JL, Hemani G, Hannon E, Dekkers KF, Castillo-Fernandez J, Luijk R, et al. Genomic and phenomic insights from an atlas of genetic effects on DNA methylation. medRxiv. 2020:2020.09.01.20180406.

64. Hannon E, Gorrie-Stone TJ, Smart MC, Burrage J, Hughes A, Bao Y, et al. Leveraging DNA-Methylation Quantitative-Trait Loci to Characterize the Relationship between Methylomic Variation, Gene Expression, and Complex Traits. Am J Hum Genet. 2018;103(5):654–65.

65. Giambartolomei C, Zhenli Liu J, Zhang W, Hauberg M, Shi H, Boocock J, et al. A Bayesian framework for multiple trait colocalization from summary association statistics. Bioinformatics (Oxford, England). 2018;34(15):2538–45.

66. Liu C, Yu J. Genome-Wide Association Studies for Cerebrospinal Fluid Soluble TREM2 in Alzheimer’s Disease. Frontiers in aging neuroscience. 2019;11:297.

67. Deming Y, Filipello F, Cignarella F, Cantoni C, Hsu S, Mikesell R, et al. The MS4A gene cluster is a key modulator of soluble TREM2 and Alzheimer’s disease risk. Science translational medicine. 2019;11(505).

68. Walker RM, Bermingham ML, Vaher K, Morris SW, Clarke T-K, Bretherick AD, et al. Epigenome-wide analyses identify DNA methylation signatures of dementia risk. Alzheimer’s & dementia (Amsterdam, Netherlands). 2020;12(1):e12078–e.

69. Cattaneo F, Russo R, Castaldo M, Chambery A, Zollo C, Esposito G, et al. Phosphoproteomic analysis sheds light on intracellular signaling cascades triggered by Formyl-Peptide Receptor 2. Scientific reports. 2019;9(1):17894.

70. Ammendola R, Parisi M, Esposito G, Cattaneo F. Pro-Resolving FPR2 Agonists Regulate NADPH Oxidase-Dependent Phosphorylation of HSP27, OSR1, and MARCKS and Activation of the Respective Upstream Kinases. Antioxidants (Basel). 2021;10(1):134.

71. Zou S, Teixeira AM, Kostadima M, Astle WJ, Radhakrishnan A, Simon LM, et al. SNP in human ARHGEF3 promoter is associated with DNase hypersensitivity, transcript level and platelet function, and Arhgef3 KO mice have increased mean platelet volume. PloS one. 2017;12(5):e0178095.

72. Riad A, Lengyel-Zhand Z, Zeng C, Weng C-C, Lee VMY, Trojanowski JQ, et al. The Sigma-2 Receptor/TMEM97, PGRMC1, and LDL Receptor Complex Are Responsible for the Cellular Uptake of Aβ42 and Its Protein Aggregates. Molecular neurobiology. 2020;57(9):3803–13.

73. Colom-Cadena M, Tulloch J, Rose J, Smith C, Spires-Jones T. TMEM97 is a potential amyloid beta receptor in human Alzheimer’s disease synapses. Alzheimer’s & Dementia. 2020;16(S2):e041782.

74. Sebastiani P, Monti S, Morris M, Gurinovich A, Toshiko T, Andersen SL, et al. A serum protein signature of APOE genotypes in centenarians. Aging Cell. 2019;18(6):e13023.

75. Libby G, Smith A, McEwan NF, Chien PF, Greene SA, Forsyth JS, et al. The Walker Project: a longitudinal study of 48,000 children born 1952-1966 (aged 36-50 years in 2002) and their families. Paediatric and perinatal epidemiology. 2004;18(4):302–12.

76. Batty GD, Morton SM, Campbell D, Clark H, Smith GD, Hall M, et al. The Aberdeen Children of the 1950s cohort study: background, methods and follow-up information on a new resource for the study of life course and intergenerational influences on health. Paediatric and perinatal epidemiology. 2004;18(3):221–39.

77. Habota T, Sandu A, Waiter G, McNeil C, Steele J, Macfarlane J, et al. Cohort profile for the STratifying Resilience and Depression Longitudinally (STRADL) study: A depression-focused investigation of Generation Scotland, using detailed clinical, cognitive, and neuroimaging assessments [version 1; peer review: 1 approved, 1 not approved]. Wellcome Open Research. 2019;4(185).

78. Navrady LB, Wolters MK, MacIntyre DJ, Clarke T-K, Campbell AI, Murray AD, et al. Cohort Profile: Stratifying Resilience and Depression Longitudinally (STRADL): a questionnaire follow-up of Generation Scotland: Scottish Family Health Study (GS:SFHS). International journal of epidemiology. 2017;47(1):13–4g.

79. Gold L, Ayers D, Bertino J, Bock C, Bock A, Brody E, et al. Aptamer-based multiplexed proteomic technology for biomarker discovery. Nature Precedings. 2010:1-.

80. Candia J, Cheung F, Kotliarov Y, Fantoni G, Sellers B, Griesman T, et al. Assessment of Variability in the SOMAscan Assay. Scientific reports. 2017;7(1):14248.

81. Smith BH, Campbell A, Linksted P, Fitzpatrick B, Jackson C, Kerr SM, et al. Cohort Profile: Generation Scotland: Scottish Family Health Study (GS:SFHS). The study, its participants and their potential for genetic research on health and illness. International journal of epidemiology. 2013;42(3):689–700.

82. Houseman EA, Accomando WP, Koestler DC, Christensen BC, Marsit CJ, Nelson HH, et al. DNA methylation arrays as surrogate measures of cell mixture distribution. BMC bioinformatics. 2012;13:86.

83. Deming Y, Li Z, Kapoor M, Harari O, Del-Aguila JL, Black K, et al. Genome-wide association study identifies four novel loci associated with Alzheimer’s endophenotypes and disease modifiers. Acta neuropathologica. 2017;133(5):839–56.

84. Hong S, Prokopenko D, Dobricic V, Kilpert F, Bos I, Vos SJB, et al. Genome-wide association study of Alzheimer’s disease CSF biomarkers in the EMIF-AD Multimodal Biomarker Discovery dataset. Translational psychiatry. 2020;10(1):403.

85. Hemani G, Zheng J, Elsworth B, Wade KH, Haberland V, Baird D, et al. The MR-Base platform supports systematic causal inference across the human phenome. eLife. 2018;7.

86. Kauwe JS, Bailey MH, Ridge PG, Perry R, Wadsworth ME, Hoyt KL, et al. Genome-wide association study of CSF levels of 59 alzheimer’s disease candidate proteins: significant associations with proteins involved in amyloid processing and inflammation. PLoS genetics. 2014;10(10):e1004758.

