## Additional File 2 - Supplementary Figures for "Genome and epigenome wide studies of plasma protein biomarkers for Alzheimer’s disease implicate TBCA and TREM2 in disease risk"

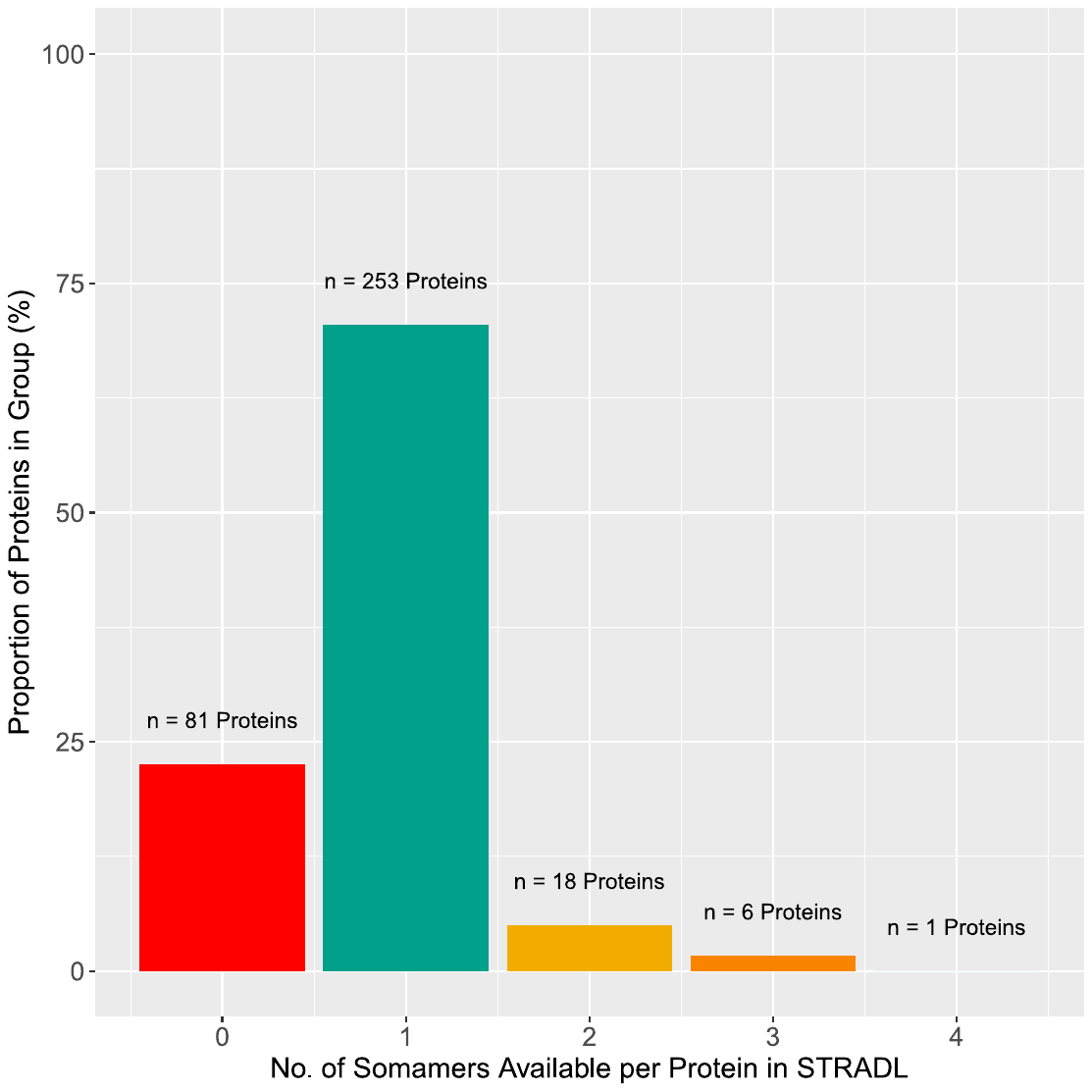

**Figure S1. Number of SOMAmers in the STRADL dataset that targeted Alzheimer’s disease-associated proteins in the literature.** In total, 359 unique protein targets (or SOMAmers) were previously associated with Alzheimer’s disease or relevant phenotypes in the literature. Of these, 278/359 (77.4%) SOMAmers had at least one corresponding SOMAmer that passed quality control in the STRADL dataset. In some instances, as shown in the figures, a given protein had more than one corresponding SOMAmer that could target it. This is because some proteins have a number of SOMAmers that can recognise different portions of the polypeptide sequence.

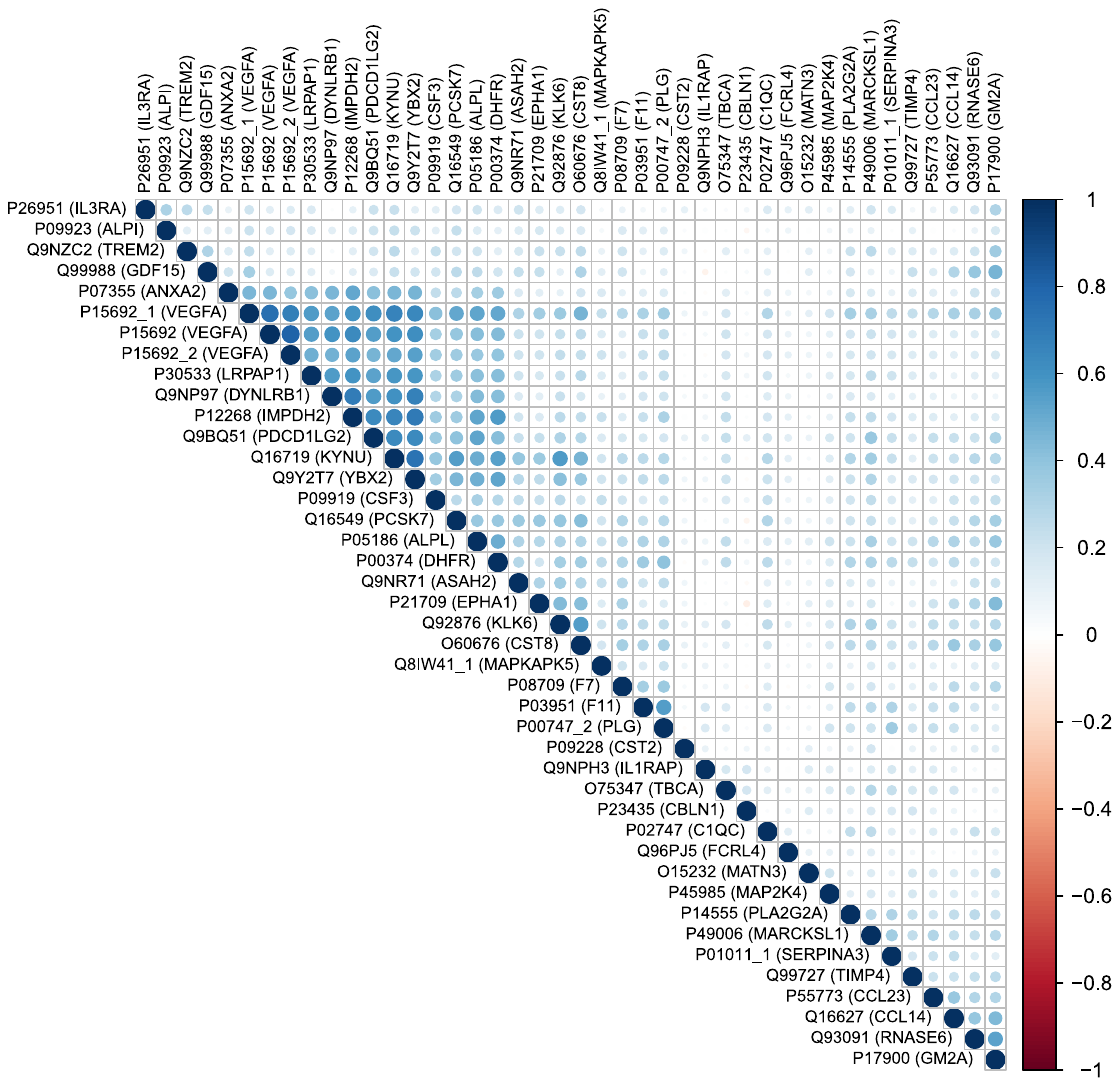

**Figure S2. Phenotypic correlations between 42 SOMAmer targets that exhibited pQTL associations in BayesR+.** Correlations were assessed using Pearson’s correlation. The interrelationships between SOMAmer levels in Generation Scotland (STRADL cohort) are shown. pQTL, protein quantitative trait locus.

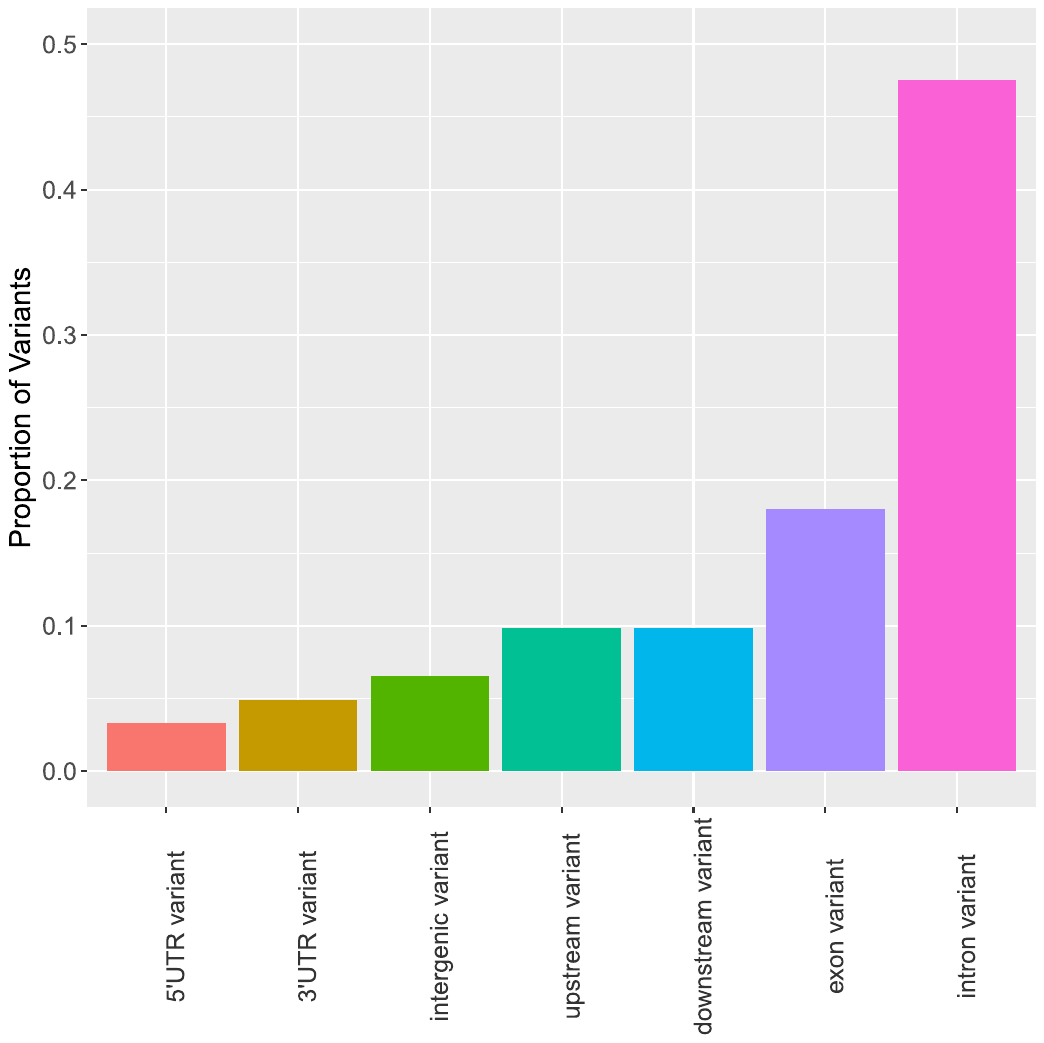

**Figure S3. Proportion of pQTL variants annotated to different genomic features.** The majority of pQTL variants (n=61) associated with plasma protein levels in BayesR+ were annotated to intronic regions (47.5%). Variants were annotated using Ensembl Variant Effect Predictor. pQTL, protein quantitative trait locus.

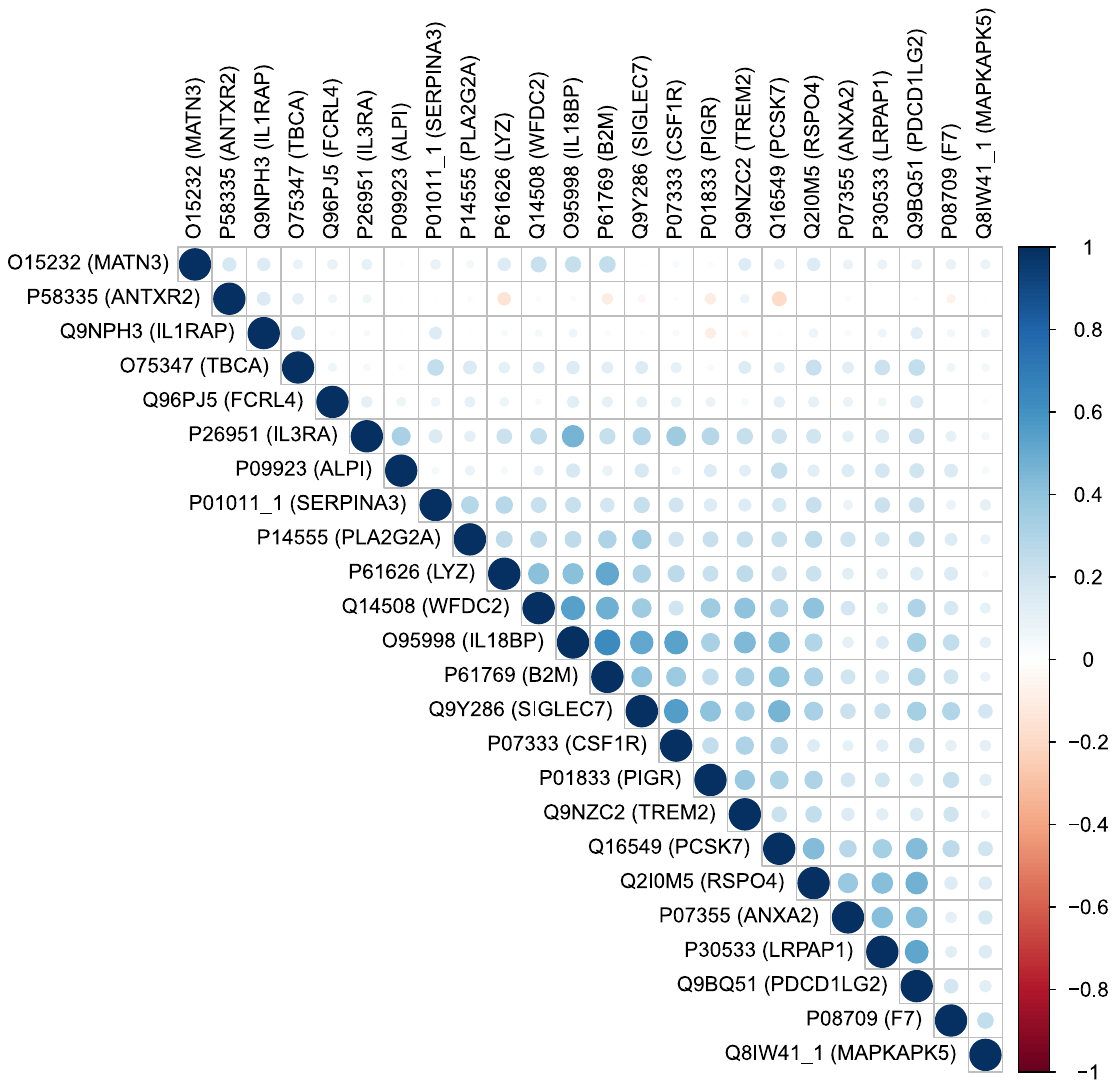

**Figure S4. Correlation structure between 24 proteins that had CpG site associations in BayesR+ (posterior inclusion probability > 95%).** The intercorrelations between proteins with significant CpG site associations in BayesR+ are shown. Hierarchical clustering was applied in order to determine sets of SOMAmer targets with strong correlation structure.

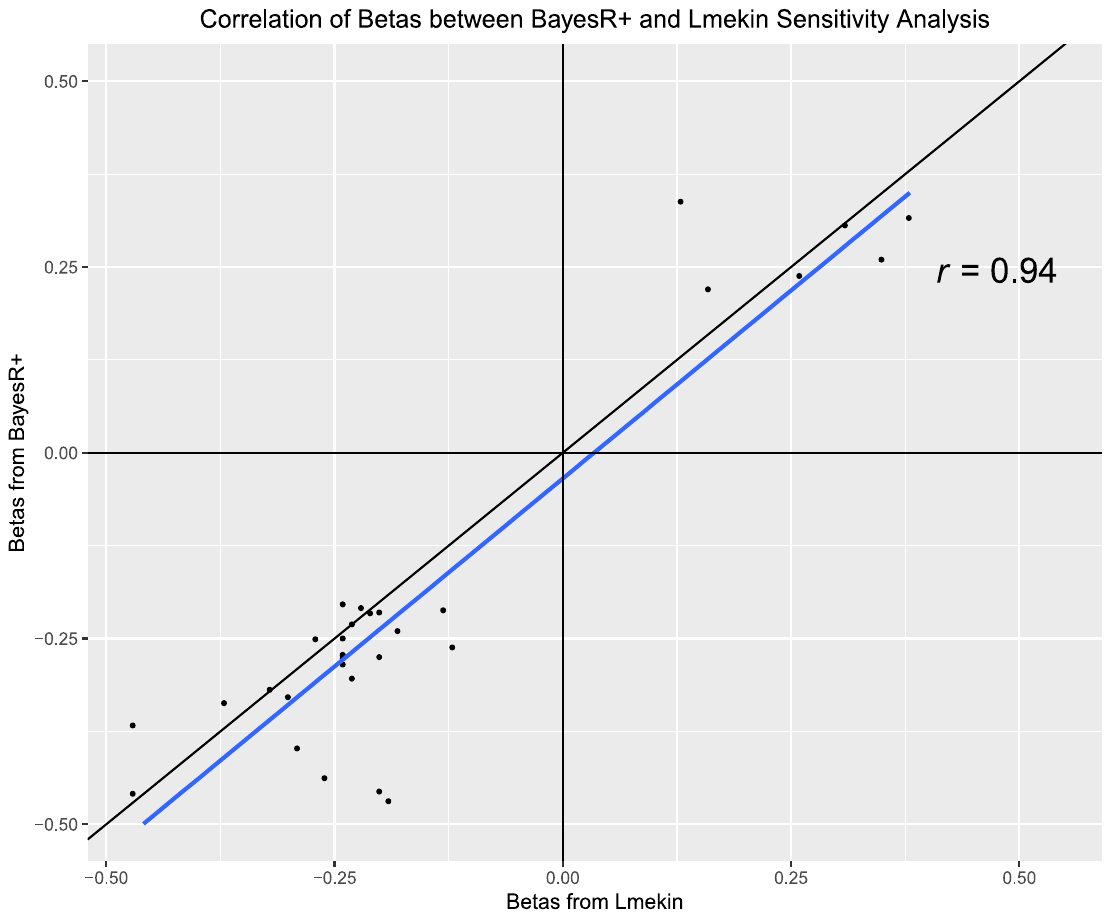

**Figure S5. Correlation between effect sizes from EWAS using BayesR+ and linear mixed effects models (lmekin).** Effect sizes for 32 CpG sites identified in the BayesR+ EWAS were correlated with corresponding effect sizes from linear mixed effects models that accounted for relatedness across samples (lmekin). Effect sizes were strongly correlated between BayesR+ and sensitivity EWAS using lmekin. EWAS, epigenome-wide association study.

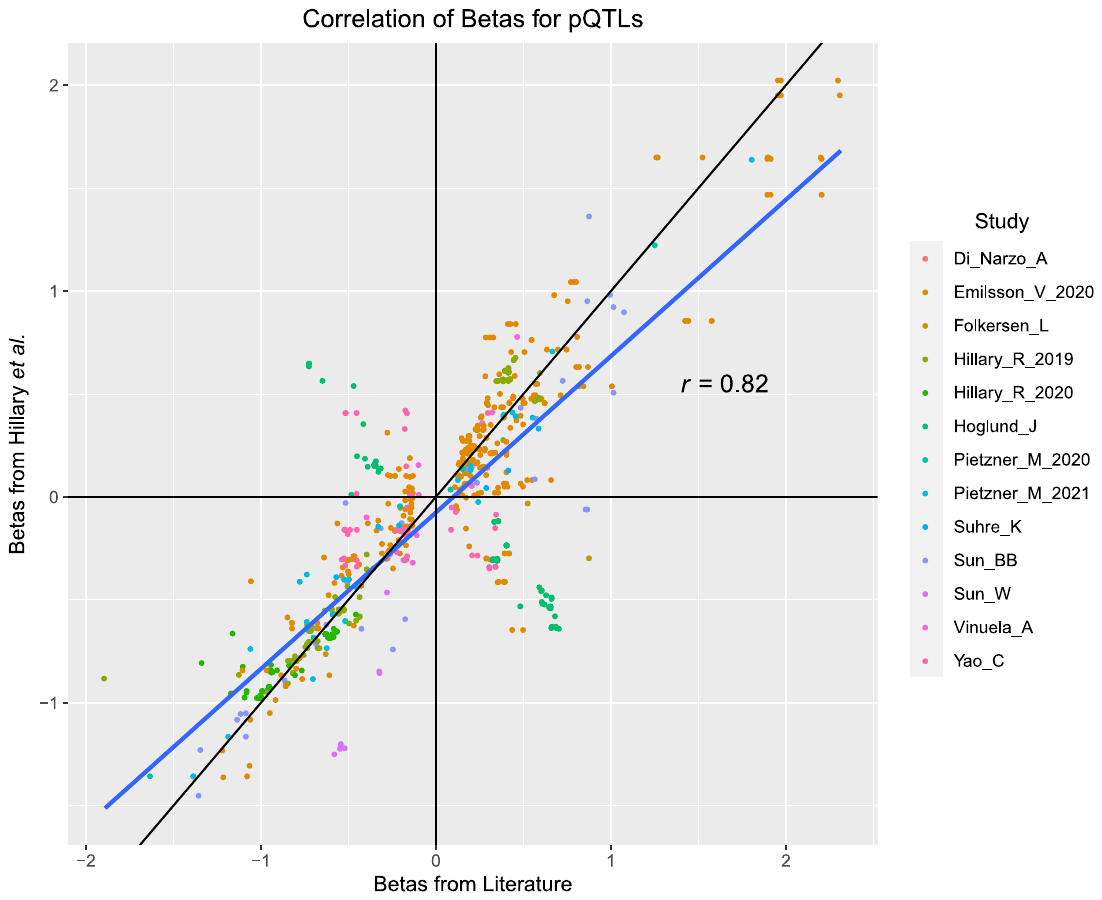

**Figure S6.** **Correlation between pQTL effect sizes in literature and those in present study.** All known pQTLs for the 40 unique proteins with pQTLs in our BayesR+ GWAS were extracted using Phenoscanner, GWAS Catalog and manual extraction of data from relevant studies. Of note, many of these pQTLs were non-significant in our study (n=796 comparisons). This strategy is separate from that in the main text. In the main text, we examined the correlation between our significant pQTLs and corresponding effect sizes in the literature. Here, we compared effect sizes for a much larger set of known significant pQTLs in the literature against effect sizes our dataset. Many of these known pQTLs were non-significant in our study. There was a strong correlation between effect sizes for known pQTLs and corresponding variants tested in our GWAS. GWAS, genome-wide association study; pQTL, protein quantitative trait locus.
