## Additional File 3 - Supplementary Methods for "Genome and epigenome wide studies of plasma protein biomarkers for Alzheimer’s disease implicate TBCA and TREM2 in disease risk"

**Mapping of SOMAmers from literature to those measured in Generation Scotland**

Multiple SOMAmers may recognise different epitopes on the same target protein. Consequently, 25 proteins identified in the literature had >1 available SOMAmer measured in Generation Scotland that could target the protein e.g. P01011 and P01011_1 map to the same target of SERPINA3. In total, 253 proteins had one SOMAmer, 18 had two SOMAmers, six had three SOMAmers and one (P02775, Platelet basic protein) had four SOMAmers that could target the protein in Generation Scotland. Therefore, 311 SOMAmers were available in the Generation Scotland dataset for the 359 proteins of interest in the literature. However, SOMAmers for P02671 (Fibrinogen alpha chain) and P02679 (Fibrinogen gamma chain) mapped to the larger fibrinogen protein product and therefore created duplicate entries. After removing duplicates, there were 309 unique SOMAmers available for analyses. These 309 SOMAmers targeted 281 proteins of interest (of the 359 identified in the literature).

**Search strategy and selection criteria**

We searched MEDLINE (Ovid interface, Ovid MEDLINE in-process and other non-indexed citations and Ovid MEDLINE 1946 onwards), Embase (Ovid interface, 1980 onwards), Web of Science (core collection, Thomson Reuters) and medRxiv/bioRxiv to identify relevant articles indexed as of 28 May 2021. We used the following search terms or their synonyms appropriate to each database: (“SOMAscan”.mp OR “Somalogic”.mp) AND (exp "plasma protein*" / OR exp "plasma proteomic" / OR exp “plasma levels” /) AND (“Alzheimer’s” OR *endophenotypes i.e. amyloid/tau/brain volume*).  Inclusion criteria were as follows: (i) original research article, (ii) proteins were measured in plasma, (iii) proteins were measured using SOMAscan® technology and (iv) proteins were associated with Alzheimer’s disease or related phenotypes. Twenty-five articles were identified in the systematic review, and one further manuscript was identified through a supplemental manual search of the literature. Of these 26 articles, 23 remained after removal of duplicate entries. Twelve met inclusion criteria. The other eleven articles were excluded for the following reasons: not related to Alzheimer’s disease (n=2), not measured in plasma (n=3), not an original research article (n=3) and not measured using SOMAscan® technology (n=3).

**Characterisation of *cis* and *trans* effects**

Genome-wide significant pQTLs and CpG sites were categorised into *cis* and *trans* effects. *Cis* effects were defined as sites that were located within 10 Mb of the transcription start site (TSS) of the gene encoding the target protein of interest. *Trans* effects were defined as those loci that lied outside of this region or were located on a chromosome distinct from the chromosome on which the target’s TSS is located. TSS positions were catalogued using *biomaRt* and Ensembl v83 (1, 2).

**Replication of existing pQTLs in the literature**

In a separate strategy to that described in the main text, we extracted all known pQTLs in the literature for the 40 proteins with pQTL associations in BayesR+. Following the extraction of known pQTLs, we assessed the correlation between their effect sizes and corresponding effect sizes of SNPs in our summary statistics. Of note, many of these extracted pQTLs were non-significant in our study. In this strategy, we observed a strong correlation between effect sizes for known genome-wide significant pQTLs and corresponding SNPs in our GWAS (n=796 comparisons, *r*=0.82, 95% CI = [0.79, 0.84], Additional File 2: Figure S6) (3-14).

**GARFIELD**

GARFIELD (**G**WAS **A**nalysis of **R**egulatory or **F**unctional **I**nformation **E**nrichment with **LD** correction), a non-parametric functional enrichment analysis, was used to test whether pQTLs were enriched for functional and regulatory characteristics (15). GARFIELD first performs ‘greedy pruning’ to identify independent variants among input SNPs using a LD threshold of *r*^2^≥0.01 and distance information. GARFIELD lists 1,005 features that are derived primarily from GENCODE (16), ENCODE (17) and ROADMAP projects (18). Variant functional annotation is performed if a variant, or a correlated variant (*r*^2^≥0.8), overlaps a regulatory annotation. Logistic regression is then performed to calculate odds ratios and enrichment *P* values for each annotation. Permutations are performed whilst accounting for confounding factors. These covariates are the number of LD proxies at *r*^2^≥0.8, minor allele frequency and distance to the nearest TSS. GARFIELD was performed for SNPs that passed a significance threshold of 1 x 10^-5^ and also those that passed a Bonferroni-corrected significance threshold of 1.62 x 10^-10^. Enrichments were deemed significant if they surpassed a significance threshold in GARFIELD that was corrected for the effective number of annotations (P<9.0 x 10^-5^).

Significant enrichments were present for pQTLs associated with ANXA2 and PLG levels. Protein QTLs associated with ANXA2 levels were enriched for chromatin peaks in brain tissue and DNase I hypersensitivity hotspots in blood. For PLG levels, pQTLs were enriched for variants that are associated with histone modifications in liver tissue (Additional File 1: Table S25).

**Pathway enrichment analyses using DNA methylation data**

Using methylation data, pathway enrichment was assessed among KEGG pathways and Gene Ontology (GO) terms with hypergeometric tests using the phyper function in R. All gene symbols from the EPIC array annotation (null set of sites) were converted to Entrez IDs using *biomaRt* (1, 2). GO terms and their corresponding gene sets were retrieved from the Molecular Signatures Database (MSigDB)-C5 (19). KEGG pathways were downloaded from the KEGG REST server (20).

**Cross-referencing of protein QTLs with existing methylation QTL databases**

Fifteen proteins exhibited both pQTL and CpG associations in BayesR+. There were 45 possible pQTL-CpG pairs across these proteins. We queried GoDMC and phenoscanner mQTL databases to identify whether any of these pairs were previously reported as mQTL-CpG associations (21, 22). However, only 29 pairs could be looked-up in these databases as these studies only examined probes present on the 450k array. We also tested all possible 45 SNP-CpG pairs, including those present on the EPIC array, in our own mQTL analyses. Methylation QTL analyses were performed using additive linear regression models and regressing CpG sites (beta values) on SNPs (0, 1, 2) while adjusting for age, sex, DNAm batch, set, Houseman-estimated white blood cell proportions and 20 genetic PCs (n=778).

Across GoDMC, phenoscanner and our mQTL data, 30 pairs represented mQTL effects at P < 5 x 10^-8^ (Additional File 1: Table S19). In instances where an mQTL effect was present in more than one database, summary statistics from the study with the largest sample size were used. *Coloc* also required the extraction of pQTL and mQTL summary statistics ±200 kb from the SNP of interest. Therefore, in instances where multiple SNPs within the same locus were mQTLs for the same CpG site, only the most significant mQTL effect in the locus was brought forward for colocalisation analyses (n=20 spread across 14 proteins).

**Quality control of proteomic data in Generation Scotland**

The following information outlines each stage of the quality control processes carried out within the 5k SOMAscan® v4 platform (23). Of the 5,284 reagents, 12 are spike-in controls, 286 are negative control/non-human targets, 7 are deprecated and 4,979 are human SOMAmers which target 4,776 unique protein targets. These 4,979 SOMAmers are spread across three dilution bits accordingly: 160 in the 0.005% bin, 797 in the 0.5% dilution group and 4,022 reagents are in the 20% bin. Across the 96-well plates, 11 wells are dedicated to replicate controls (5 calibrator samples, 3 quality control samples and 3 buffer or no protein samples) and 85 are reserved for biological samples.

- **Hybridisation control normalisation** is applied to control for nuisance variance within individual wells. A scaling factor is calculated as the median ratio of reference relative fluorescence intensities (RFUs) for 12 spike-ins against the observed RFUs in that sample or well. The reference RFUs are the median RFUs of these control SOMAmers across the entire plate of samples.
- **Intra-plate median signal normalisation** is performed to minimise variation across wells in a plate that might be caused by variability in pipetting, reagent concentration, washing steps, assay timing and differences in overall input protein concentration. This is applied separately to wells of the same class (i.e. separately for each buffer, calibrator, quality control type) and within SOMAmers of the same dilution factor (0.005%, 0.5%, and 20%). This creates a number of sample-SOMAmer groupings. The RFU of each SOMAmer (within a sample-SOMAmer group) is divided by the median of this SOMAmer’s RFUs across the entire plate. Then, a scale factor is applied to each well but only for SOMAmers in the SOMAmer-sample grouping. The scale factor associated with a given well is calculated as the inverse of the median ratio for that sample across all SOMAmers in the sample-SOMAmer grouping. In a given sample, RFUs for the SOMAmer in this grouping are median-normalised by multiplying RFUs by the scaling factor.
- **Calibration Normalisation** accounts for variability across plates within a run. This is typically caused by variability introduced by differences in scanner intensity. RFUs for dedicated calibrator samples in a plate are each divided by a reference value. The median of this ratio across calibrators in a plate is used to calculate a single scaling factor for the plate.
- **Calibration** refers to a normalisation procedure that accounts for variability between assay runs and/or experiments. This is performed on a SOMAmer-by-SOMAmer basis. Dedicated calibrator controls are utilised in this step. A SOMAmer-specific reference value is divided by the median of calibrator control RFUs and this gives the calibration scaling factor for the SOMAmer across the entire run.
- **Adaptive Normalisation by Maximum Likelihood** is an optional step which was performed in the Generation Scotland cohort that utilises estimates for the median signal and median absolute deviation of each SOMAmer taken from a reference sample (n ~ 1,000). This is performed separately for each dilution bin. This method provides a scaling factor for the SOMAmer that maximises the probability that a sample’s RFU comes from the sampling distribution. The method assumes that more than 30% of analytes are consistent with reference-based assumptions. Adaptive normalisation reduces technical variability between wells and inter-sample biological variability contributing to differences in total protein signal.
- **Post-calibration quality control** is carried out after the above steps. Three pooled quality control replicates are randomly distributed on the 96-well plate. For each SOMAmer, the accuracy of the median replicate signal on the plate is compared against a reference value. The result is a vector of quality control accuracy ratios across the SOMAmers. This provides information on whether there is still significant post-calibration variability and also on the quality of each assay run. In total, at least 85% of quality control ratios must be between 0.8 and 1.2 in a plate to meet acceptance criteria. Furthermore, a plate also has to show plate scaling factors between 0.4 and 2.5 prior to acceptance and release.

**Quality control of genotype data in Generation Scotland**

Quality control was performed to remove SNPs with a call rate of less than 98%, individuals with a genotyping rate of less than 98%, SNPs with a Hardy-Weinberg equilibrium *P*<1 x 10^-6^ and a minor allele frequency of greater than 0.01 or 1% (24). This left 561,125 tagged SNPs available for analyses (n=1,065 individuals). The --recode command in PLINK was used to convert genotype data in allele dosages (0, 1 and 2) (25). Missing data were mean imputed and allele dosages were scaled to mean zero and unit variance across 1,064 individuals who possessed both genotyping and SOMAscan® data. These steps were performed as BayesR+ cannot accept missing values and must have scaled markers as input.

**Quality control of DNA methylation data in Generation Scotland**

DNA methylation in the STRADL cohort (of Generation Scotland) was assayed into two separate sets (n_set1_=504, n_set2_=306). *Meffil* was used to exclude individuals who showed a mismatch between DNAm-predicted sex and recorded sex, samples in which more than 0.5% of CpGs had a detection *P*>0.01, outliers for bisulphite conversion control probes, samples with a median signal intensity >3 standard deviations lower than expected and samples showing evidence of dye bias (26). Following this, *shinyMethyl* was used to exclude outliers based on visual inspection of plots showing the log median intensity of methylated versus unmethylated signals per array (27). *Meffil* was used again to identify and exclude probes which had a beadcount of less than 3 in more than 5% of samples and/or probes in which >1% of samples had a detection *P*>0.01. Plots from multidimensional scaling (MDS) were investigated to inspect for further outlier samples. Forty male participants were identified as outliers according to X chromosome DNAm levels. These individuals were removed from analyses. Data were re-normalised and inspection of MDS plots confirmed that no further outliers were present. Data were normalised using the dasen method in *wateRmelon* (28) and converted to M values using the beta2m function in *lumi* (29). There were 778 individuals with methylation and proteomic data in STRADL.

In total, 793,706 CpG sites were retained after quality control in set 1 and 773,860 CpG sites remained in set 2. Additionally, 772,619 CpG sites were present in both sets. Therefore, both sets were truncated to these CpG sites. Within each set, DNAm levels were mean imputed as the BayesR+ software cannot accept missing values. Both sets were then combined and adjusted for DNAm batch, set, age and sex. Each CpG site was scaled to mean zero and unit variance.

**Cross-referencing pQTLs with publicly available eQTL or mQTL data**

*Cis* protein QTLs identified by BayesR+ were cross-referenced with FDR-corrected significant *cis* eQTL data from the eQTLGen consortium. In the eQTL dataset, 85% of samples were derived from whole blood and 15% of samples corresponded to peripheral blood mononuclear cell data (30). Protein QTLs were retained if they overlapped with *cis* eQTLs for the transcript that corresponded to the SOMAmer’s gene.

**Mendelian randomisation**

(i) *Cis* expression QTLs obtained from eQTLGen consortium were used as IV to test whether there was evidence for an association between differential gene expression levels and circulating protein levels (30). In addition, we tested whether plasma protein levels may have had an effect on gene expression levels. For this reverse test, *cis* pQTLs were as IV and the outcome variable was *cis* eQTL data for the same gene as reported by the eQTLGen consortium (30). Pruning was applied to genome-wide significant SNPs at *r*^2^<0.1.
(ii) We tested for causal relationships between DNA methylation levels at genes encoding SOMAmer targets and the plasma levels of corresponding protein. Methylation QTLs were used as IV with protein levels as the outcome. As described above, if an mQTL was present in more than one database (GoDMC, phenoscanner or our mQTL analyses (n=778)), coefficients from the study with the largest sample size were retained (21, 22). The Wald ratio test was used to test for associations between DNA methylation and protein levels. The converse was also tested. For this, we used pQTLs from BayesR+ to test for causal relationships between plasma protein levels and DNA methylation profiles.
(iii) Independent protein QTLs (LD *r*^2^<0.1) were used as instrumental variables (IV) to test for causal relationships between 42 SOMAmer levels and 20 AD-associated traits (31). These traits were family history of AD (31), brain and hippocampal volume (32, 33), rate of cognitive decline (34), 8 quantitative measures of CSF amyloid and tau levels in EMIF-AD (35), additional measures of CSF Aβ_42_, tau and phospho-tau_181_ in a larger meta-analysis (36), CSF clusterin (37), and CSF levels of APOE, proBNP, s100-beta, sortillin and YKL-40 (38). Two-sample MR was performed using MR-base. A stringent Bonferroni-corrected significance threshold of 5.95 x 10^-5^ (<0.05/840 tests = 20 traits x 42 SOMAmers) was applied (39). The converse relationship was also tested i.e. trait exhibiting a causal effect on protein levels. For this, genome-wide significant SNPs at 5 x 10^-8^ were extracted from GWA studies and pruned at *r*^2^<0.1. Pruned SNPs were used as IV to test for causal relationships with plasma protein levels as the outcome.

**References**

1.	Durinck S, Moreau Y, Kasprzyk A, Davis S, De Moor B, Brazma A, et al. BioMart and Bioconductor: a powerful link between biological databases and microarray data analysis. Bioinformatics (Oxford, England). 2005;21(16):3439-40.

2.	Durinck S, Spellman PT, Birney E, Huber W. Mapping identifiers for the integration of genomic datasets with the R/Bioconductor package biomaRt. Nature protocols. 2009;4(8):1184.

3.	Hillary RF, Trejo-Banos D, Kousathanas A, McCartney DL, Harris SE, Stevenson AJ, et al. Multi-method genome- and epigenome-wide studies of inflammatory protein levels in healthy older adults. Genome medicine. 2020;12(1):60.

4.	Di Narzo AF, Telesco SE, Brodmerkel C, Argmann C, Peters LA, Li K, et al. High-Throughput Characterization of Blood Serum Proteomics of IBD Patients with Respect to Aging and Genetic Factors. PLoS genetics. 2017;13(1):e1006565.

5.	Emilsson V, Ilkov M, Lamb JR, Finkel N, Gudmundsson EF, Pitts R, et al. Co-regulatory networks of human serum proteins link genetics to disease. Science (New York, NY). 2018;361(6404):769-73.

6.	Folkersen L, Fauman E, Sabater-Lleal M, Strawbridge RJ, Frånberg M, Sennblad B, et al. Mapping of 79 loci for 83 plasma protein biomarkers in cardiovascular disease. PLoS genetics. 2017;13(4):e1006706-e.

7.	Hillary RF, McCartney DL, Harris SE, Stevenson AJ, Seeboth A, Zhang Q, et al. Genome and epigenome wide studies of neurological protein biomarkers in the Lothian Birth Cohort 1936. Nature communications. 2019;10(1):3160.

8.	Hoglund J, Rafati N, Rask-Andersen M, Enroth S, Karlsson T, Ek WE, et al. Improved power and precision with whole genome sequencing data in genome-wide association studies of inflammatory biomarkers. Scientific reports. 2019;9(1):16844.

9.	Pietzner M, Wheeler E, Carrasco-Zanini J, Kerrison ND, Oerton E, Koprulu M, et al. Cross-platform proteomics to advance genetic prioritisation strategies. bioRxiv. 2021:2021.03.18.435919.

10.	Pietzner M, Wheeler E, Carrasco-Zanini J, Raffler J, Kerrison ND, Oerton E, et al. Genetic architecture of host proteins involved in SARS-CoV-2 infection. Nature communications. 2020;11(1):6397.

11.	Sun BB, Maranville JC, Peters JE, Stacey D, Staley JR, Blackshaw J, et al. Genomic atlas of the human plasma proteome. Nature. 2018;558(7708):73-9.

12.	Sun W, Kechris K, Jacobson S, Drummond MB, Hawkins GA, Yang J, et al. Common Genetic Polymorphisms Influence Blood Biomarker Measurements in COPD. PLoS genetics. 2016;12(8):e1006011-e.

13.	Yao C, Chen G, Song C, Keefe J, Mendelson M, Huan T, et al. Genome‐wide mapping of plasma protein QTLs identifies putatively causal genes and pathways for cardiovascular disease. Nature communications. 2018;9(1):3268.

14.	Viñuela A, Brown AA, Fernandez J, Hong M-G, Brorsson CA, Koivula RW, et al. Genetic analysis of blood molecular phenotypes reveals regulatory networks affecting complex traits: a DIRECT study. medRxiv. 2021:2021.03.26.21254347.

15.	Iotchkova V, Ritchie GRS, Geihs M, Morganella S, Min JL, Walter K, et al. GARFIELD classifies disease-relevant genomic features through integration of functional annotations with association signals. Nature genetics. 2019;51(2):343-53.

16.	Harrow J, Frankish A, Gonzalez JM, Tapanari E, Diekhans M, Kokocinski F, et al. GENCODE: the reference human genome annotation for The ENCODE Project. Genome Res. 2012;22(9):1760-74.

17.	Consortium EP. An integrated encyclopedia of DNA elements in the human genome. Nature. 2012;489(7414):57-74.

18.	Bernstein BE, Stamatoyannopoulos JA, Costello JF, Ren B, Milosavljevic A, Meissner A, et al. The NIH Roadmap Epigenomics Mapping Consortium. Nature biotechnology. 2010;28(10):1045-8.

19.	Liberzon A, Birger C, Thorvaldsdóttir H, Ghandi M, Mesirov JP, Tamayo P. The Molecular Signatures Database (MSigDB) hallmark gene set collection. Cell systems. 2015;1(6):417-25.

20.	Tenenbaum D. KEGGREST: Client-side REST access to KEGG. R package version. 2016;1(1).

21.	Min JL, Hemani G, Hannon E, Dekkers KF, Castillo-Fernandez J, Luijk R, et al. Genomic and phenomic insights from an atlas of genetic effects on DNA methylation. medRxiv. 2020:2020.09.01.20180406.

22.	Staley JR, Blackshaw J, Kamat MA, Ellis S, Surendran P, Sun BB, et al. PhenoScanner: a database of human genotype-phenotype associations. Bioinformatics (Oxford, England). 2016;32(20):3207-9.

23.	Candia J, Cheung F, Kotliarov Y, Fantoni G, Sellers B, Griesman T, et al. Assessment of Variability in the SOMAscan Assay. Scientific reports. 2017;7(1):14248.

24.	Smith BH, Campbell H, Blackwood D, Connell J, Connor M, Deary IJ, et al. Generation Scotland: the Scottish Family Health Study; a new resource for researching genes and heritability. BMC medical genetics. 2006;7(1):74.

25.	Purcell S, Neale B, Todd-Brown K, Thomas L, Ferreira MAR, Bender D, et al. PLINK: a tool set for whole-genome association and population-based linkage analyses. American journal of human genetics. 2007;81(3):559-75.

26.	Min JL, Hemani G, Davey Smith G, Relton C, Suderman M. Meffil: efficient normalization and analysis of very large DNA methylation datasets. Bioinformatics (Oxford, England). 2018;34(23):3983-9.

27.	Fortin JP, Fertig E, Hansen K. shinyMethyl: interactive quality control of Illumina 450k DNA methylation arrays in R. F1000Research. 2014;3:175.

28.	Pidsley R, Y Wong CC, Volta M, Lunnon K, Mill J, Schalkwyk LC. A data-driven approach to preprocessing Illumina 450K methylation array data. BMC genomics. 2013;14(1):293.

29.	Du P, Kibbe WA, Lin SM. lumi: a pipeline for processing Illumina microarray. Bioinformatics (Oxford, England). 2008;24(13):1547-8.

30.	Võsa U, Claringbould A, Westra H-J, Bonder MJ, Deelen P, Zeng B, et al. Unraveling the polygenic architecture of complex traits using blood eQTL metaanalysis. bioRxiv. 2018:447367.

31.	Jansen IE, Savage JE, Watanabe K, Bryois J, Williams DM, Steinberg S, et al. Genome-wide meta-analysis identifies new loci and functional pathways influencing Alzheimer’s disease risk. Nature genetics. 2019;51(3):404-13.

32.	Hibar DP, Stein JL, Renteria ME, Arias-Vasquez A, Desrivières S, Jahanshad N, et al. Common genetic variants influence human subcortical brain structures. Nature. 2015;520(7546):224-9.

33.	Elliott LT, Sharp K, Alfaro-Almagro F, Shi S, Miller KL, Douaud G, et al. Genome-wide association studies of brain imaging phenotypes in UK Biobank. Nature. 2018;562(7726):210-6.

34.	Sherva R, Tripodis Y, Bennett DA, Chibnik LB, Crane PK, de Jager PL, et al. Genome-wide association study of the rate of cognitive decline in Alzheimer's disease. Alzheimer's & dementia : the journal of the Alzheimer's Association. 2014;10(1):45-52.

35.	Hong S, Prokopenko D, Dobricic V, Kilpert F, Bos I, Vos SJB, et al. Genome-wide association study of Alzheimer’s disease CSF biomarkers in the EMIF-AD Multimodal Biomarker Discovery dataset. Translational psychiatry. 2020;10(1):403.

36.	Deming Y, Li Z, Kapoor M, Harari O, Del-Aguila JL, Black K, et al. Genome-wide association study identifies four novel loci associated with Alzheimer's endophenotypes and disease modifiers. Acta neuropathologica. 2017;133(5):839-56.

37.	Deming Y, Xia J, Cai Y, Lord J, Holmans P, Bertelsen S, et al. A potential endophenotype for Alzheimer's disease: cerebrospinal fluid clusterin. Neurobiol Aging. 2016;37:208.e1-.e9.

38.	Kauwe JS, Bailey MH, Ridge PG, Perry R, Wadsworth ME, Hoyt KL, et al. Genome-wide association study of CSF levels of 59 alzheimer's disease candidate proteins: significant associations with proteins involved in amyloid processing and inflammation. PLoS genetics. 2014;10(10):e1004758.

39.	Hemani G, Zheng J, Elsworth B, Wade KH, Haberland V, Baird D, et al. The MR-Base platform supports systematic causal inference across the human phenome. eLife. 2018;7.
