## Additional File 4 - Combined Variance Components BayesR for "Genome and epigenome wide studies of plasma protein biomarkers for Alzheimer’s disease implicate TBCA and TREM2 in disease risk"

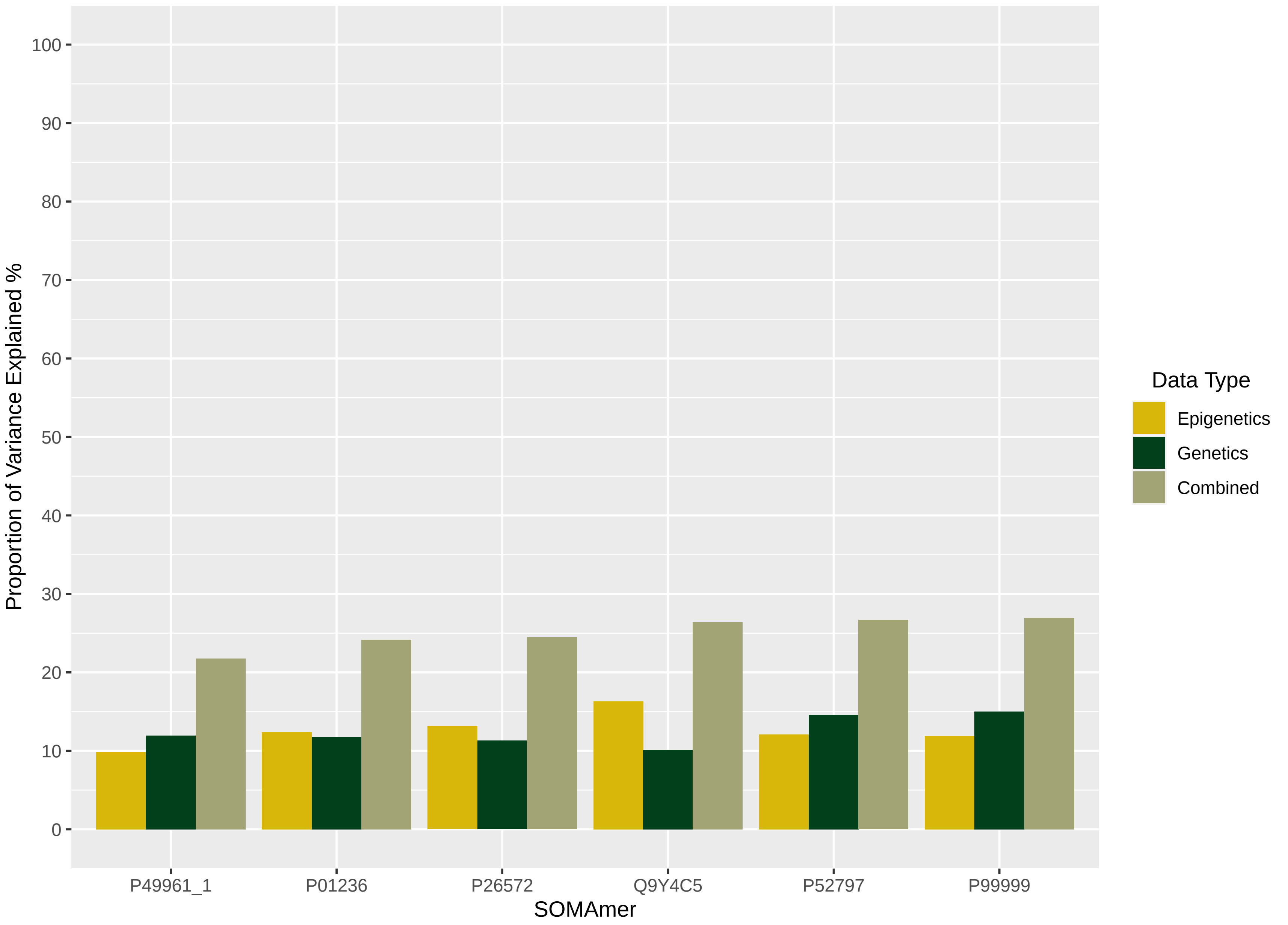

Proportion of Variance Explained %
100
90
80
70
60
50
40
30
20
10
0
P49961_1
P01236
P26572
Q9Y4C5
P52797
P99999
SOMAmer
Data Type
Epigenetics
Genetics
Combined

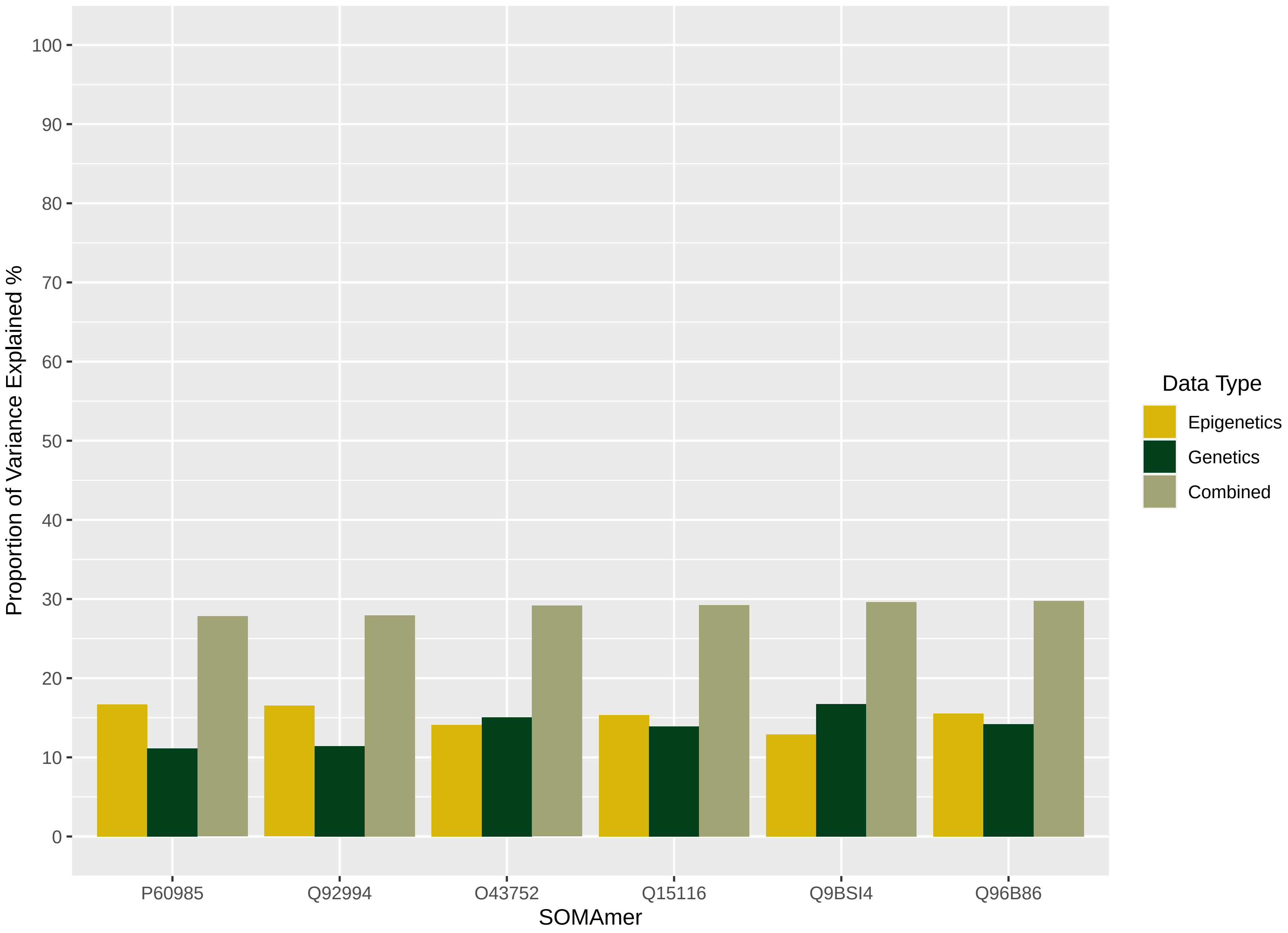

Proportion of Variance Explained %
100
90
80
70
60
50
40
30
20
10
0
P60985
Q92994
O43752
Q15116
Q9BSI4
Q96B86
SOMAmer
Data Type
Epigenetics
Genetics
Combined

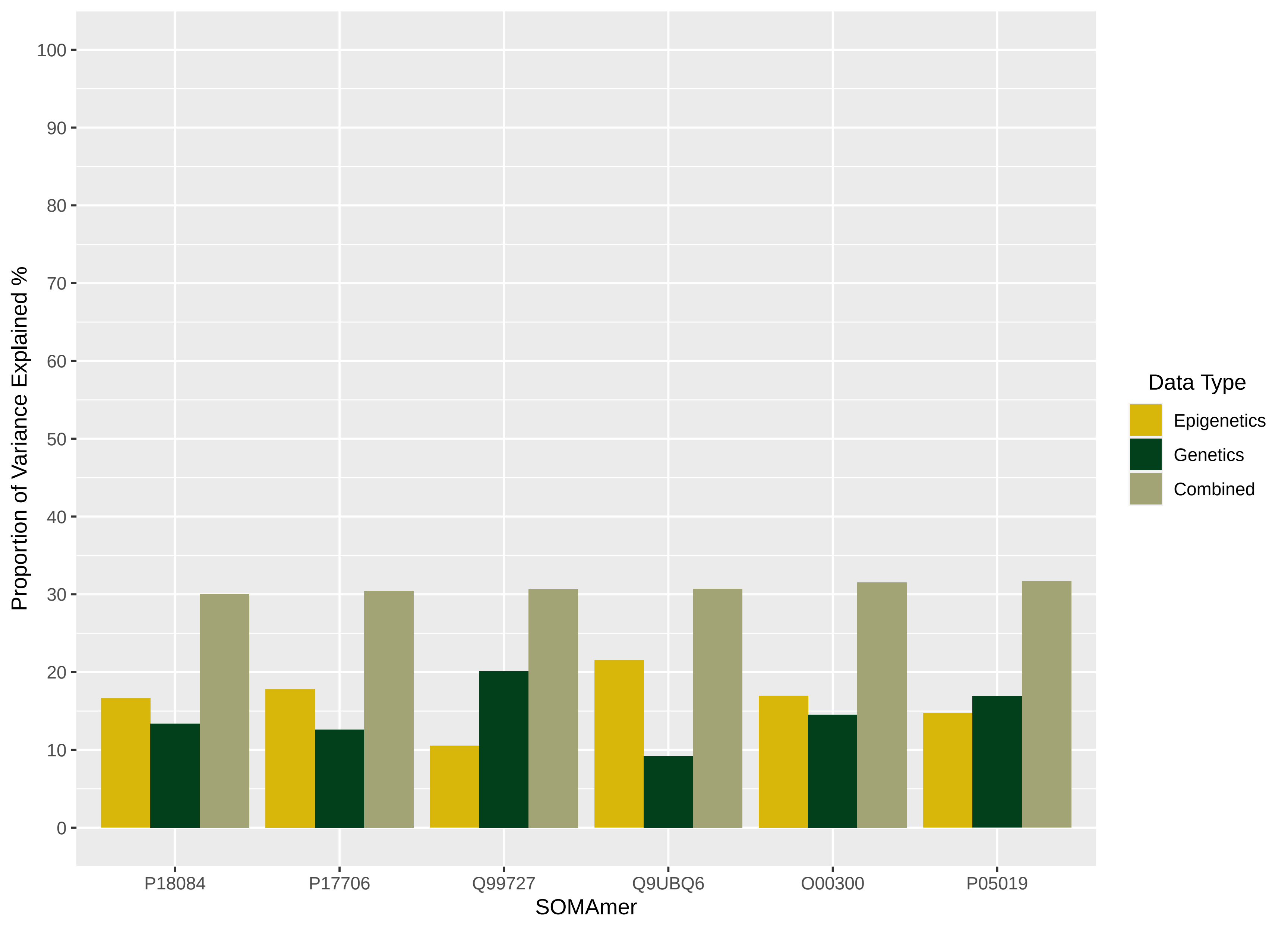

Proportion of Variance Explained %
0
10
20
30
40
50
60
70
80
90
100
P18084
P17706
Q99727
Q9UBQ6
O00300
P05019
SOMAmer
Data Type
Epigenetics
Genetics
Combined

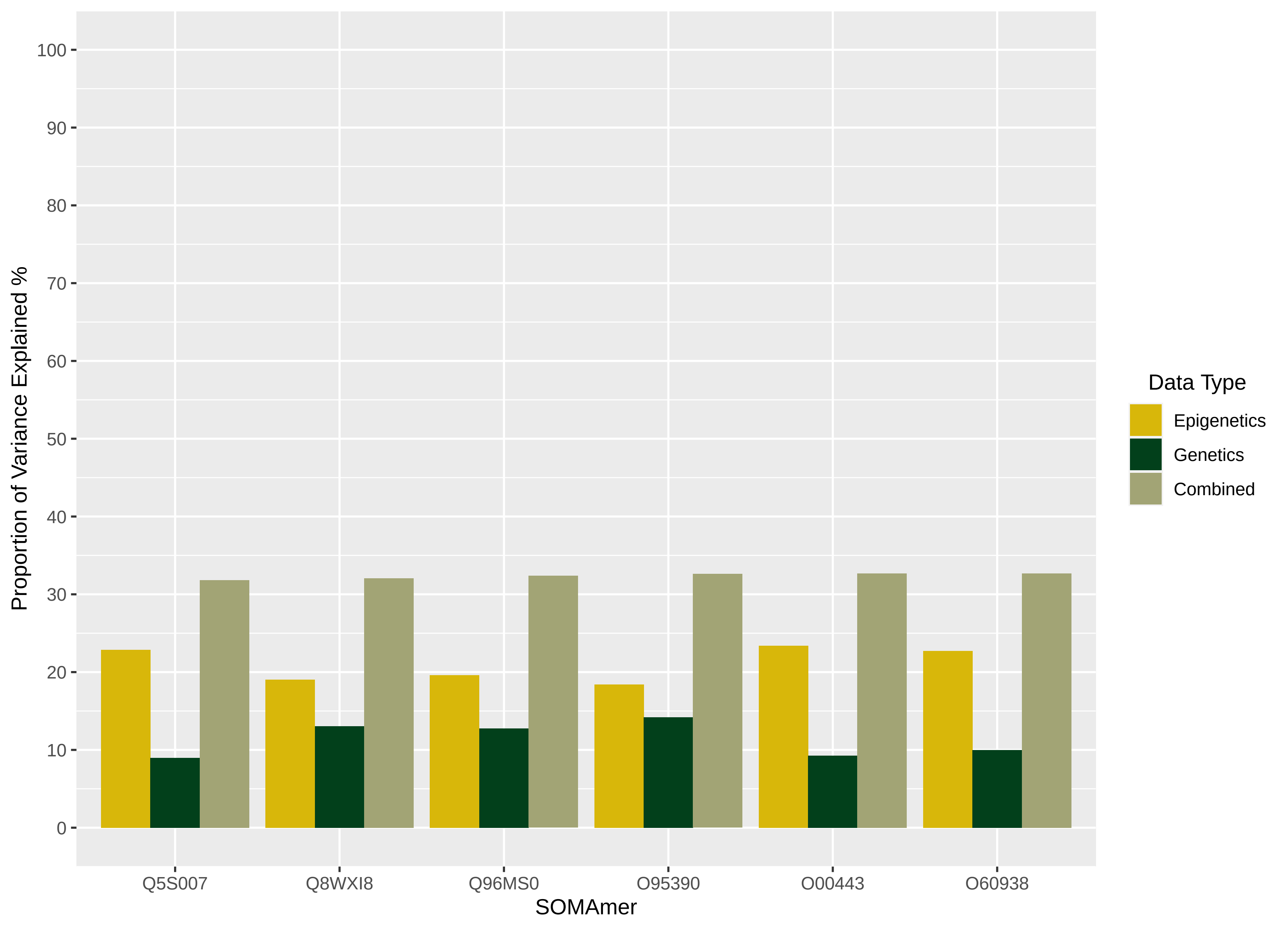
Proportion of Variance Explained %
100
90
80
70
60
50
40
30
20
10
0
Q5S007
Q8WXI8
Q96MS0
O95390
O00443
O60938
SOMAmer
Data Type
Epigenetics
Genetics
Combined

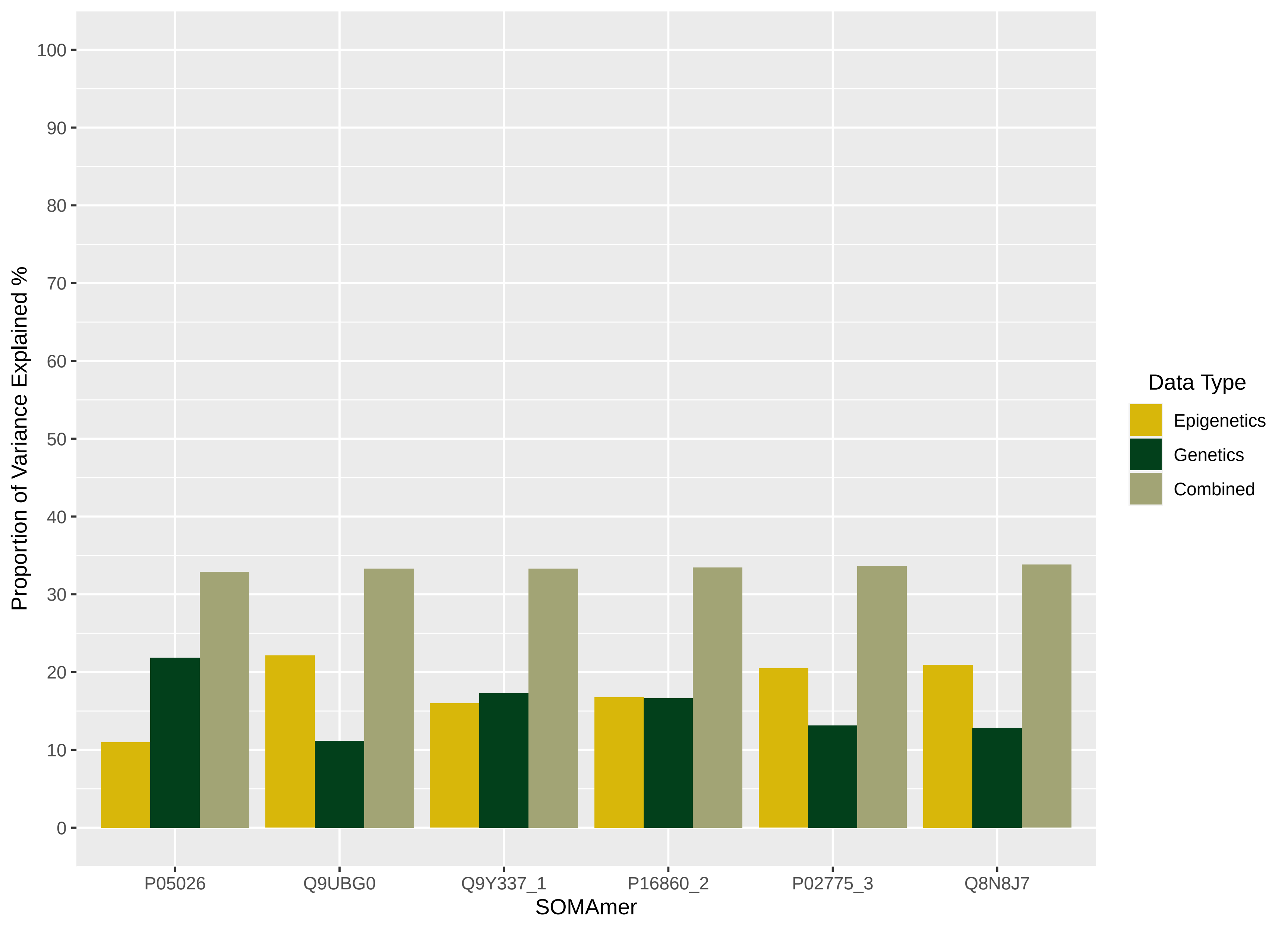

Proportion of Variance Explained %
100
90
80
70
60
50
40
30
20
10
0
P05026
Q9UBG0
Q9Y337_1
P16860_2
P02775_3
Q8N8J7
SOMAmer
Data Type
Epigenetics
Genetics
Combined

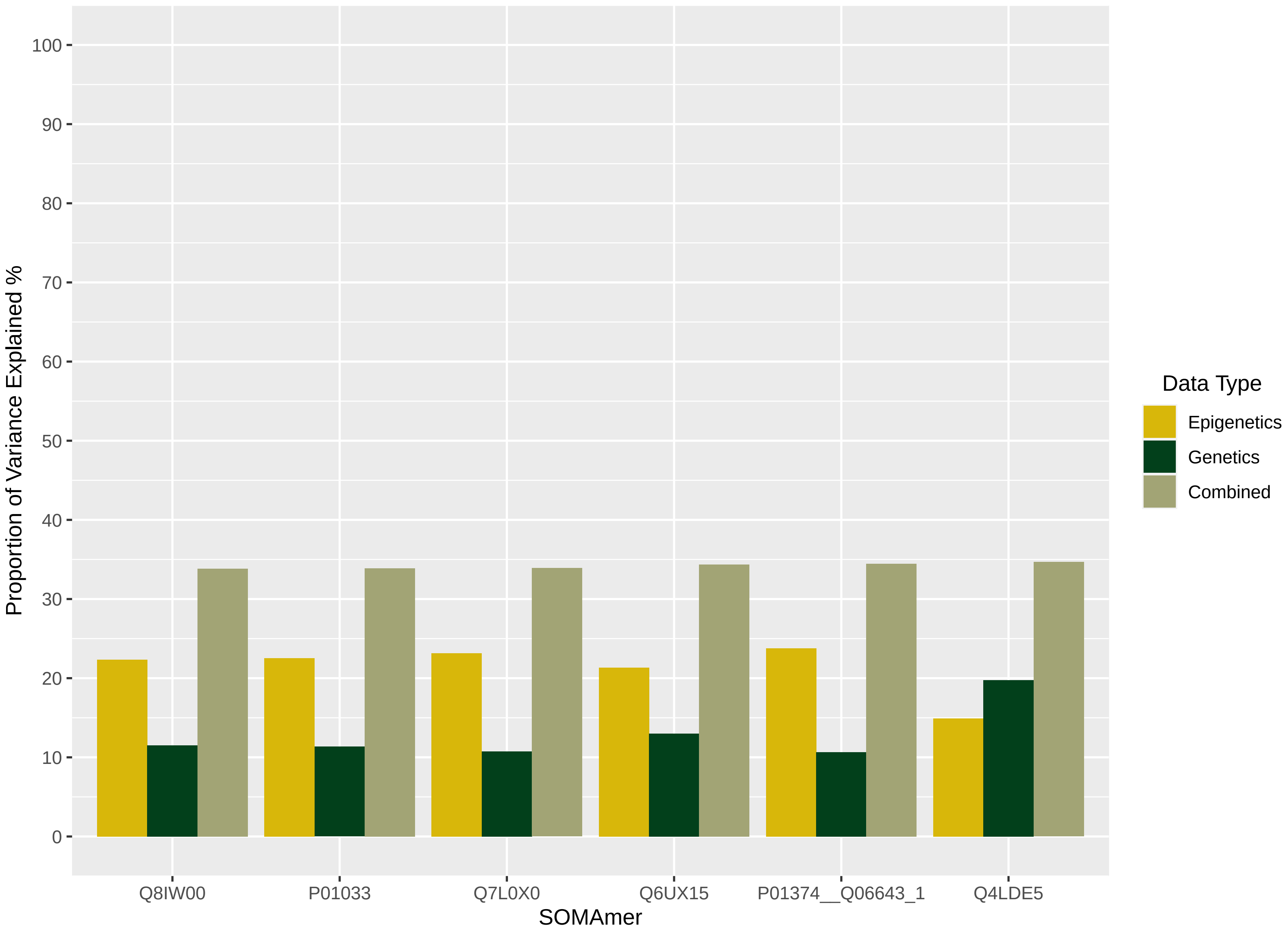

Proportion of Variance Explained %
0
10
20
30
40
50
60
70
80
90
100
Q8IW00
P01033
Q7L0X0
Q6UX15
P01374__Q06643_1
Q4LDE5
SOMAmer
Data Type
Epigenetics
Genetics
Combined

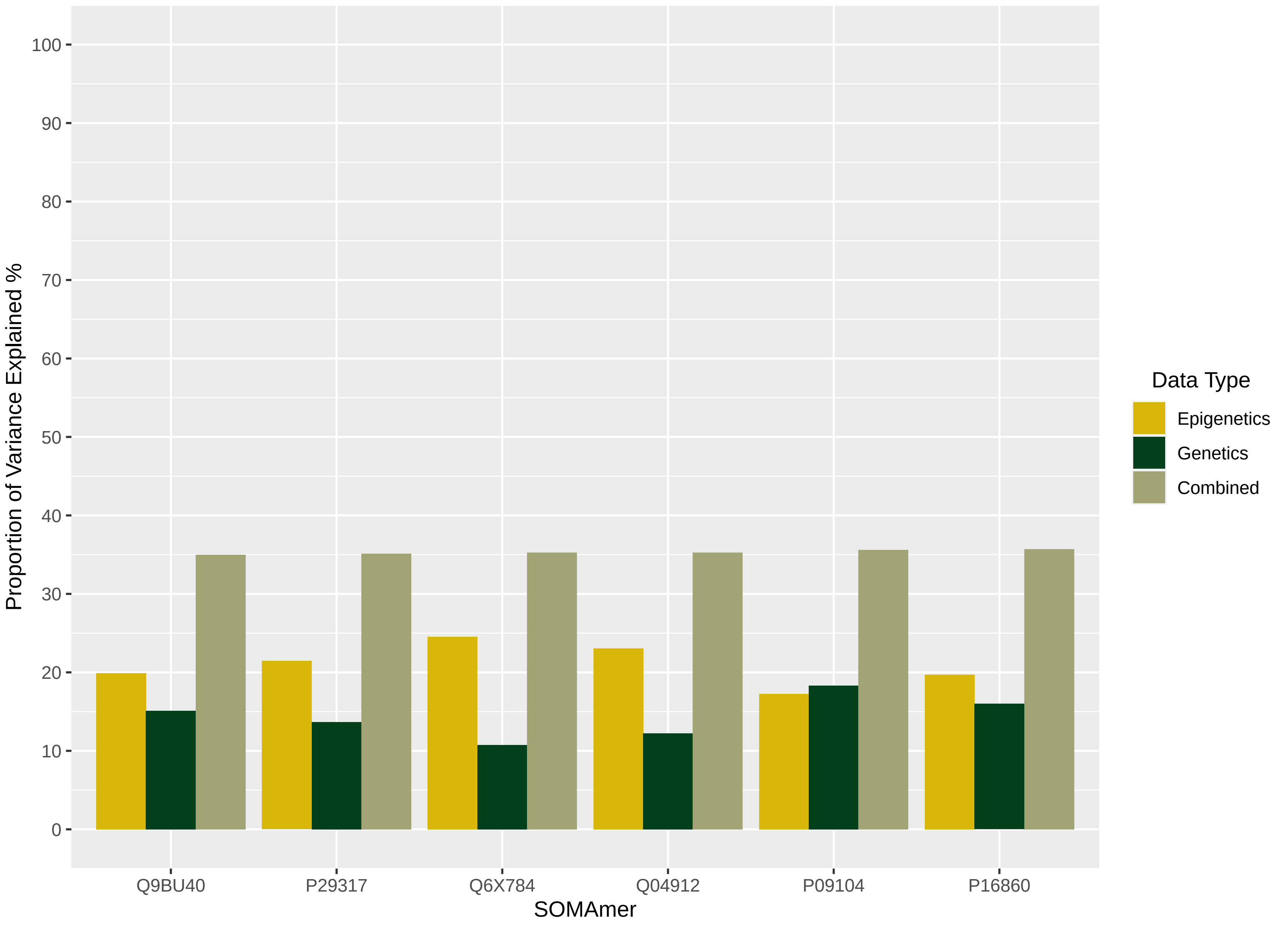

Proportion of Variance Explained %
100
90
80
70
60
50
40
30
20
10
0
Q9BU40
P29317
Q6X784
Q04912
P09104
P16860
SOMAmer
Data Type
Epigenetics
Genetics
Combined

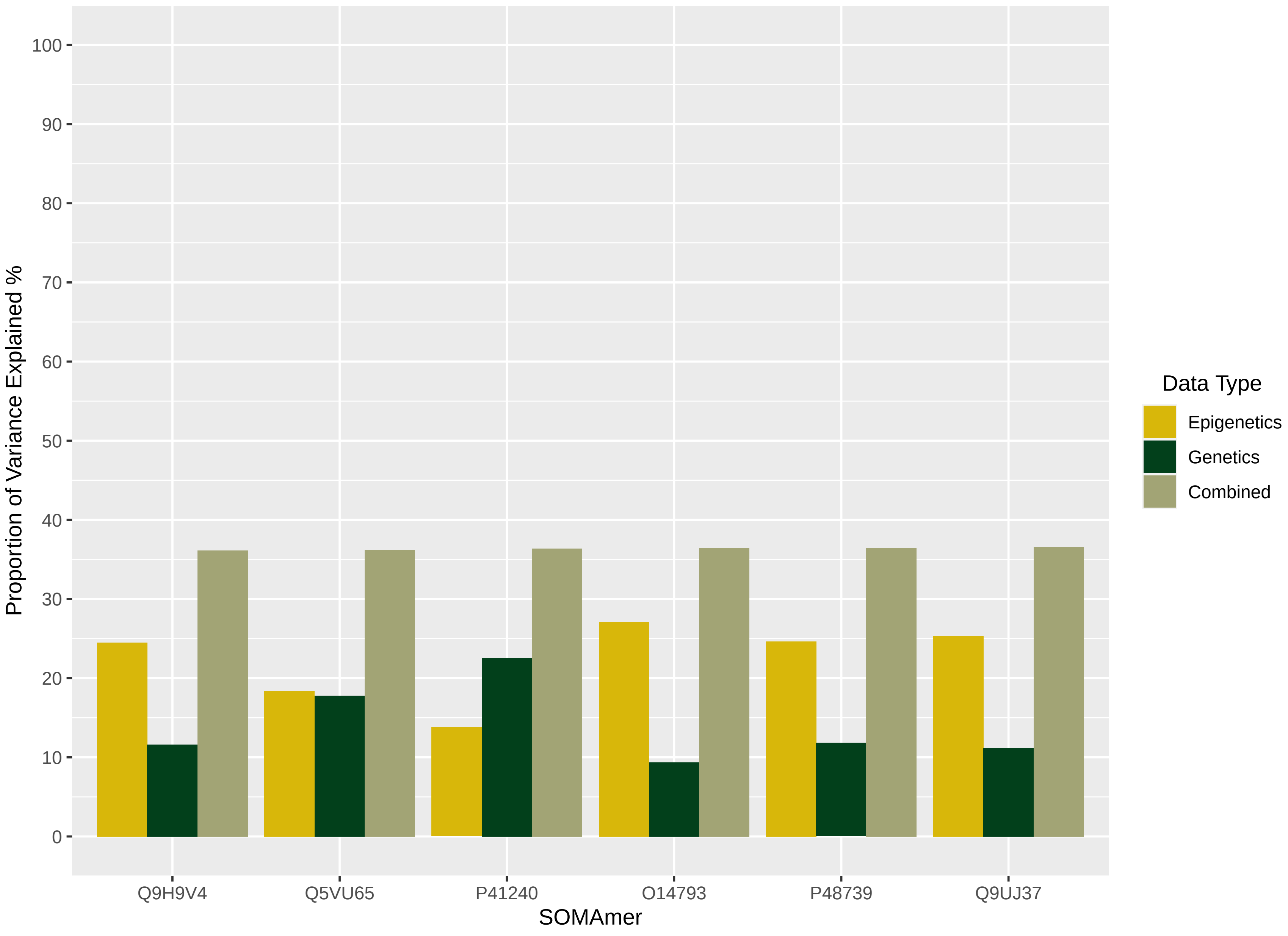

Proportion of Variance Explained %
100
90
80
70
60
50
40
30
20
10
0
Q9H9V4
Q5VU65
P41240
O14793
P48739
Q9UJ37
SOMAmer
Data Type
Epigenetics
Genetics
Combined

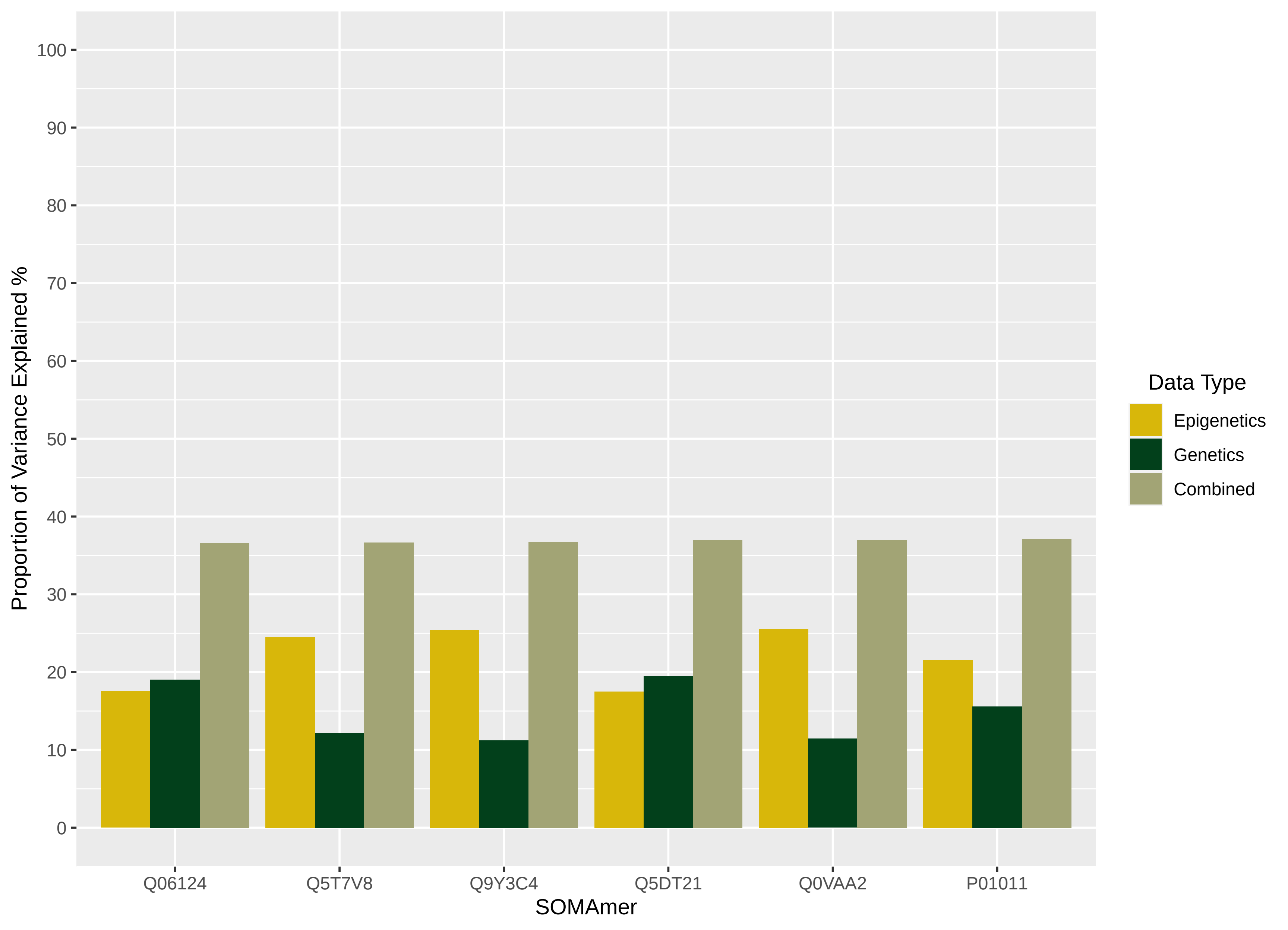

Proportion of Variance Explained %
100
90
80
70
60
50
40
30
20
10
0
Q06124
Q5T7V8
Q9Y3C4
Q5DT21
Q0VAA2
P01011
SOMAmer
Data Type
Epigenetics
Genetics
Combined

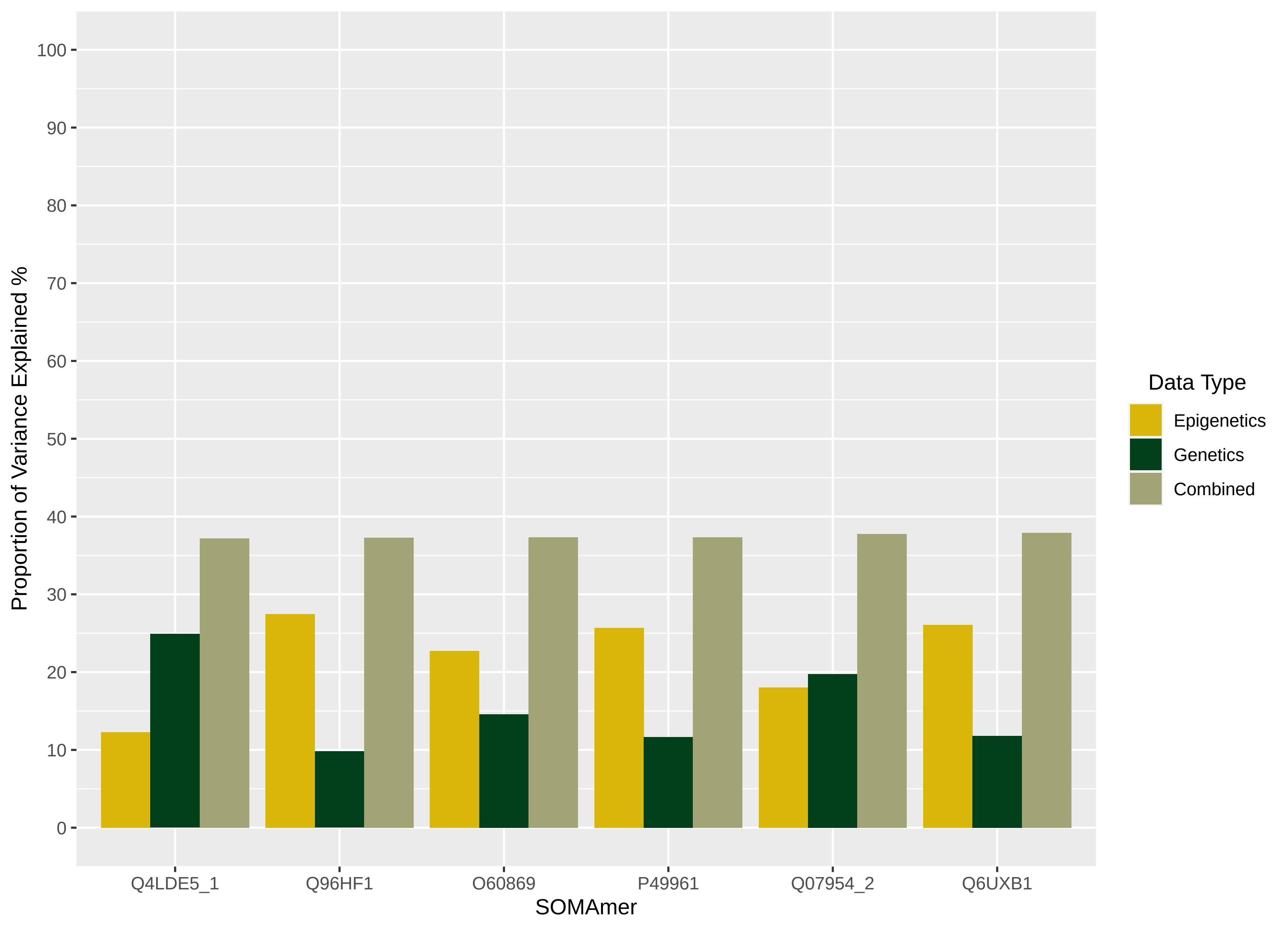
Proportion of Variance Explained %
0
10
20
30
40
50
60
70
80
90
100
Q4LDE5_1
Q96HF1
O60869
P49961
Q07954_2
Q6UXB1
SOMAmer
Data Type
Epigenetics
Genetics
Combined

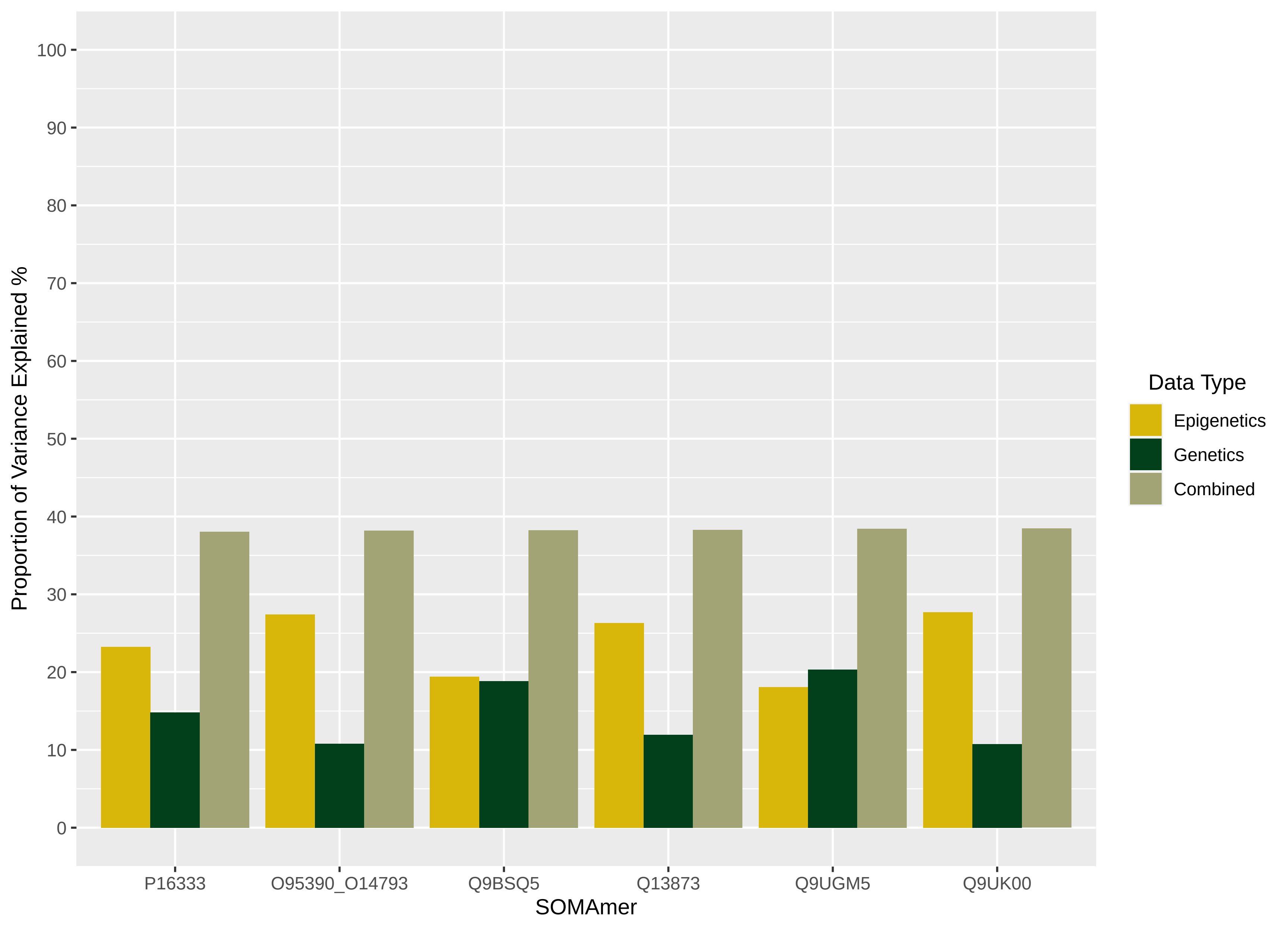

Proportion of Variance Explained %
100
90
80
70
60
50
40
30
20
10
0
P16333
O95390_O14793
Q9BSQ5
Q13873
Q9UGM5
Q9UK00
SOMAmer
Data Type
Epigenetics
Genetics
Combined

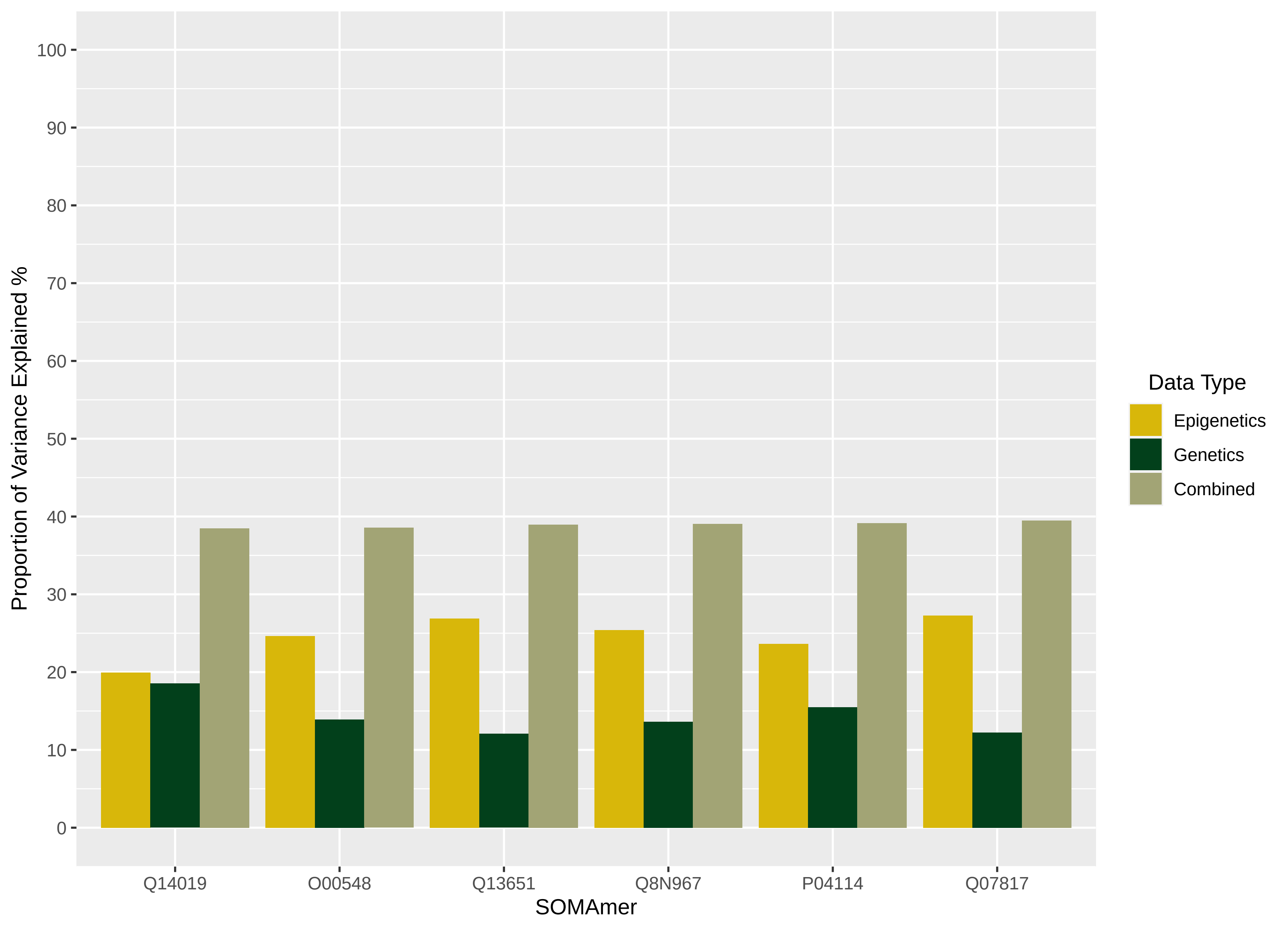

Proportion of Variance Explained %
100
90
80
70
60
50
40
30
20
10
0
Q14019
O00548
Q13651
Q8N967
P04114
Q07817
SOMAmer
Data Type
Epigenetics
Genetics
Combined

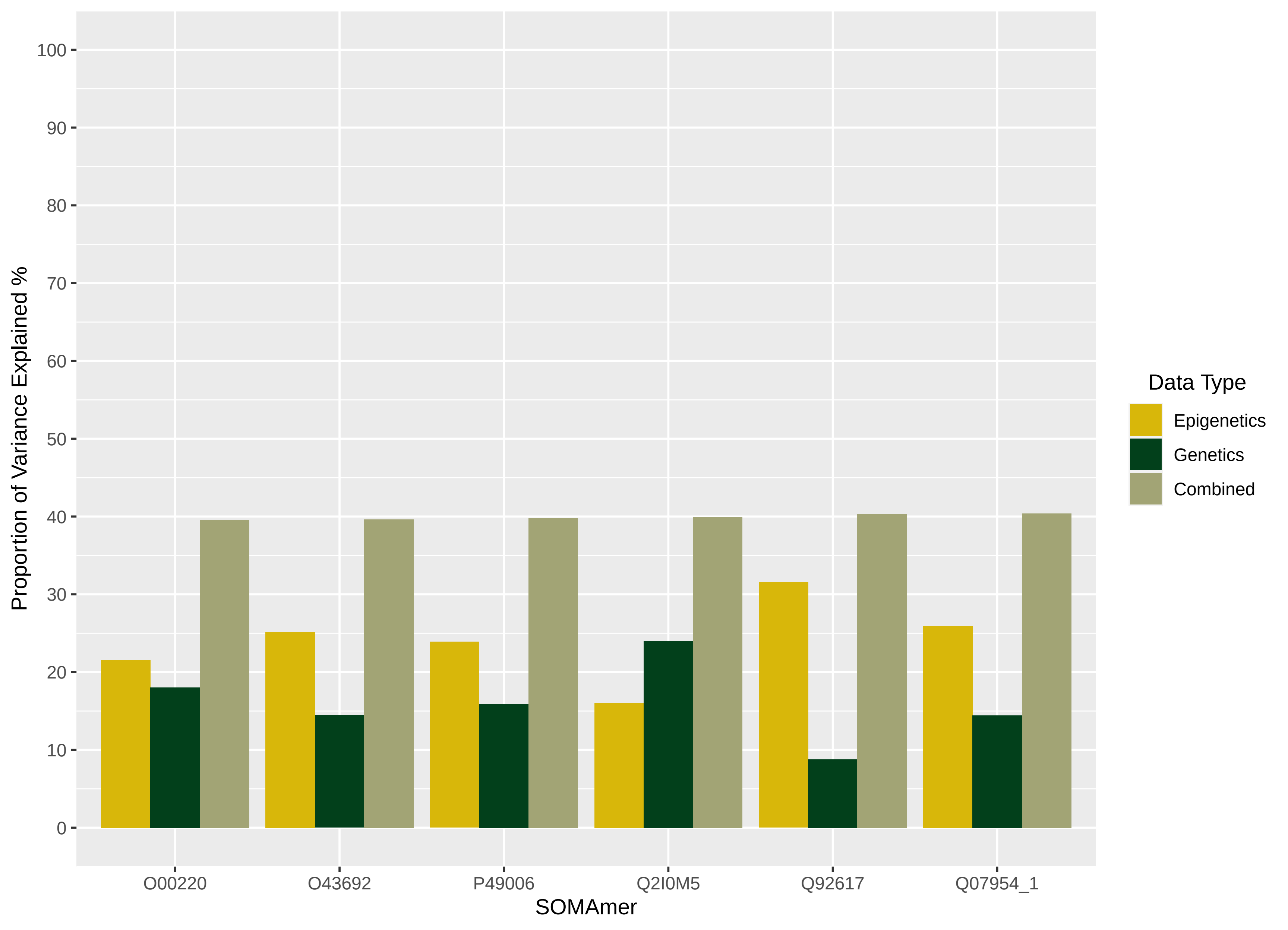
Proportion of Variance Explained %
100
90
80
70
60
50
40
30
20
10
0
O00220
O43692
P49006
Q2I0M5
Q92617
Q07954_1
SOMAmer
Data Type
Epigenetics
Genetics
Combined

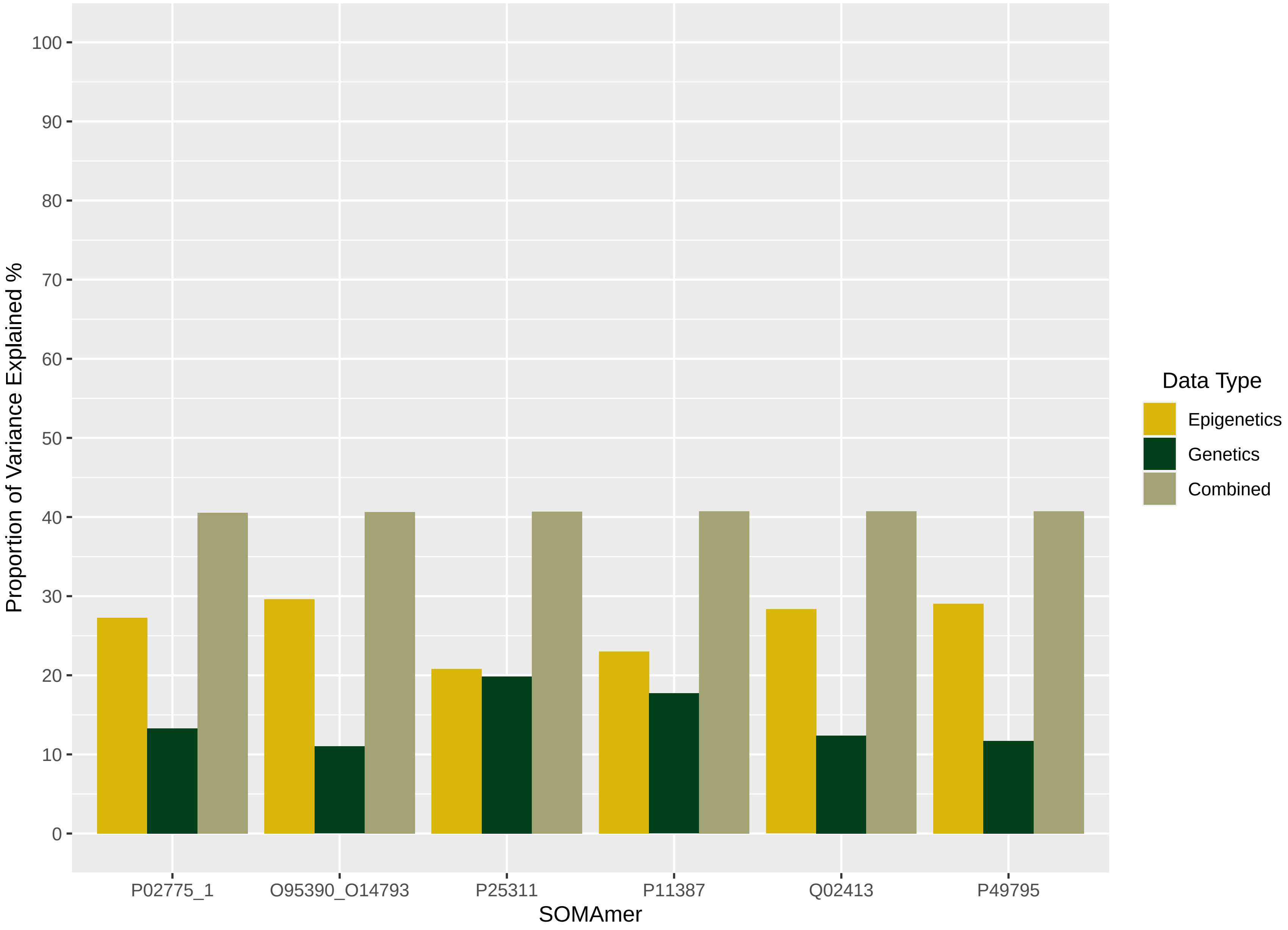

Proportion of Variance Explained %
100
90
80
70
60
50
40
30
20
10
0
P02775_1
O95390_O14793
P25311
P11387
Q02413
P49795
SOMAmer
Data Type
Epigenetics
Genetics
Combined

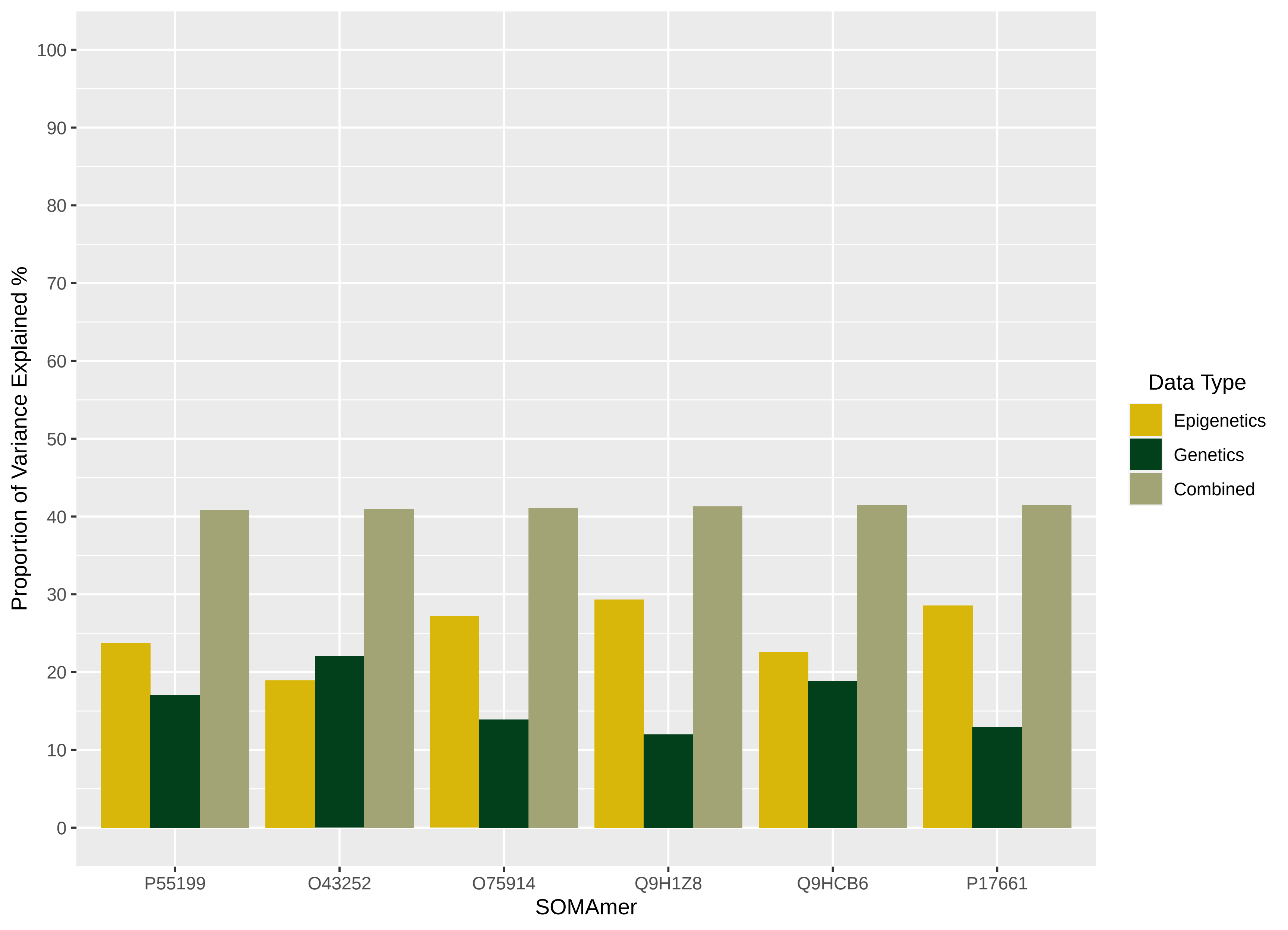

Proportion of Variance Explained %
100
90
80
70
60
50
40
30
20
10
0
P55199
O43252
O75914
Q9H1Z8
Q9HCB6
P17661
SOMAmer
Data Type
Epigenetics
Genetics
Combined

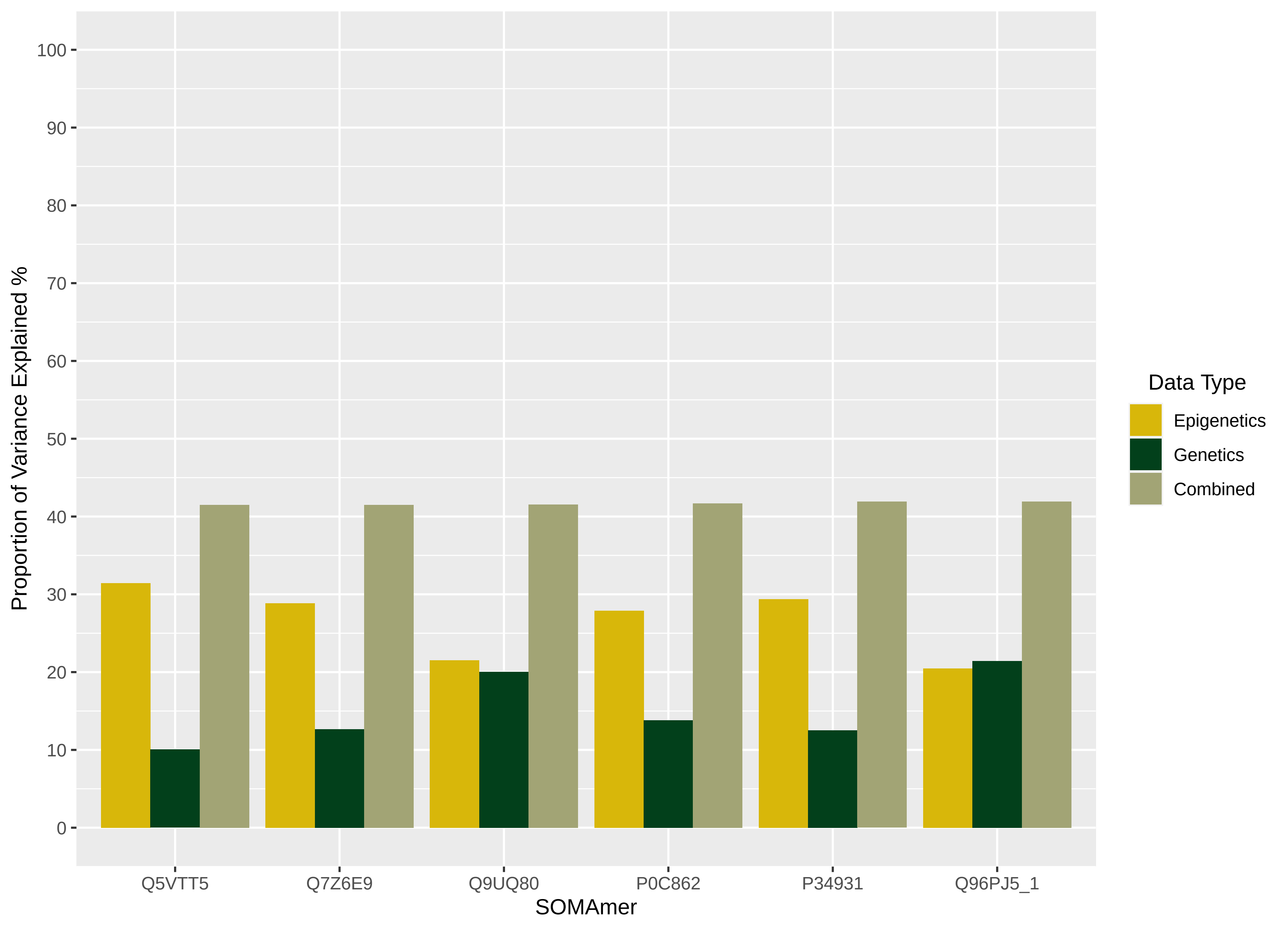

Proportion of Variance Explained %
100
90
80
70
60
50
40
30
20
10
0
Q5VTT5
Q7Z6E9
Q9UQ80
P0C862
P34931
Q96PJ5_1
SOMAmer
Data Type
Epigenetics
Genetics
Combined

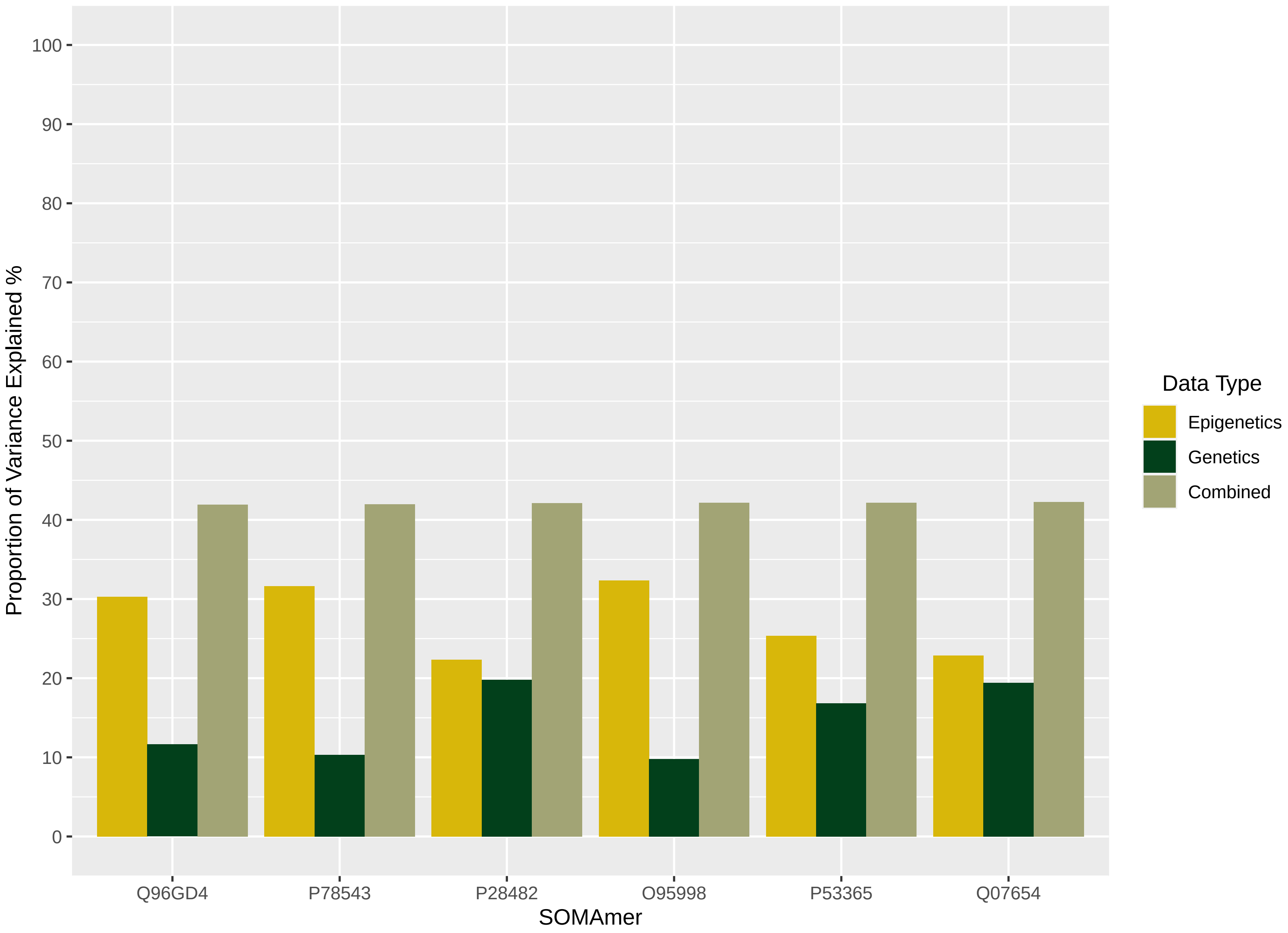
Proportion of Variance Explained %
100
90
80
70
60
50
40
30
20
10
0
Q96GD4
P78543
P28482
O95998
P53365
Q07654
SOMAmer
Data Type
Epigenetics
Genetics
Combined

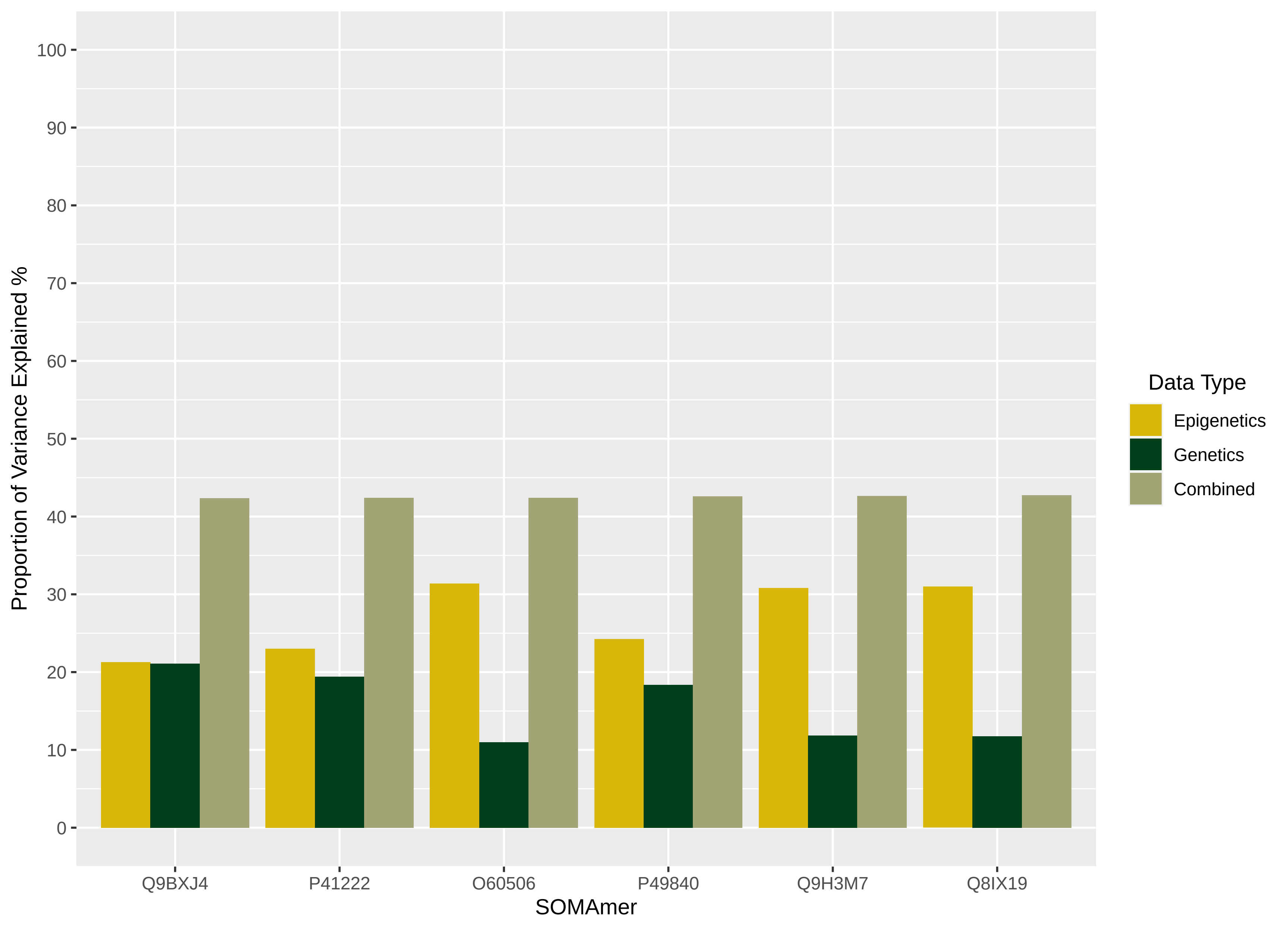
Proportion of Variance Explained %
100
90
80
70
60
50
40
30
20
10
0
Q9BXJ4
P41222
O60506
P49840
Q9H3M7
Q8IX19
SOMAmer
Data Type
Epigenetics
Genetics
Combined

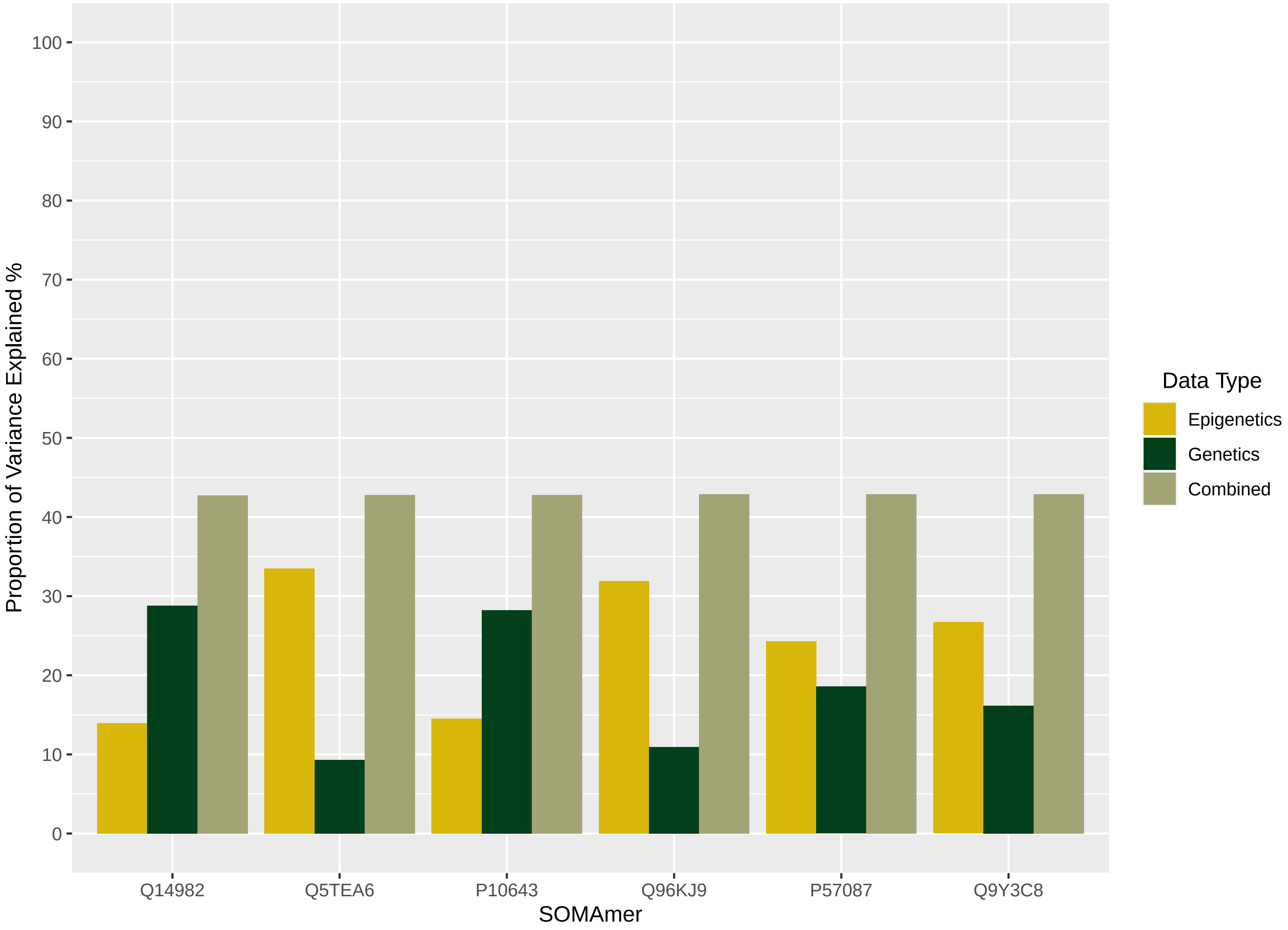
Proportion of Variance Explained %
100
90
80
70
60
50
40
30
20
10
0
Q14982
Q5TEA6
P10643
Q96KJ9
P57087
Q9Y3C8
SOMAmer
Data Type
Epigenetics
Genetics
Combined

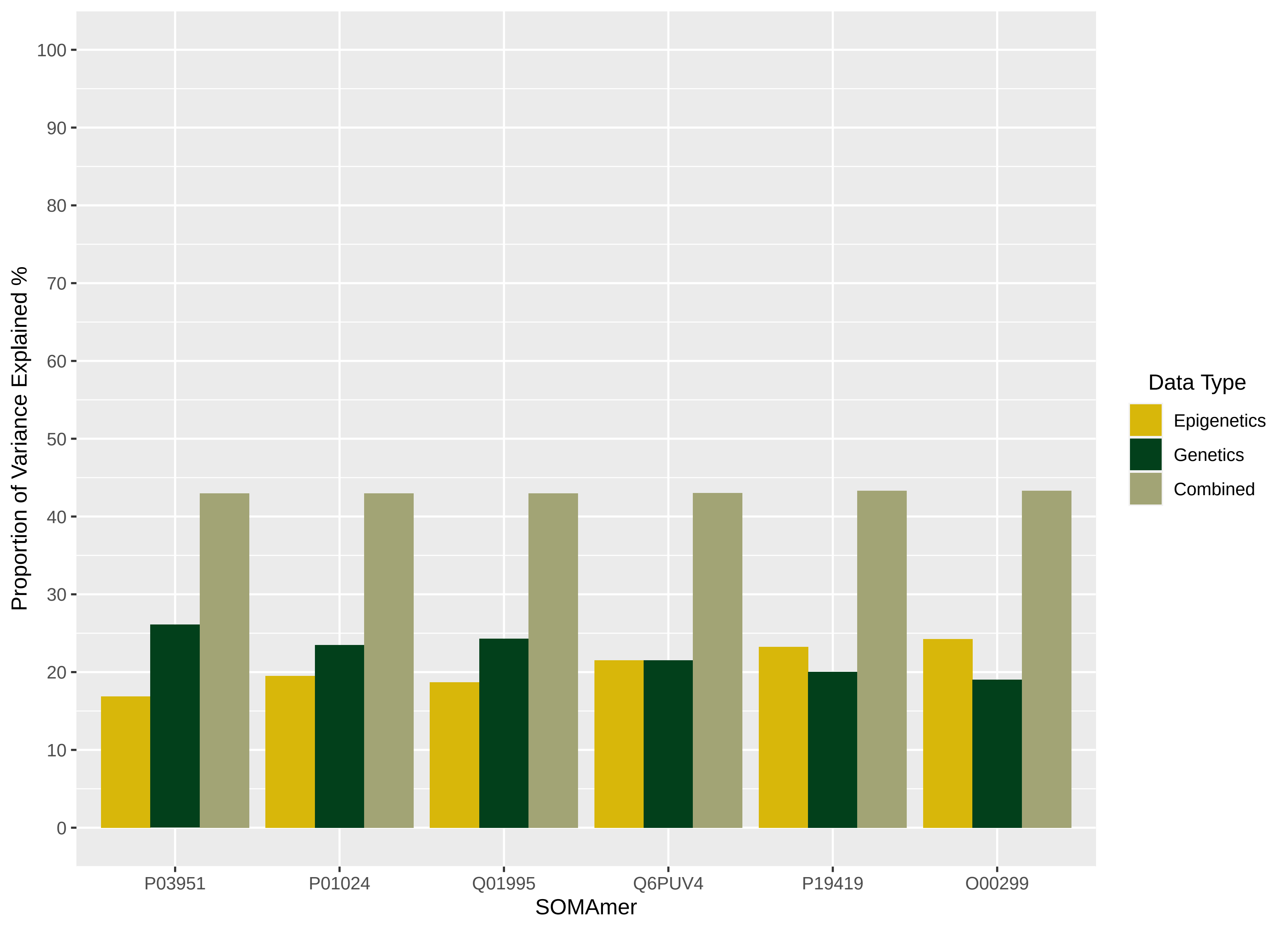

Proportion of Variance Explained %
100
90
80
70
60
50
40
30
20
10
0
P03951
P01024
Q01995
Q6PUV4
P19419
O00299
SOMAmer
Data Type
Epigenetics
Genetics
Combined

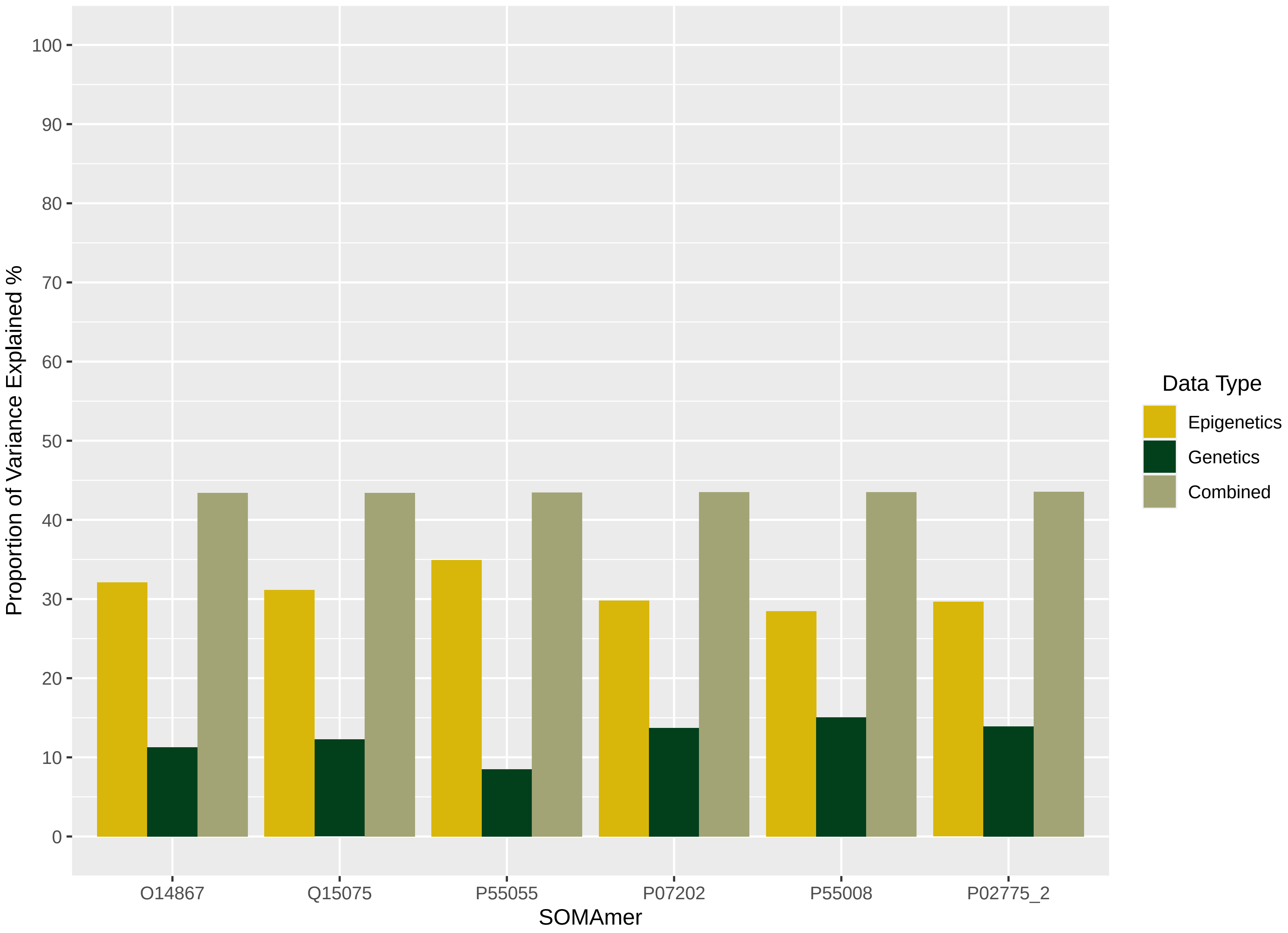

100
90
80
70
60
50
40
30
20
10
0
Proportion of Variance Explained %
O14867
Q15075
P55055
P07202
P55008
P02775_2
SOMAmer
Data Type
Epigenetics
Genetics
Combined

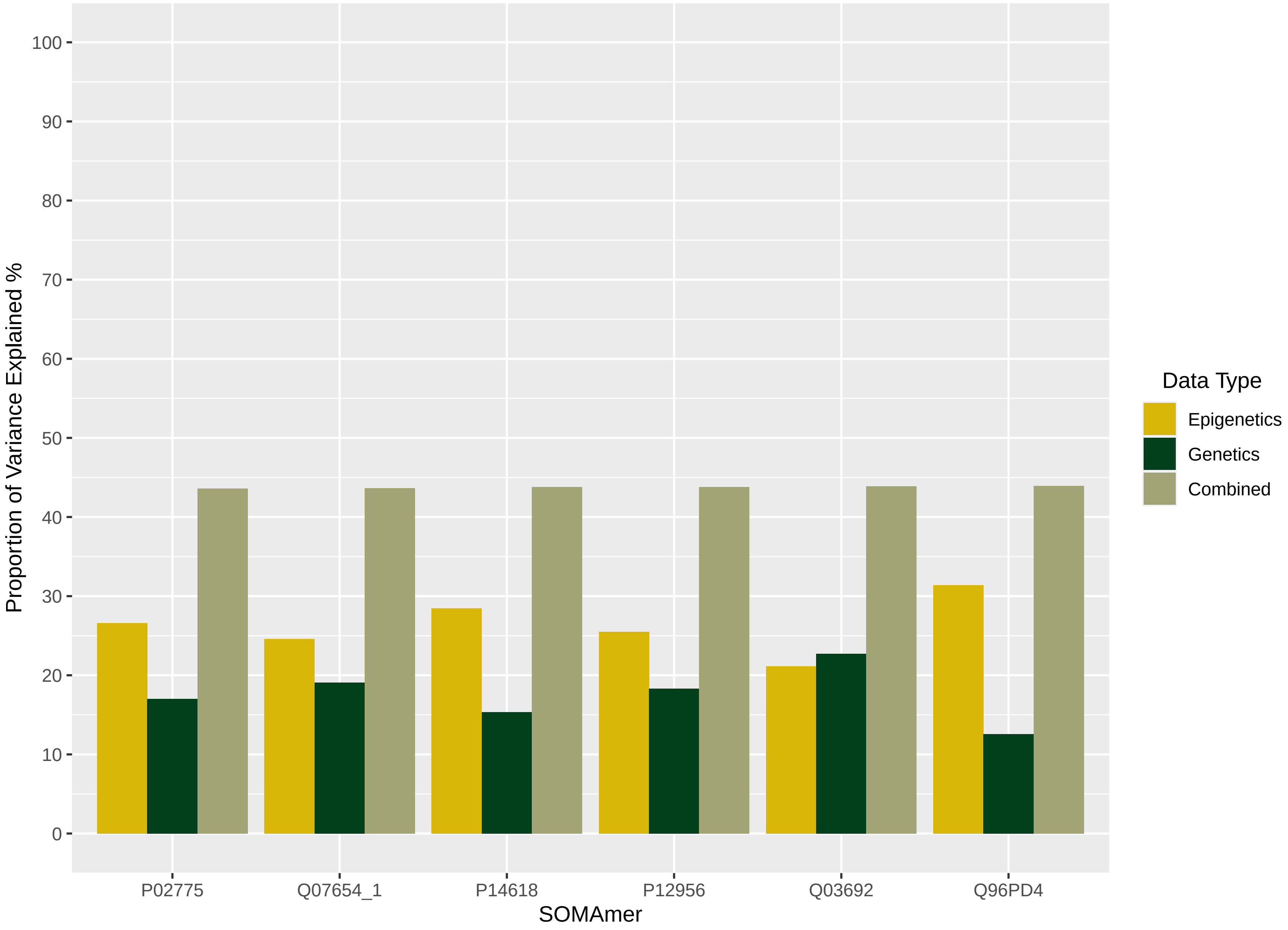

Data Type
Epigenetics
Genetics
Combined
Proportion of Variance Explained %
SOMAmer
P02775
Q07654_1
P14618
P12956
Q03692
Q96PD4
0
10
20
30
40
50
60
70
80
90
100

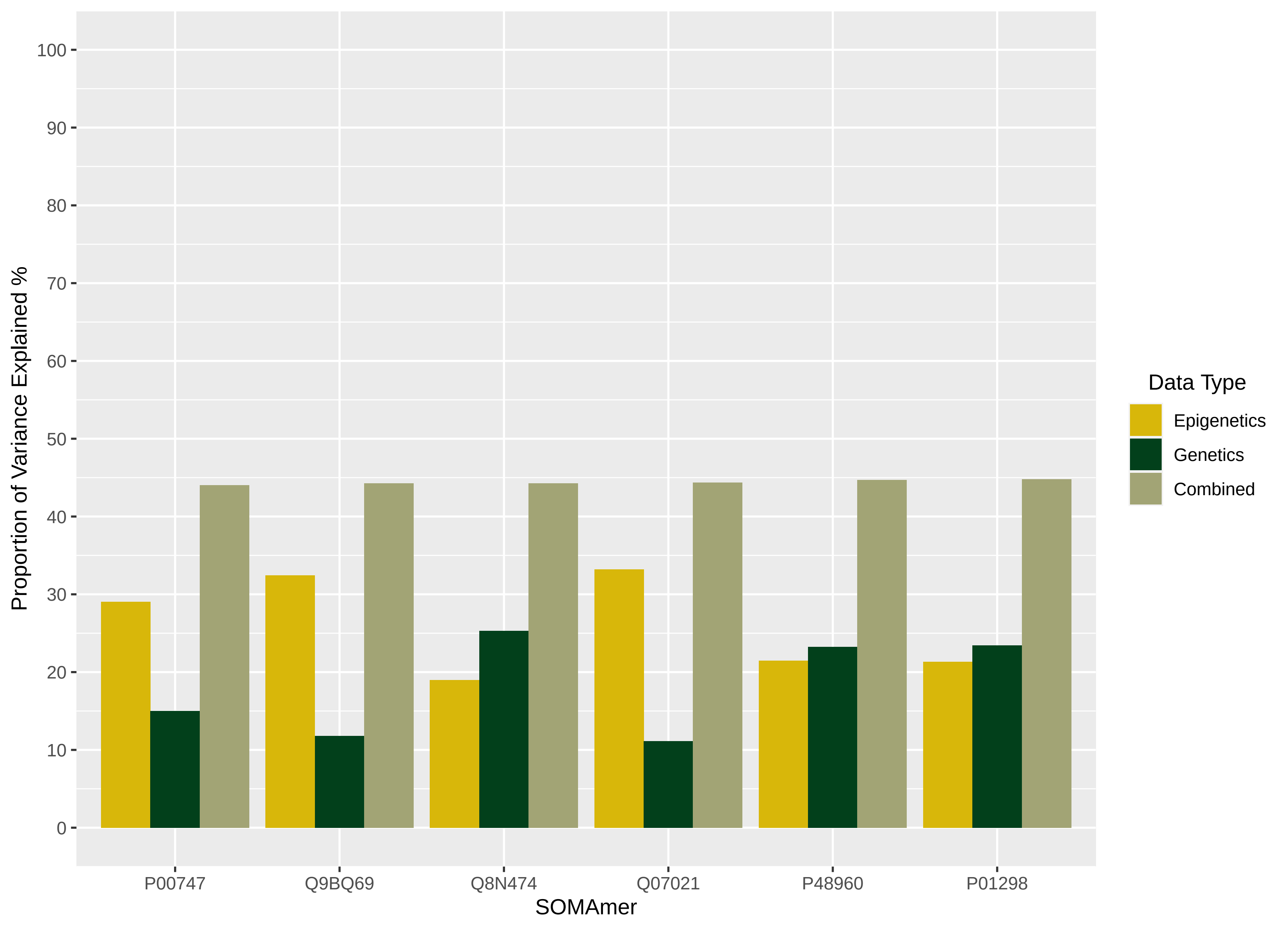

Proportion of Variance Explained %
100
90
80
70
60
50
40
30
20
10
0
P00747
Q9BQ69
Q8N474
Q07021
P48960
P01298
SOMAmer
Data Type
Epigenetics
Genetics
Combined

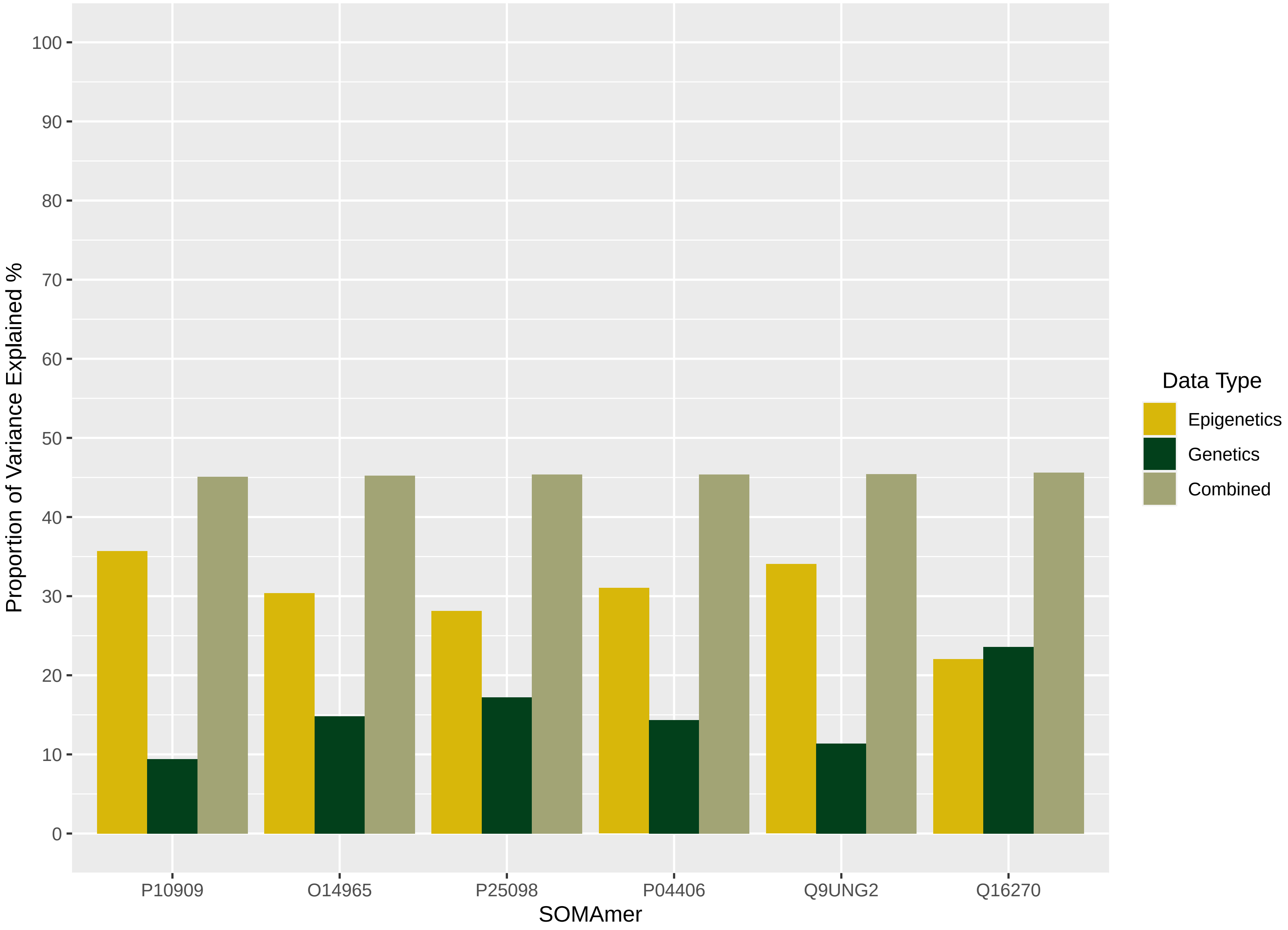

Proportion of Variance Explained %
100
90
80
70
60
50
40
30
20
10
0
P10909
O14965
P25098
P04406
Q9UNG2
Q16270
SOMAmer
Data Type
Epigenetics
Genetics
Combined

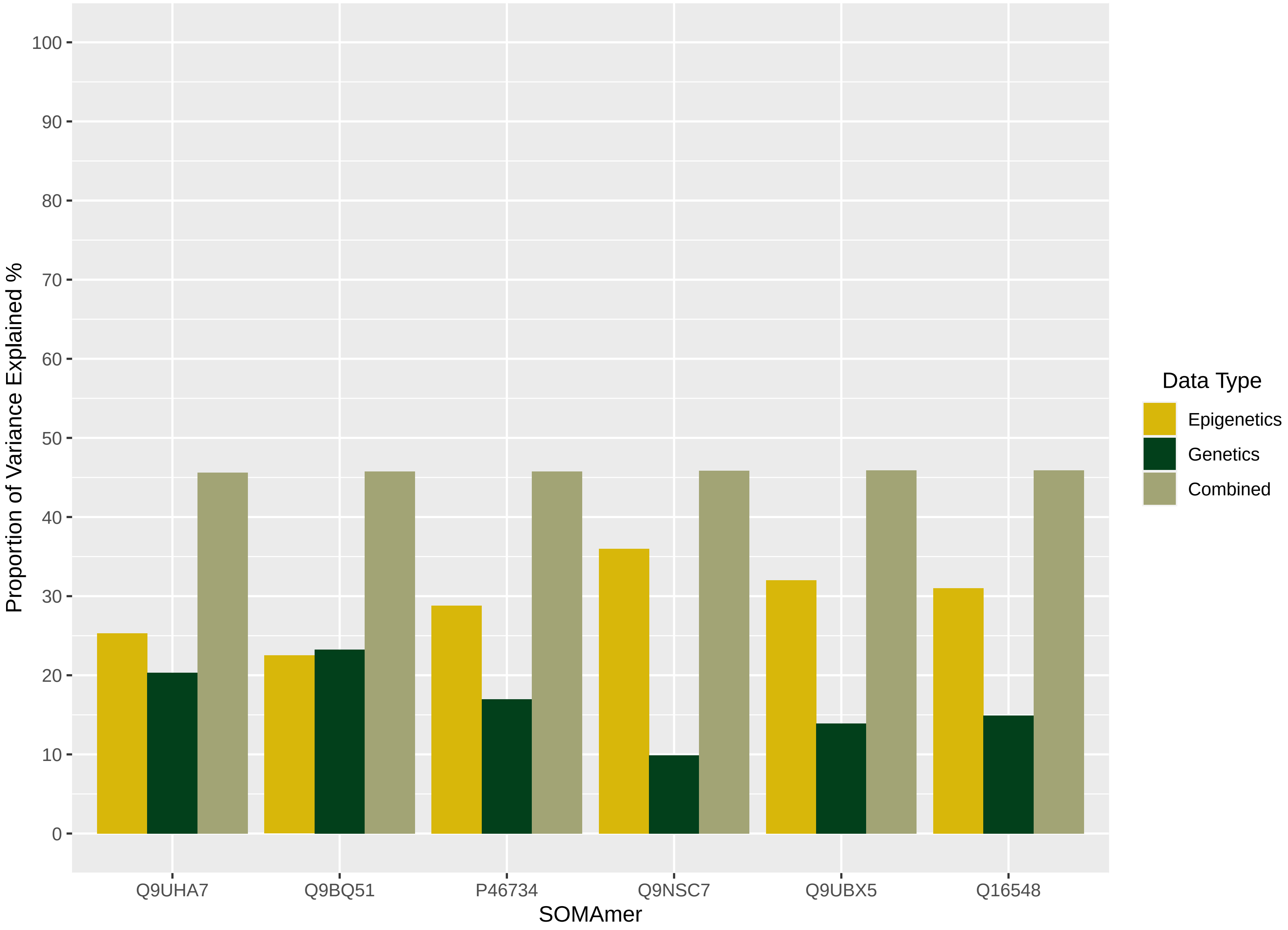

Proportion of Variance Explained %
0
10
20
30
40
50
60
70
80
90
100
Q9UHA7
Q9BQ51
P46734
Q9NSC7
Q9UBX5
Q16548
SOMAmer
Data Type
Epigenetics
Genetics
Combined

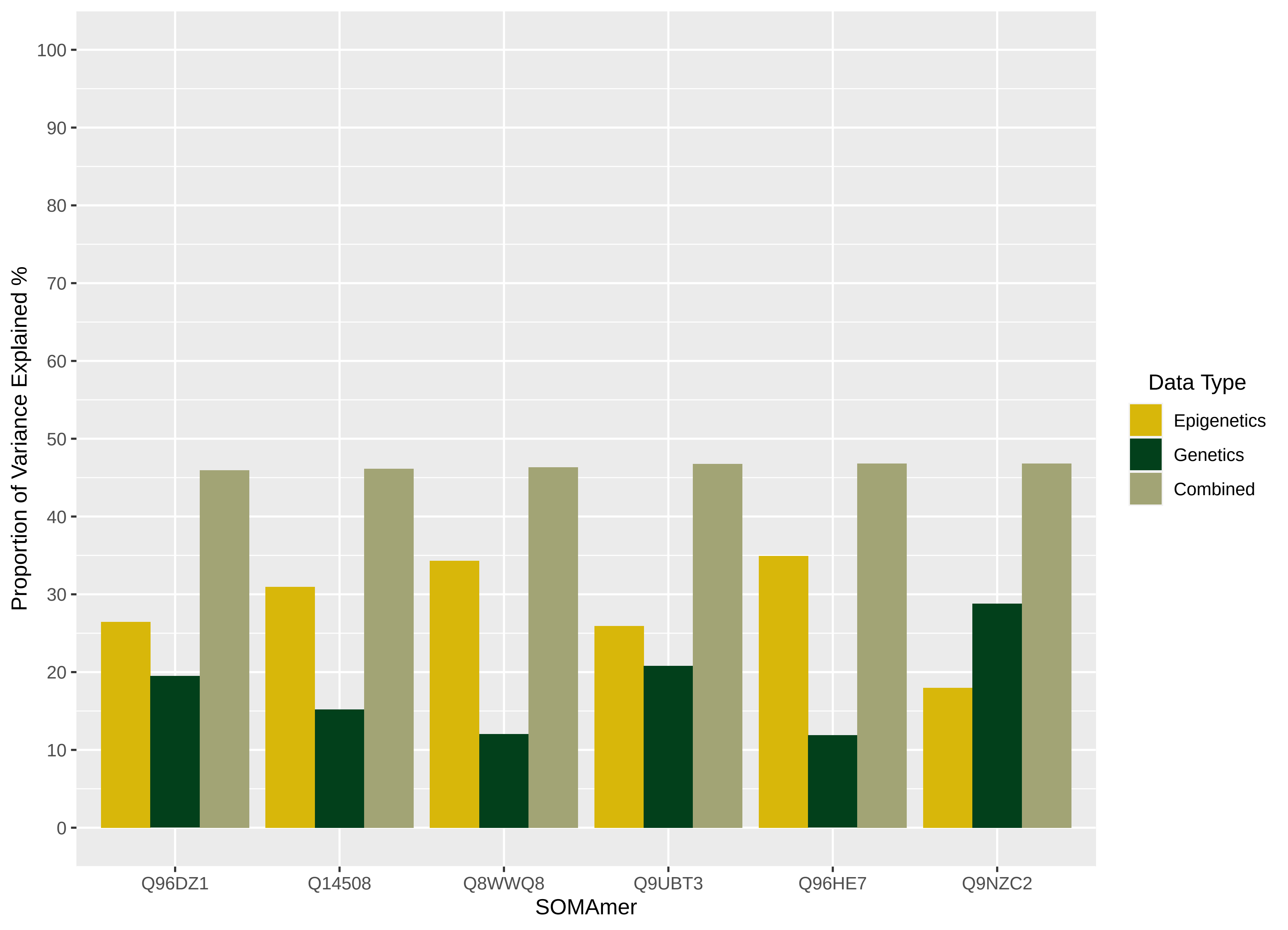

Proportion of Variance Explained %
100
90
80
70
60
50
40
30
20
10
0
Q96DZ1
Q14508
Q8WWQ8
Q9UBT3
Q96HE7
Q9NZC2
SOMAmer
Data Type
Epigenetics
Genetics
Combined

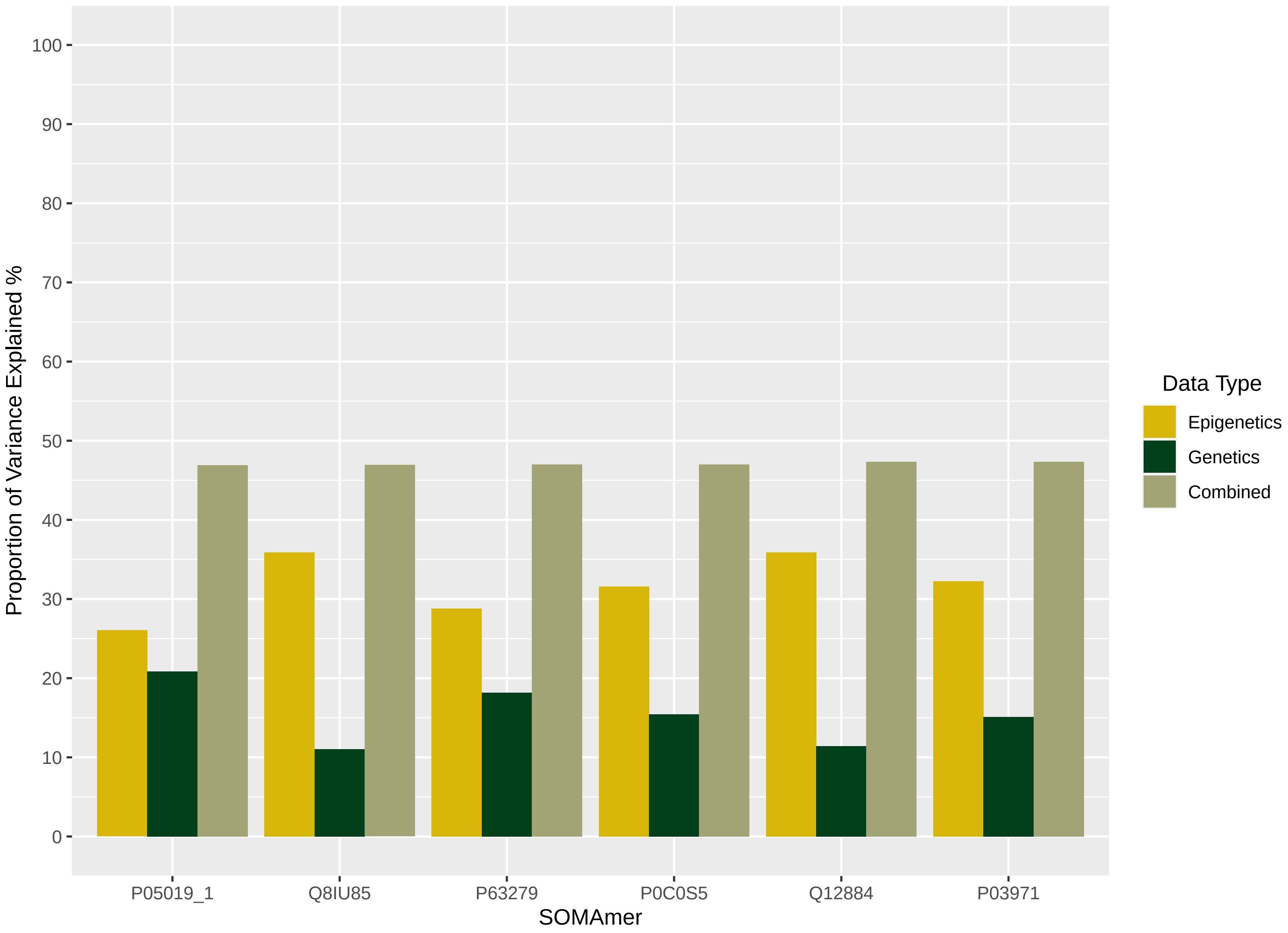

Proportion of Variance Explained %
100
90
80
70
60
50
40
30
20
10
0
P05019_1
Q8IU85
P63279
P0C0S5
Q12884
P03971
SOMAmer
Data Type
Epigenetics
Genetics
Combined

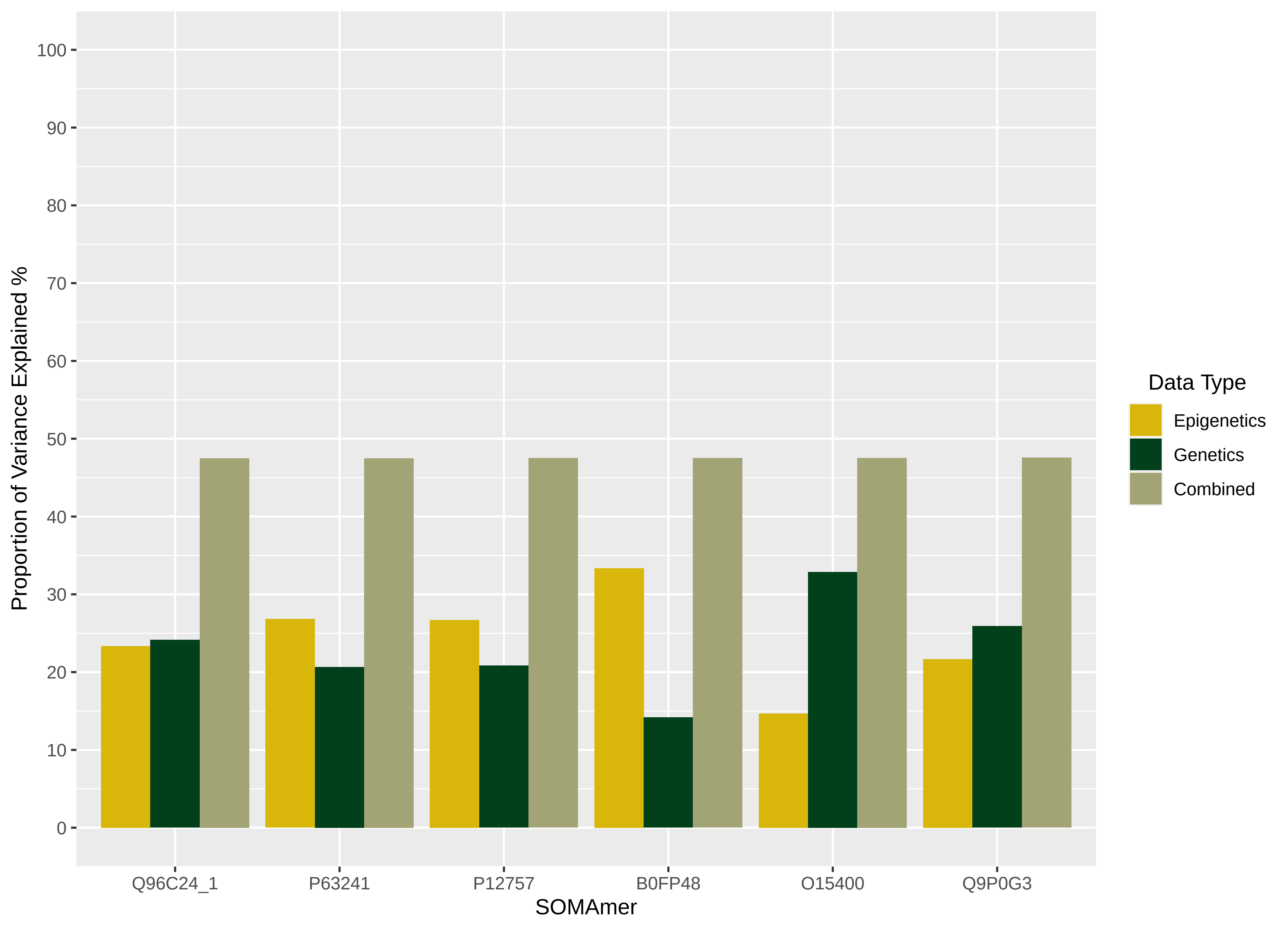
Proportion of Variance Explained %
100
90
80
70
60
50
40
30
20
10
0
Q96C24_1
P63241
P12757
B0FP48
O15400
Q9P0G3
SOMAmer
Data Type
Epigenetics
Genetics
Combined

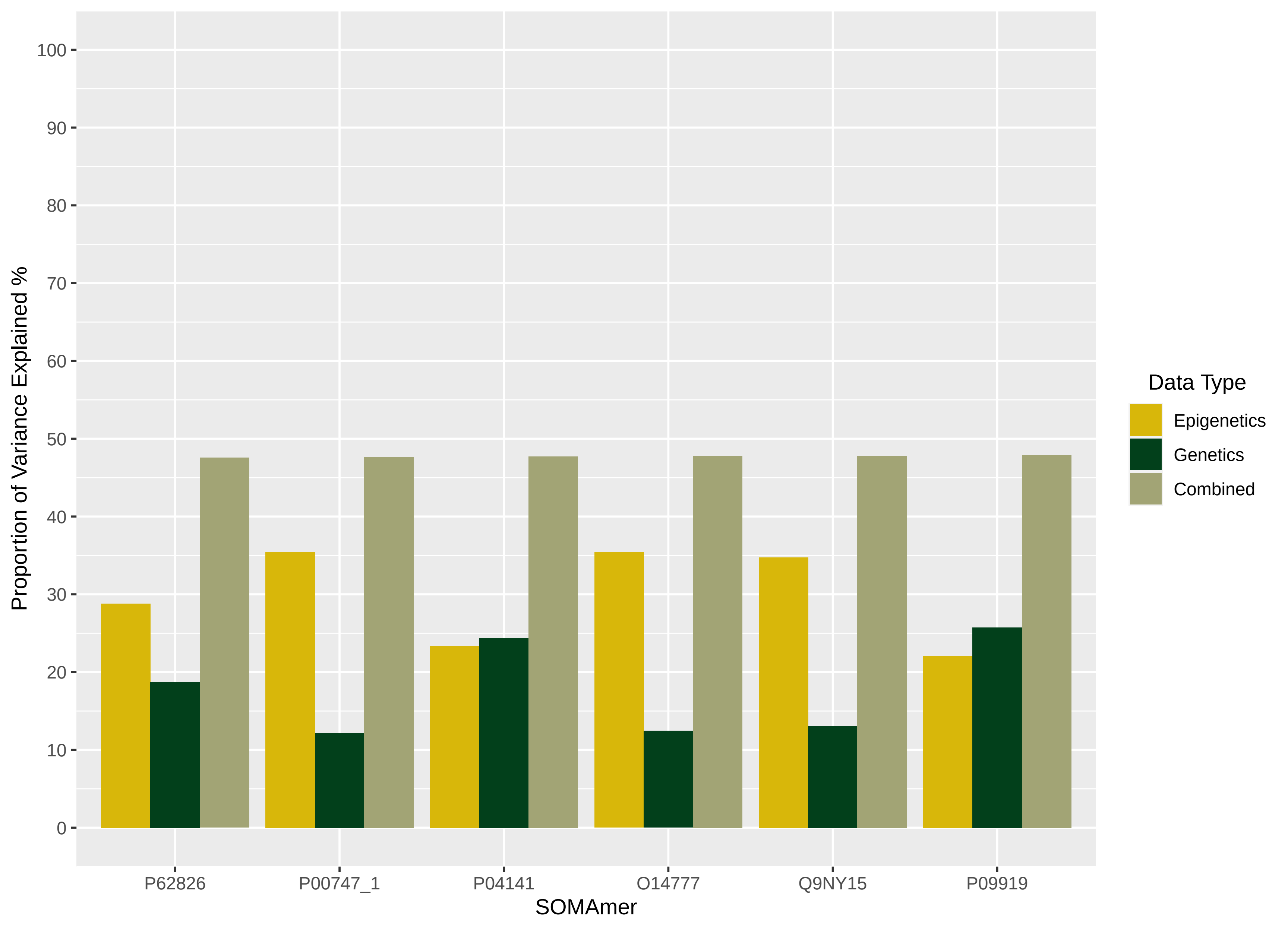

Proportion of Variance Explained %
100
90
80
70
60
50
40
30
20
10
0
P62826
P00747_1
P04141
O14777
Q9NY15
P09919
SOMAmer
Data Type
Epigenetics
Genetics
Combined

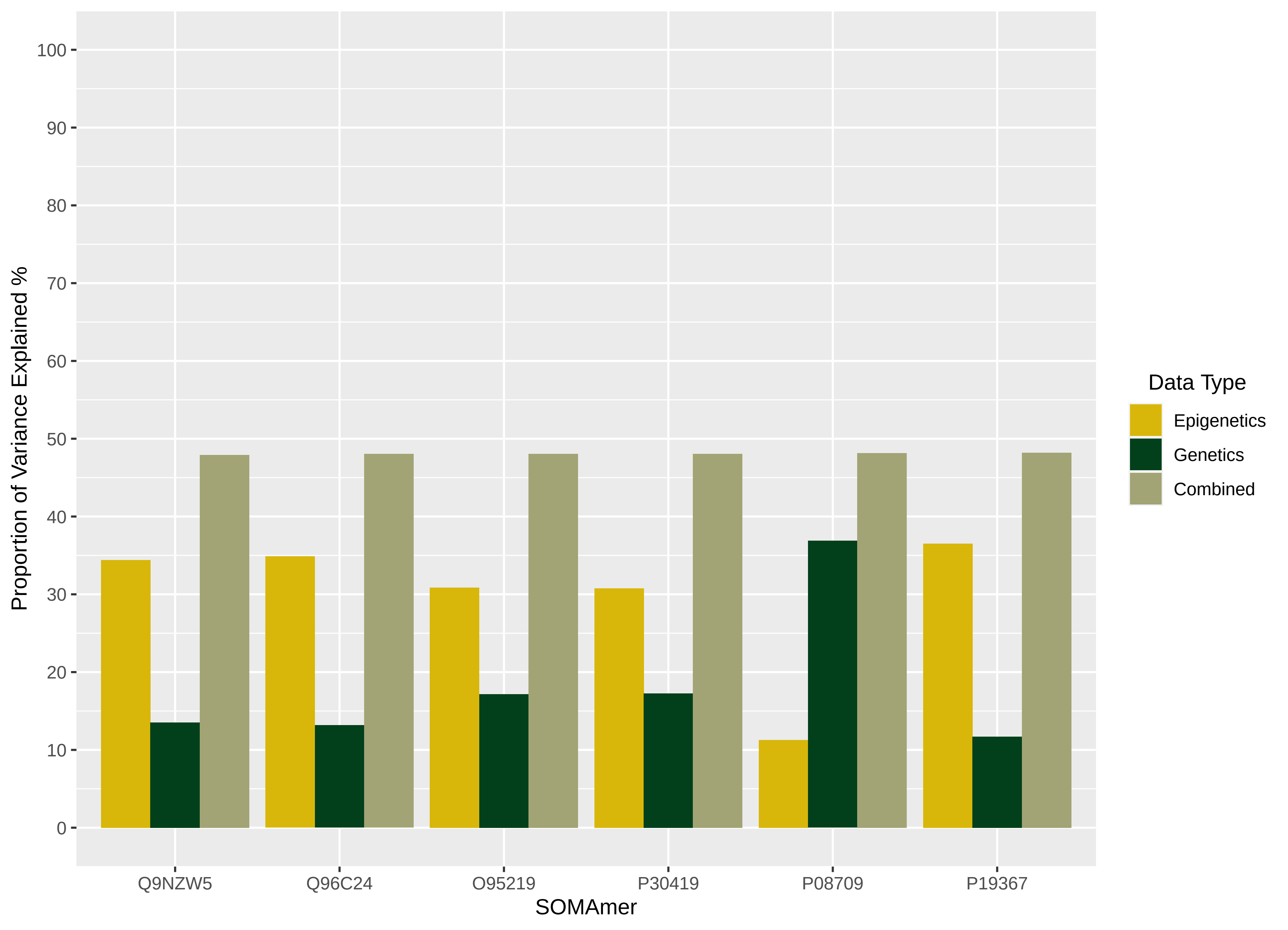

Proportion of Variance Explained %
100
90
80
70
60
50
40
30
20
10
0
Q9NZW5
Q96C24
O95219
P30419
P08709
P19367
SOMAmer
Data Type
Epigenetics
Genetics
Combined

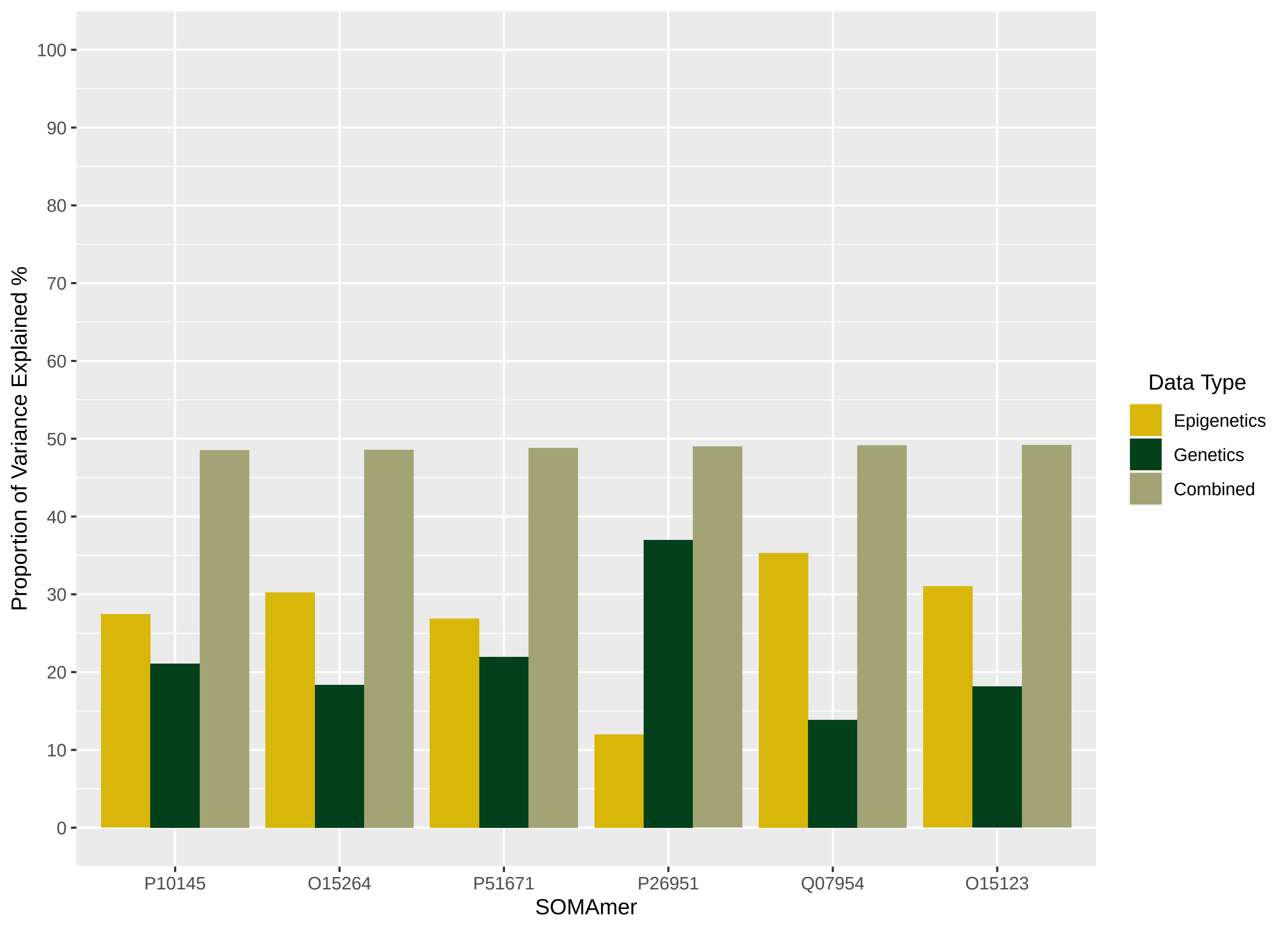

Proportion of Variance Explained %
100
90
80
70
60
50
40
30
20
10
0
P10145
O15264
P51671
P26951
Q07954
O15123
SOMAmer
Data Type
Epigenetics
Genetics
Combined

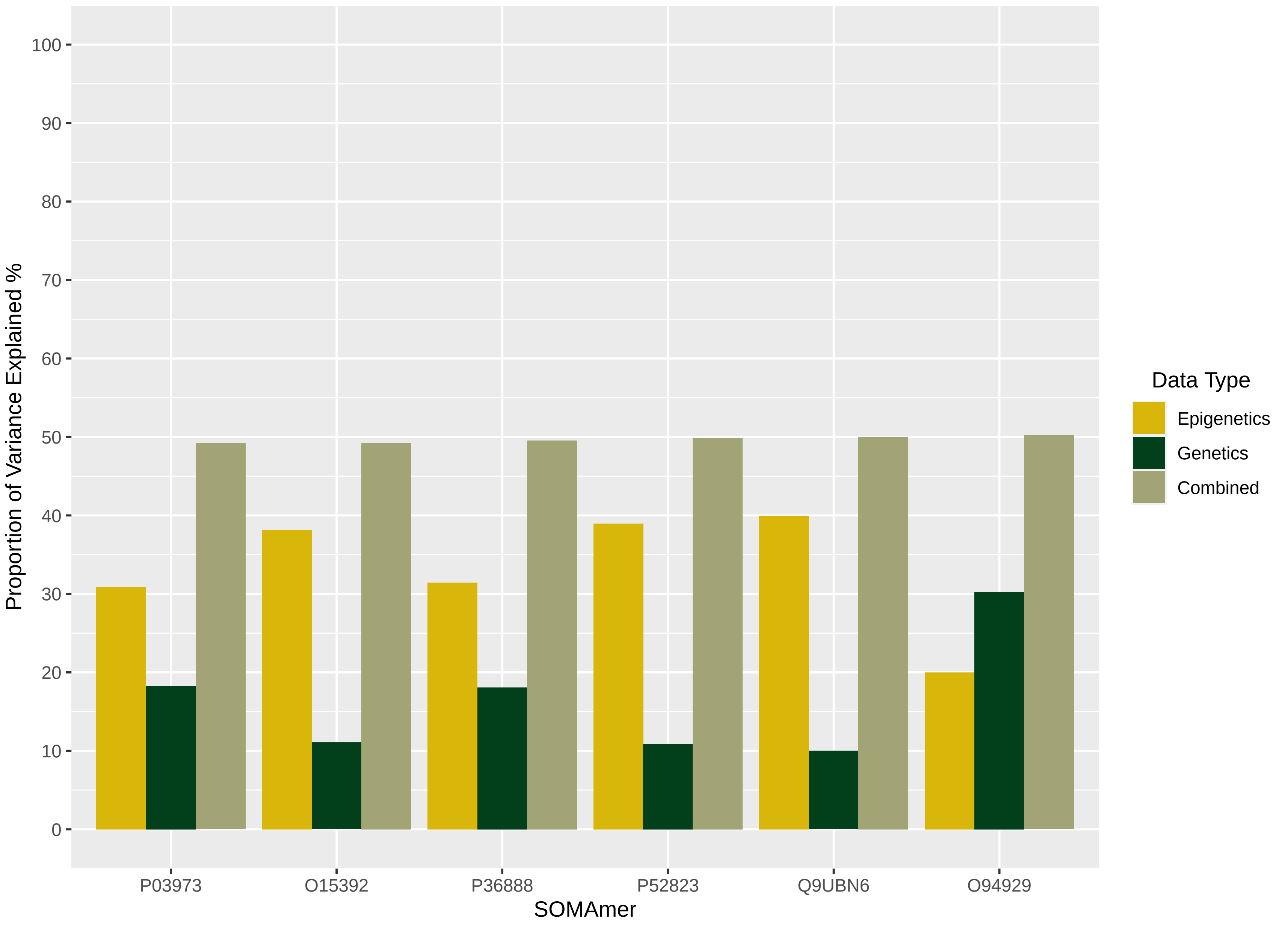

Proportion of Variance Explained %
100
90
80
70
60
50
40
30
20
10
0
P03973
O15392
P36888
P52823
Q9UBN6
O94929
SOMAmer
Data Type
Epigenetics
Genetics
Combined

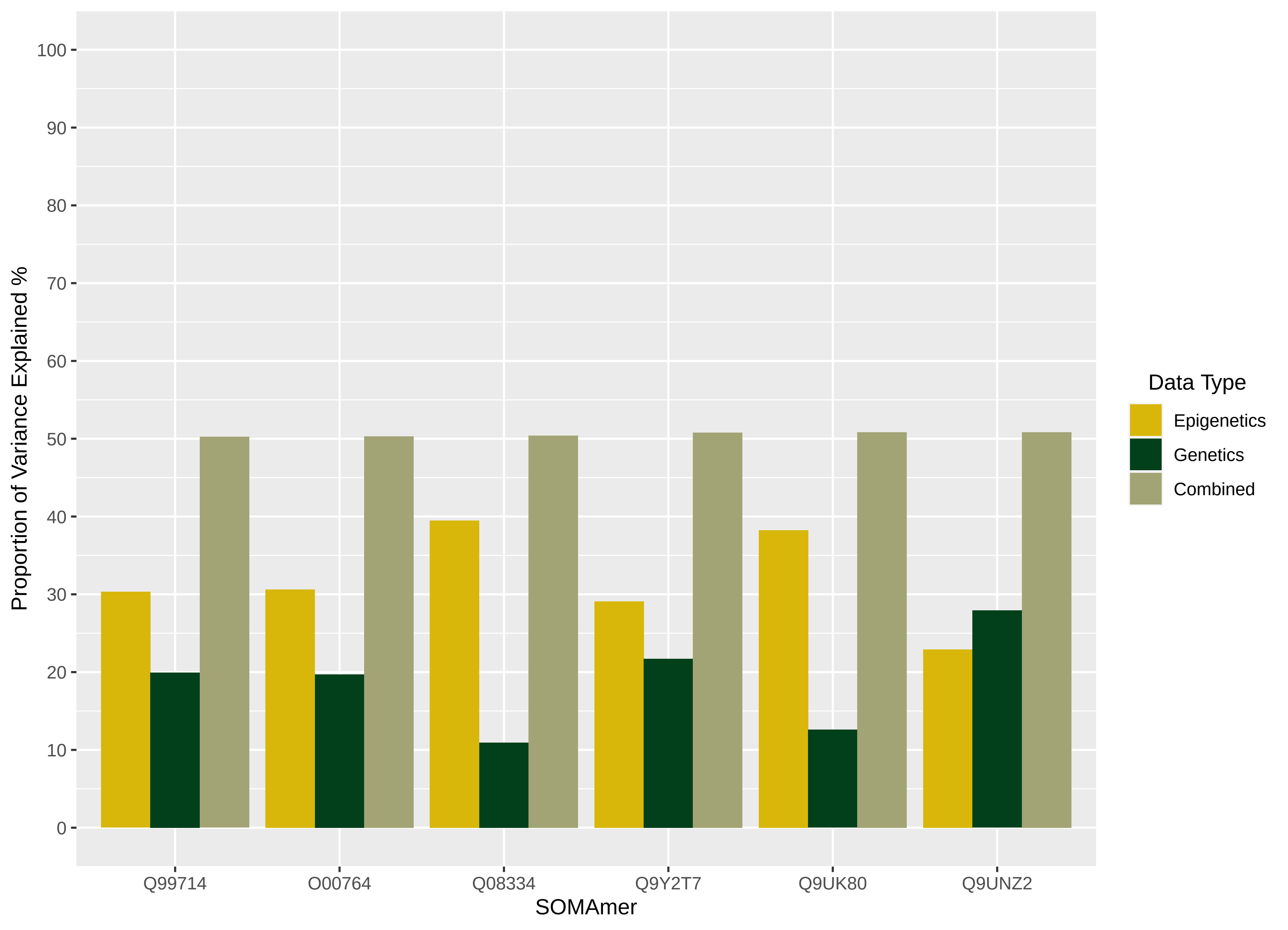

Proportion of Variance Explained %
100
90
80
70
60
50
40
30
20
10
0
Q99714
O00764
Q08334
Q9Y2T7
Q9UK80
Q9UNZ2
SOMAmer
Data Type
Epigenetics
Genetics
Combined

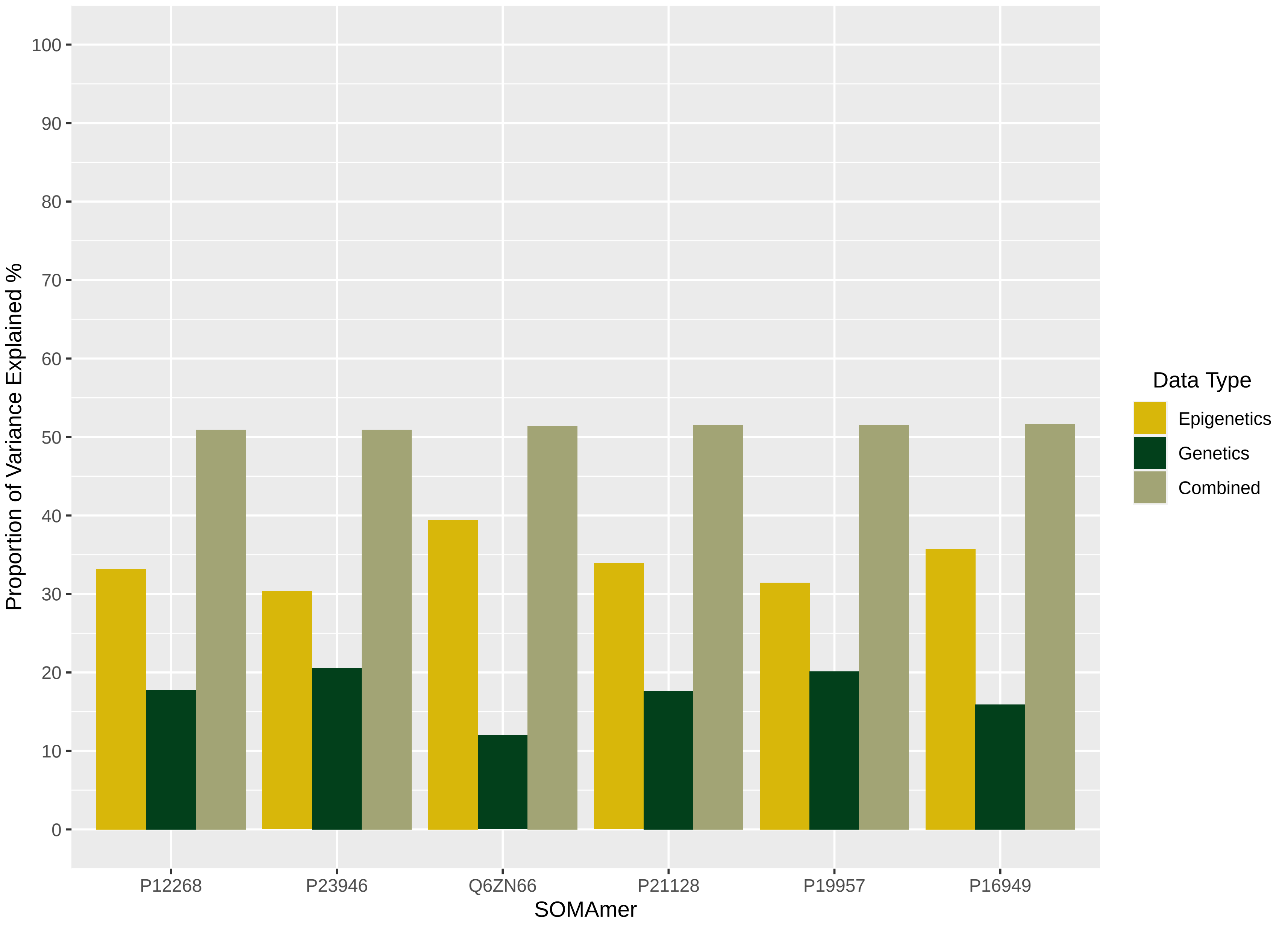

Proportion of Variance Explained %
0
10
20
30
40
50
60
70
80
90
100
P12268
P23946
Q6ZN66
P21128
P19957
P16949
SOMAmer
Data Type
Epigenetics
Genetics
Combined

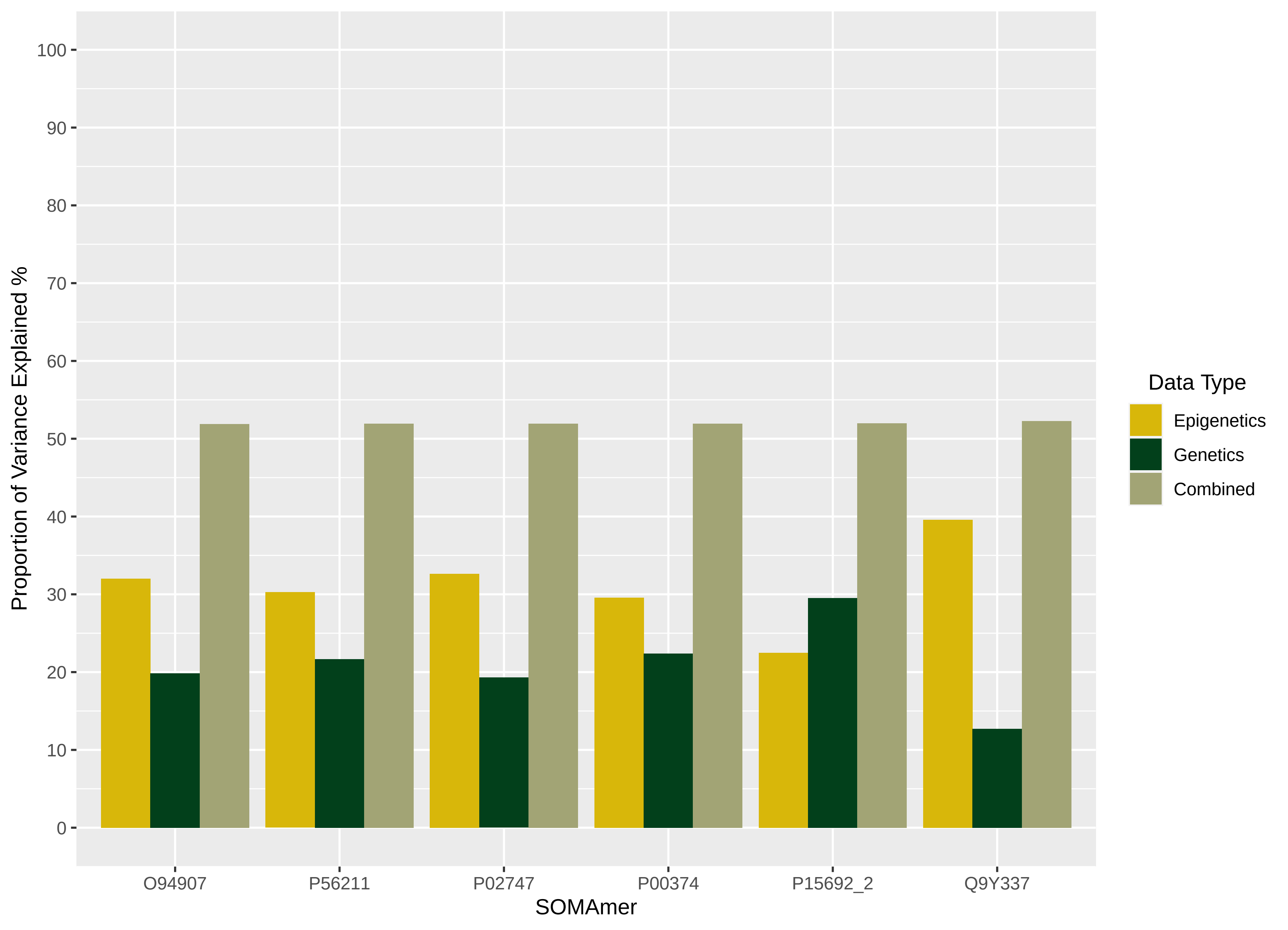

Proportion of Variance Explained %
100
90
80
70
60
50
40
30
20
10
0
O94907
P56211
P02747
P00374
P15692_2
Q9Y337
SOMAmer
Data Type
Epigenetics
Genetics
Combined

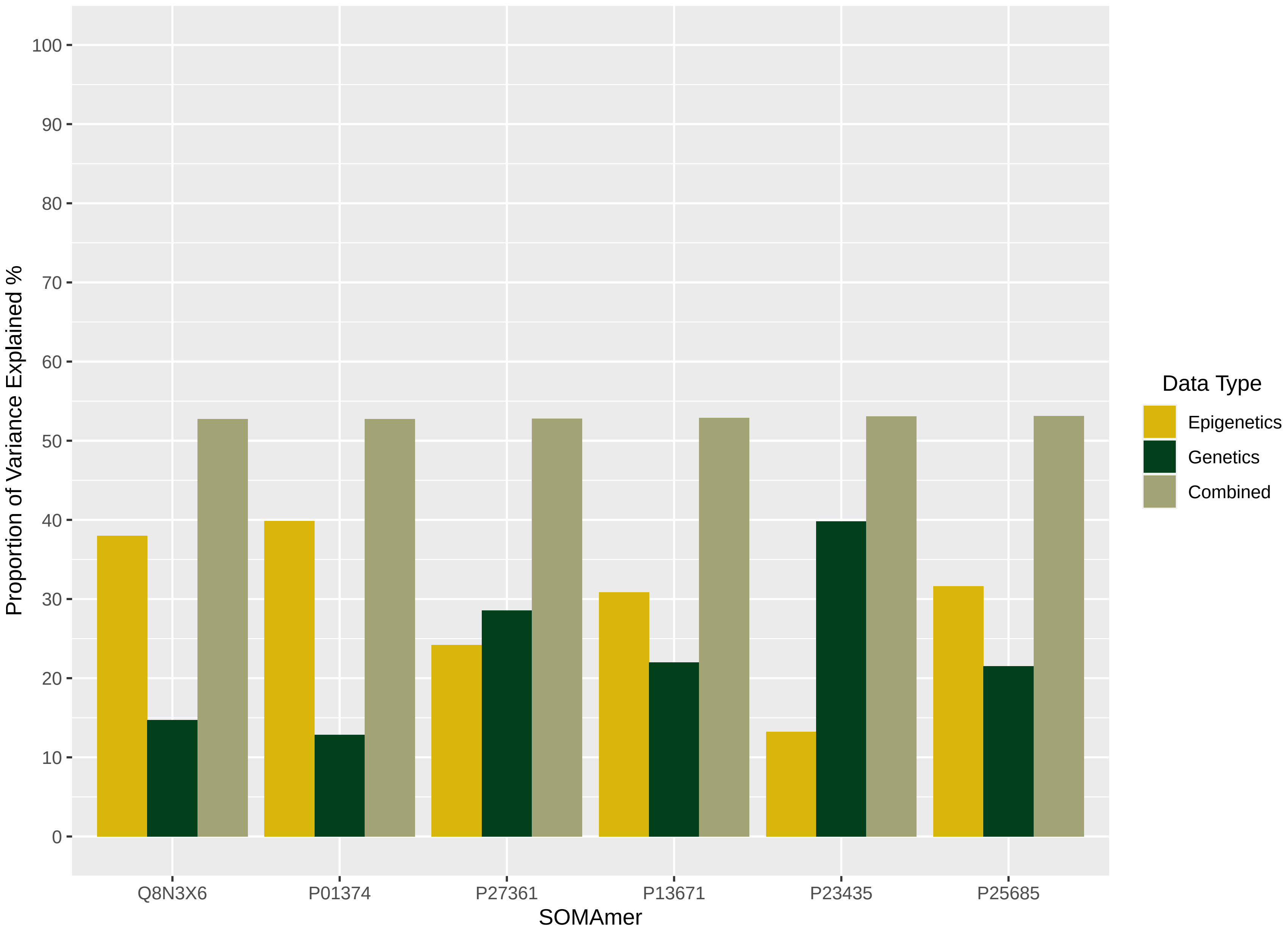

Proportion of Variance Explained %
100
90
80
70
60
50
40
30
20
10
0
Q8N3X6
P01374
P27361
P13671
P23435
P25685
SOMAmer
Data Type
Epigenetics
Genetics
Combined

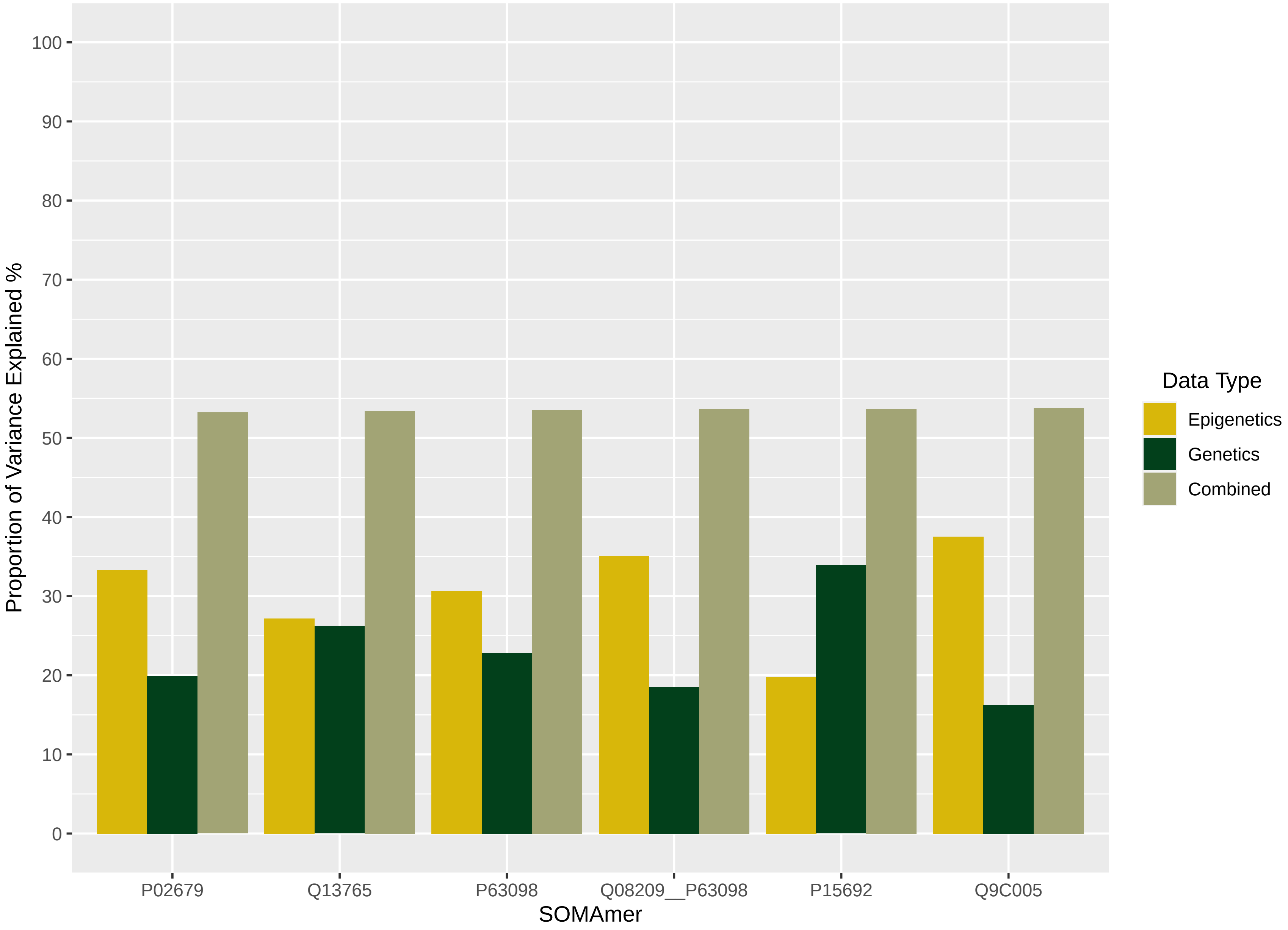

Proportion of Variance Explained %
100
90
80
70
60
50
40
30
20
10
0
P02679
Q13765
P63098
Q08209__P63098
P15692
Q9C005
SOMAmer
Data Type
Epigenetics
Genetics
Combined

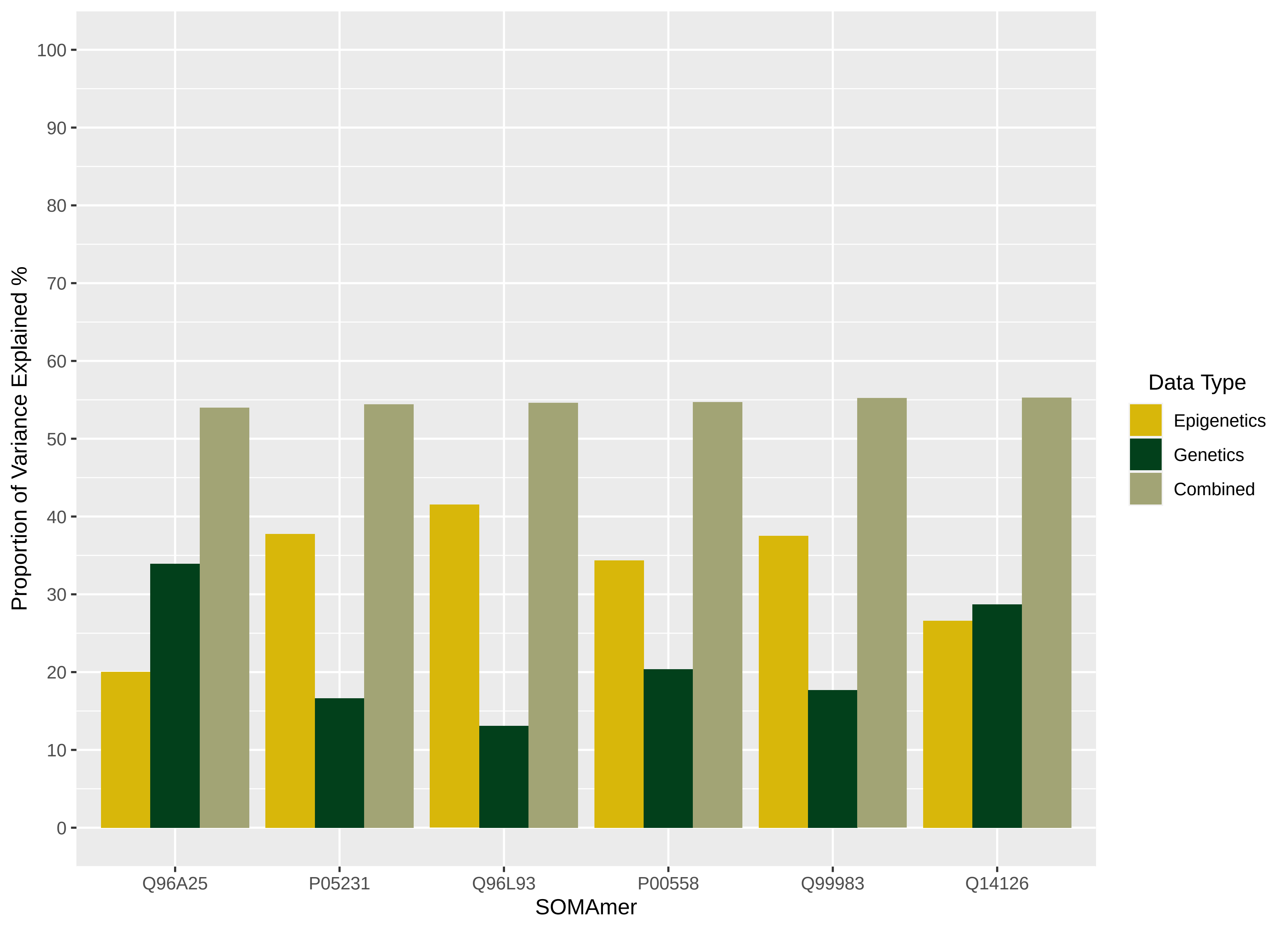

Proportion of Variance Explained %
0
10
20
30
40
50
60
70
80
90
100
Q96A25
P05231
Q96L93
P00558
Q99983
Q14126
SOMAmer
Data Type
Epigenetics
Genetics
Combined

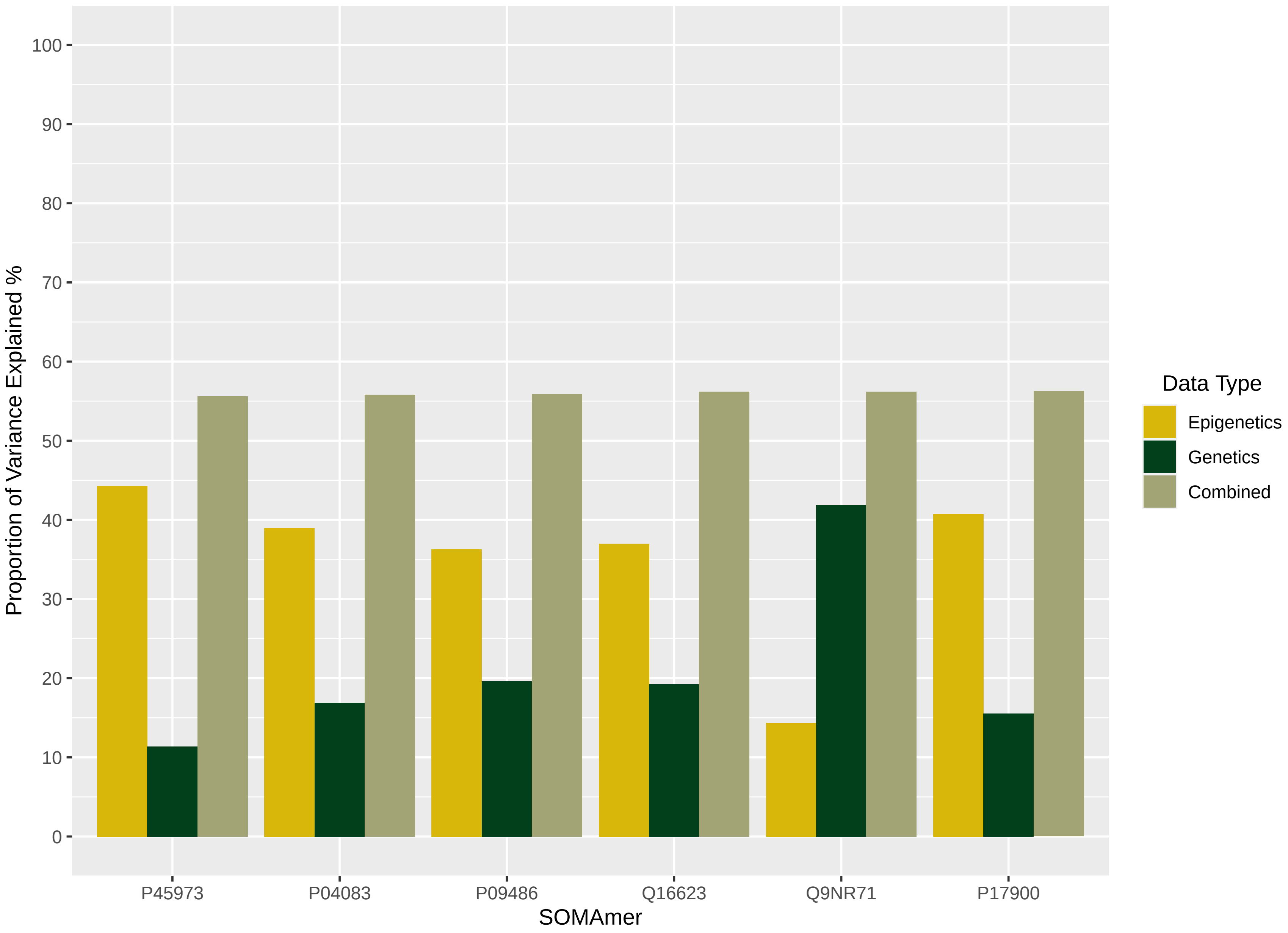

Proportion of Variance Explained %
100
90
80
70
60
50
40
30
20
10
0
P45973
P04083
P09486
Q16623
Q9NR71
P17900
SOMAmer
Data Type
Epigenetics
Genetics
Combined

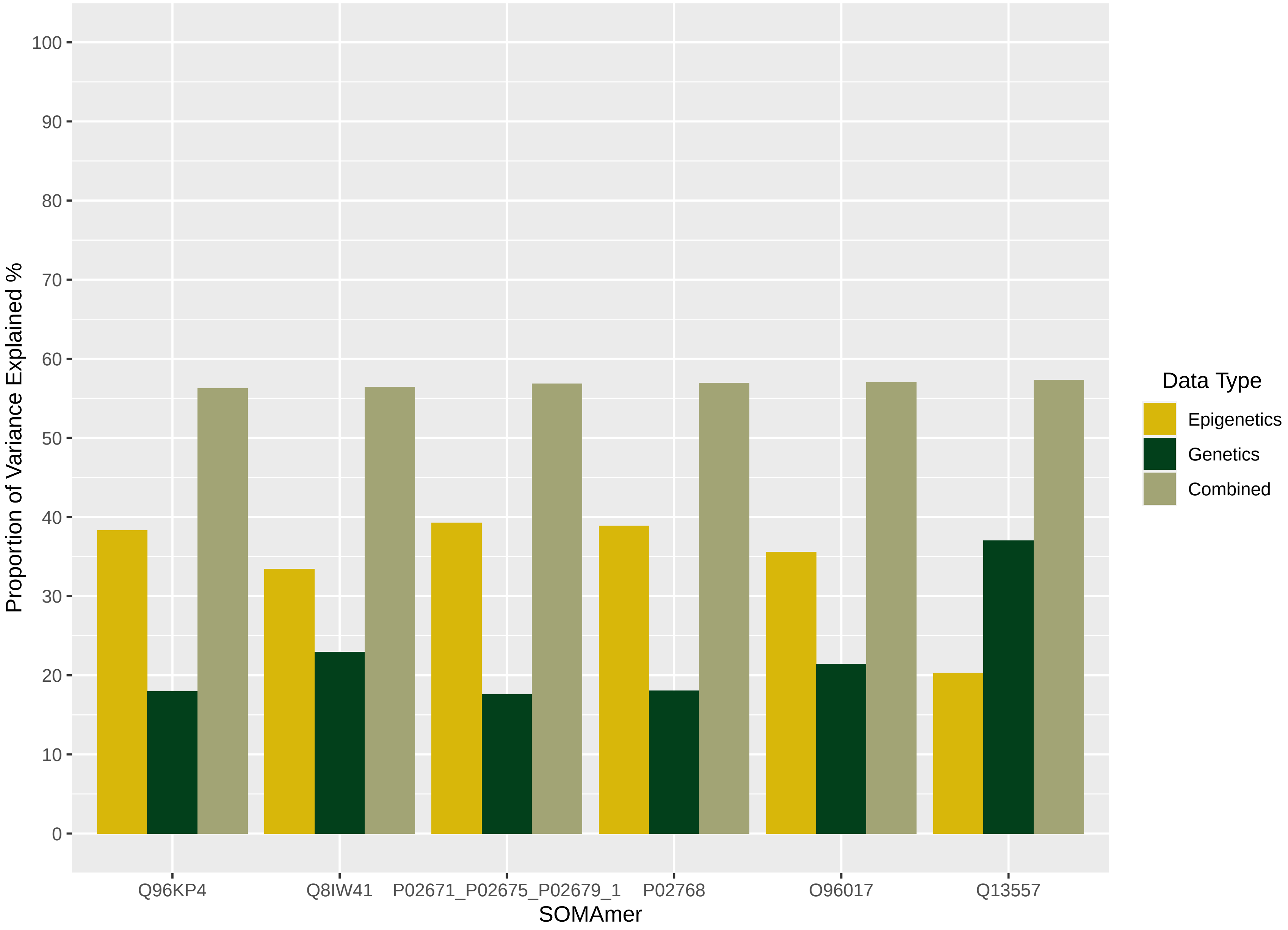

Proportion of Variance Explained %
100
90
80
70
60
50
40
30
20
10
0
Q96KP4
Q8IW41
P02671_P02675_P02679_1
P02768
O96017
Q13557
SOMAmer
Data Type
Epigenetics
Genetics
Combined

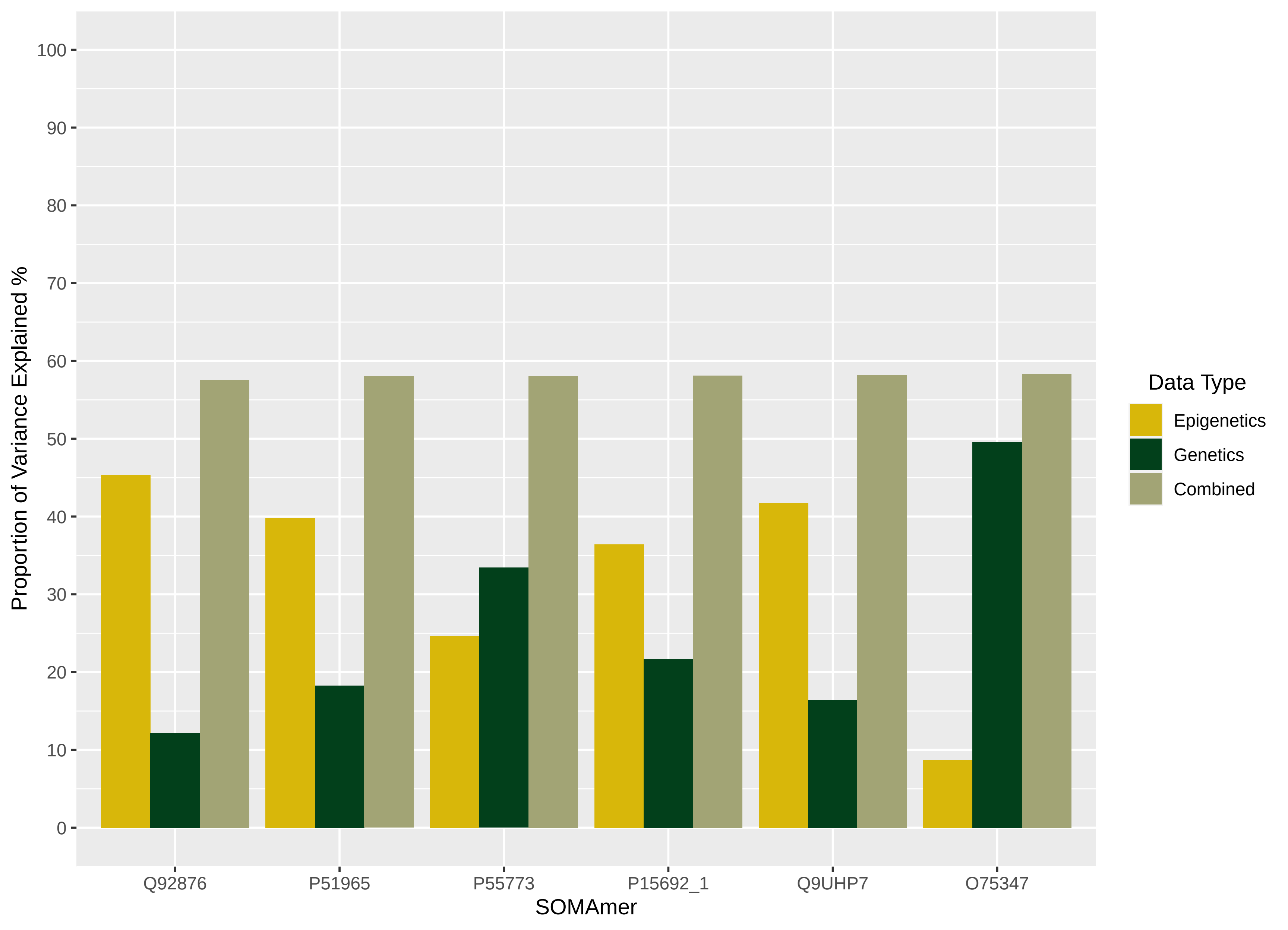

Proportion of Variance Explained %
0
10
20
30
40
50
60
70
80
90
100
Q92876
P51965
P55773
P15692_1
Q9UHP7
O75347
SOMAmer
Data Type
Epigenetics
Genetics
Combined

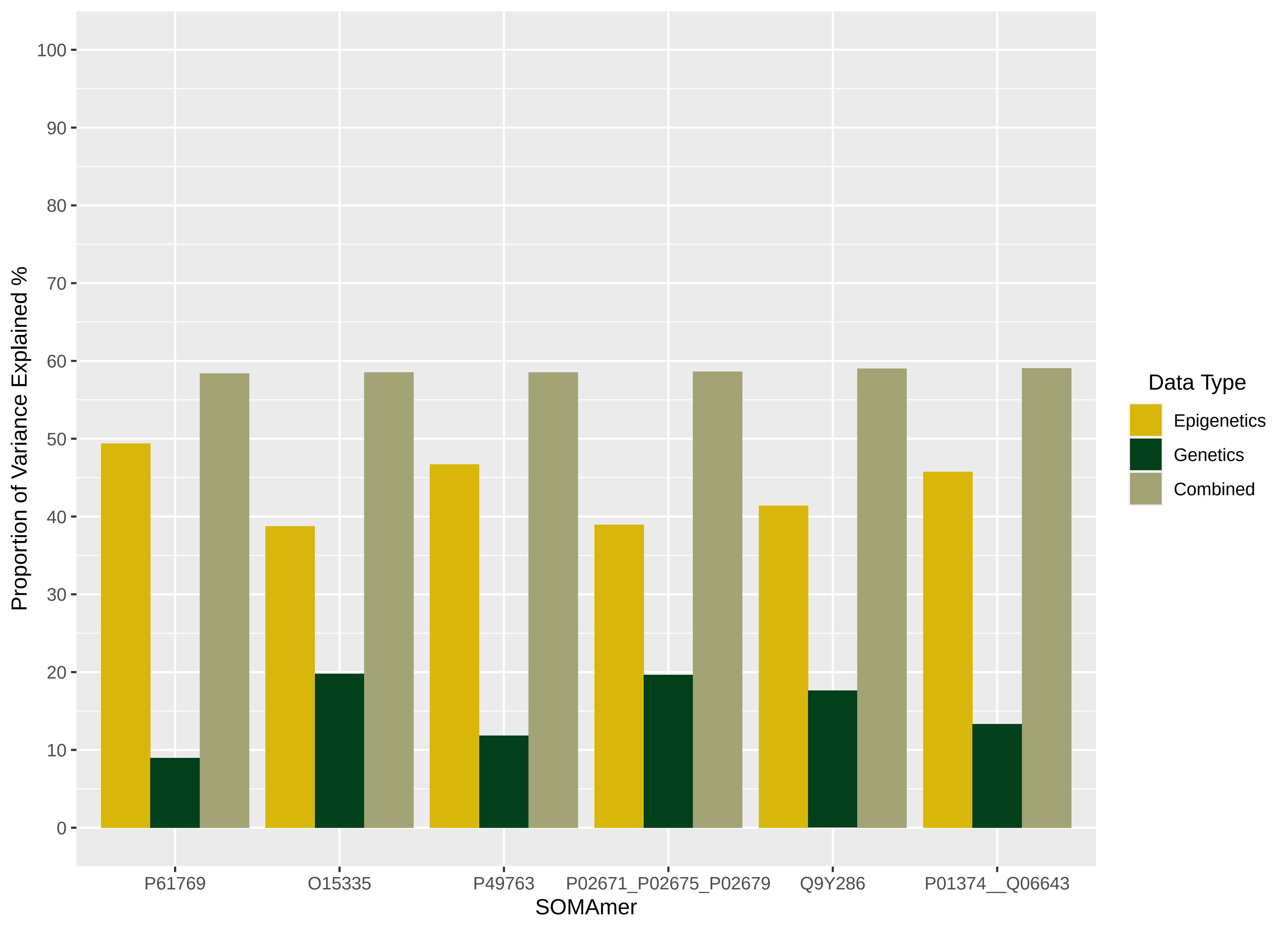

Proportion of Variance Explained %
100
90
80
70
60
50
40
30
20
10
0
P61769
O15335
P49763
P02671_P02675_P02679
Q9Y286
P01374__Q06643
SOMAmer
Data Type
Epigenetics
Genetics
Combined

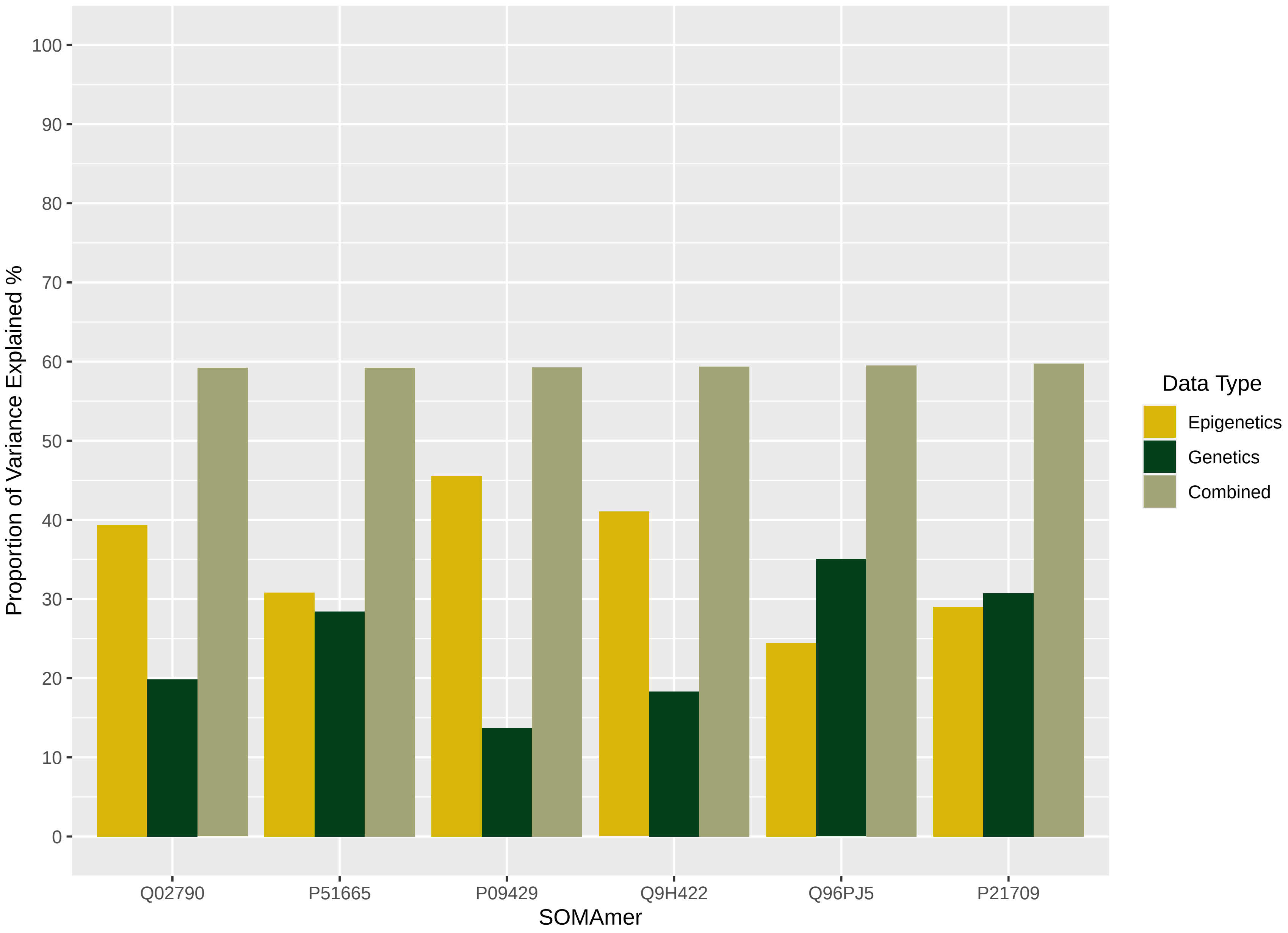

Proportion of Variance Explained %
100
90
80
70
60
50
40
30
20
10
0
Q02790
P51665
P09429
Q9H422
Q96PJ5
P21709
SOMAmer
Data Type
Epigenetics
Genetics
Combined

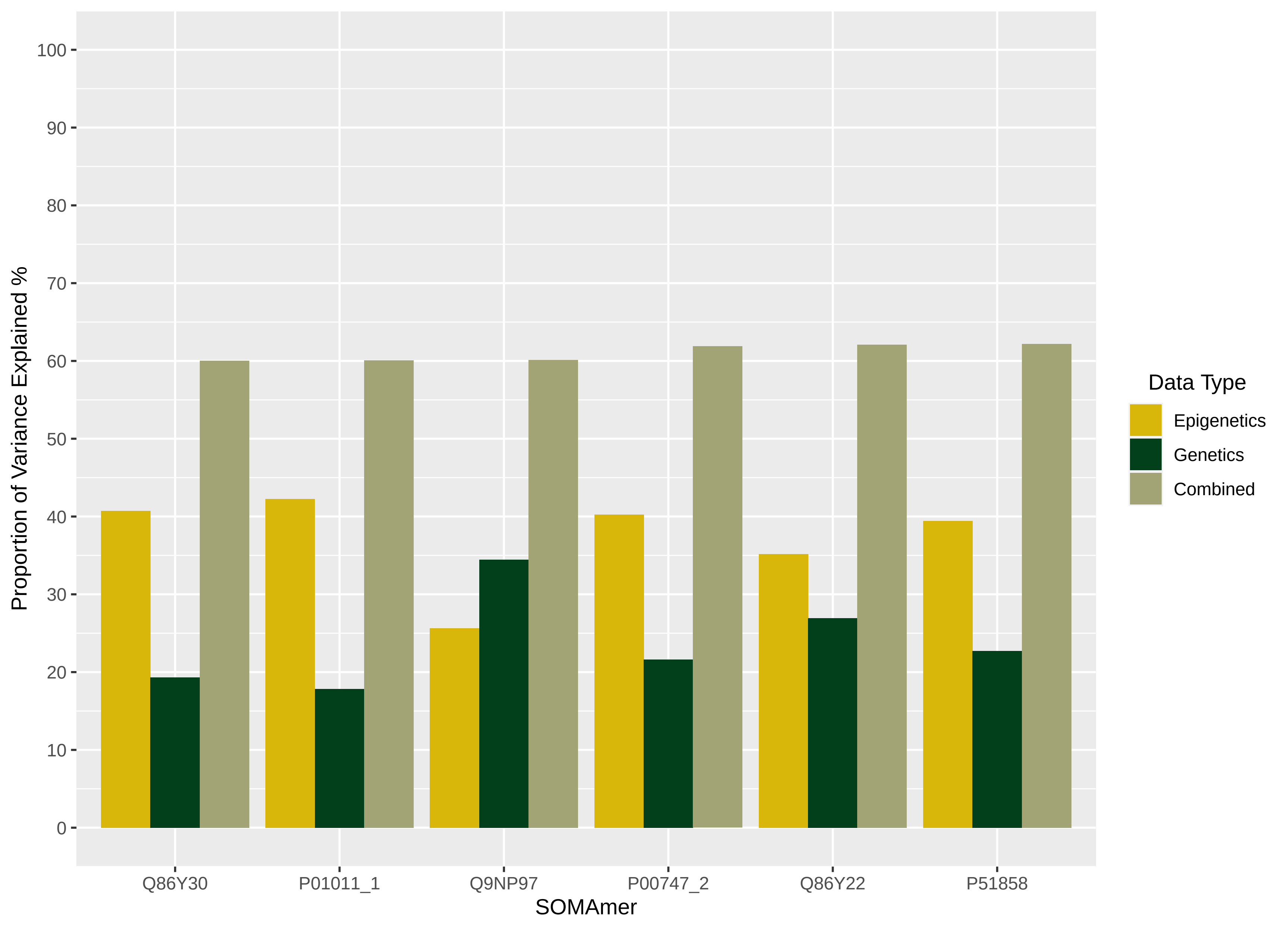

100
90
80
70
60
50
40
30
20
10
0
Proportion of Variance Explained %
Q86Y30
P01011_1
Q9NP97
P00747_2
Q86Y22
P51858
SOMAmer
Data Type
Epigenetics
Genetics
Combined

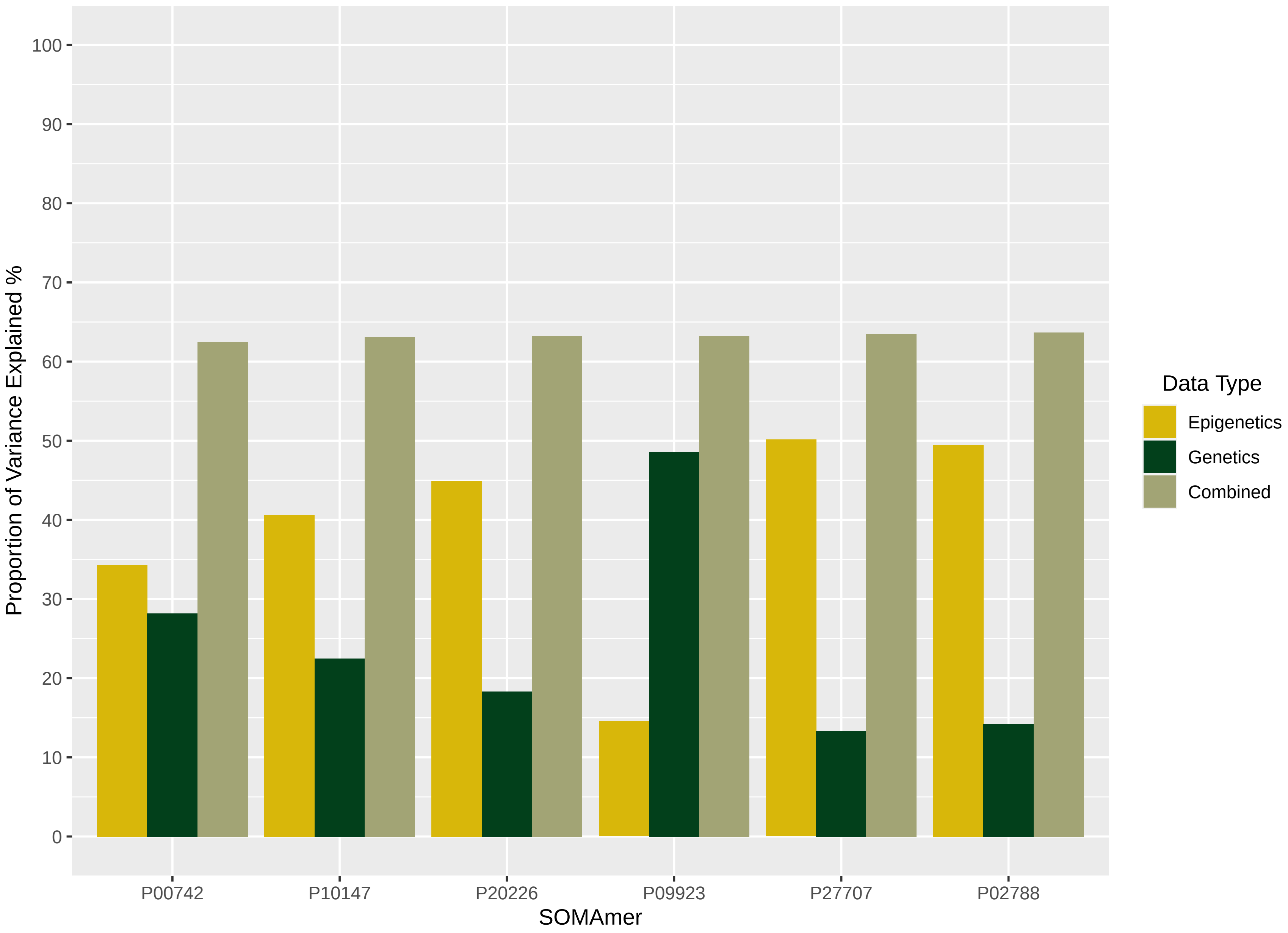

Proportion of Variance Explained %
100
90
80
70
60
50
40
30
20
10
0
P00742
P10147
P20226
P09923
P27707
P02788
SOMAmer
Data Type
Epigenetics
Genetics
Combined

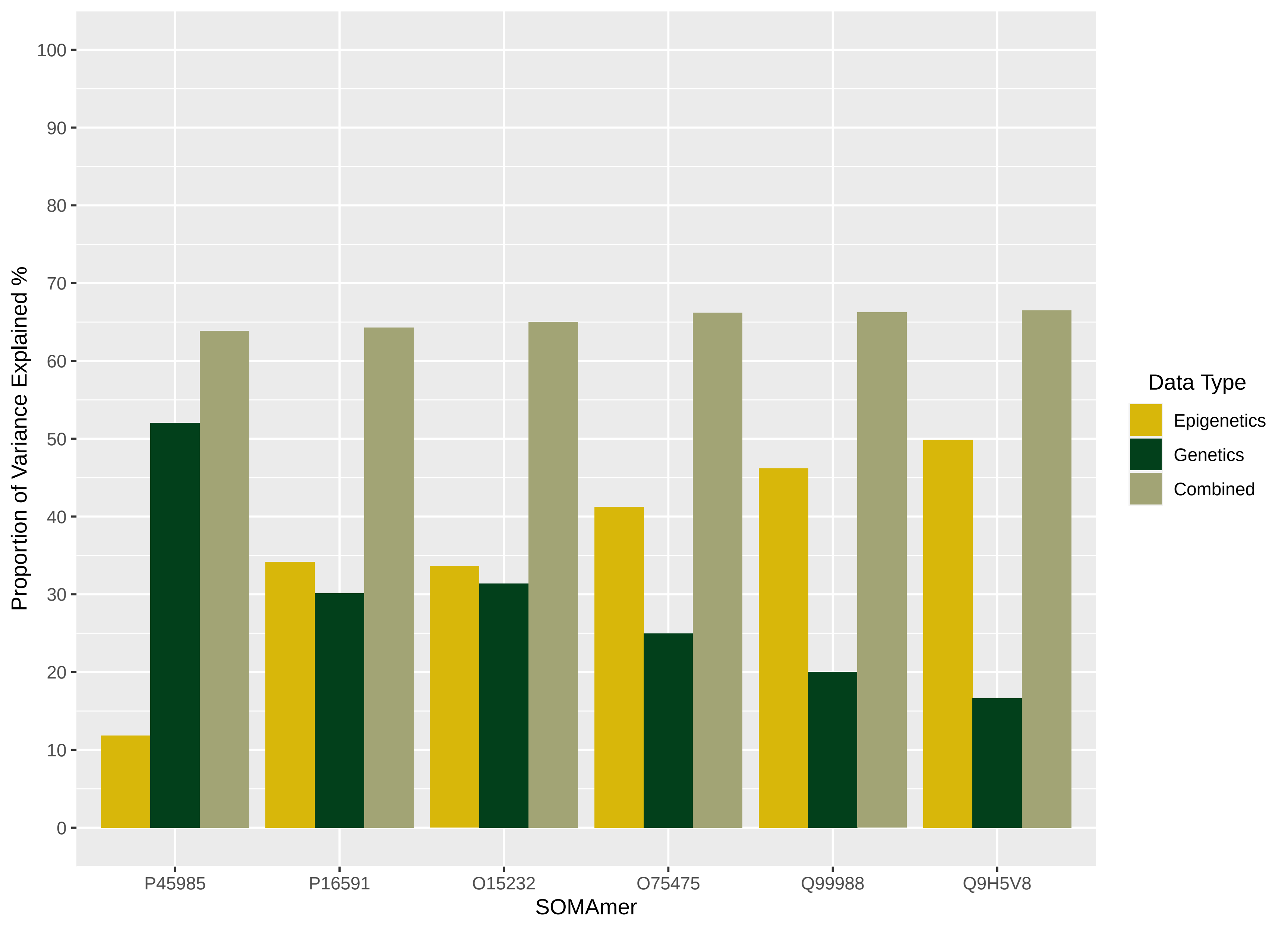

Proportion of Variance Explained %
100
90
80
70
60
50
40
30
20
10
0
P45985
P16591
O15232
O75475
Q99988
Q9H5V8
SOMAmer
Data Type
Epigenetics
Genetics
Combined

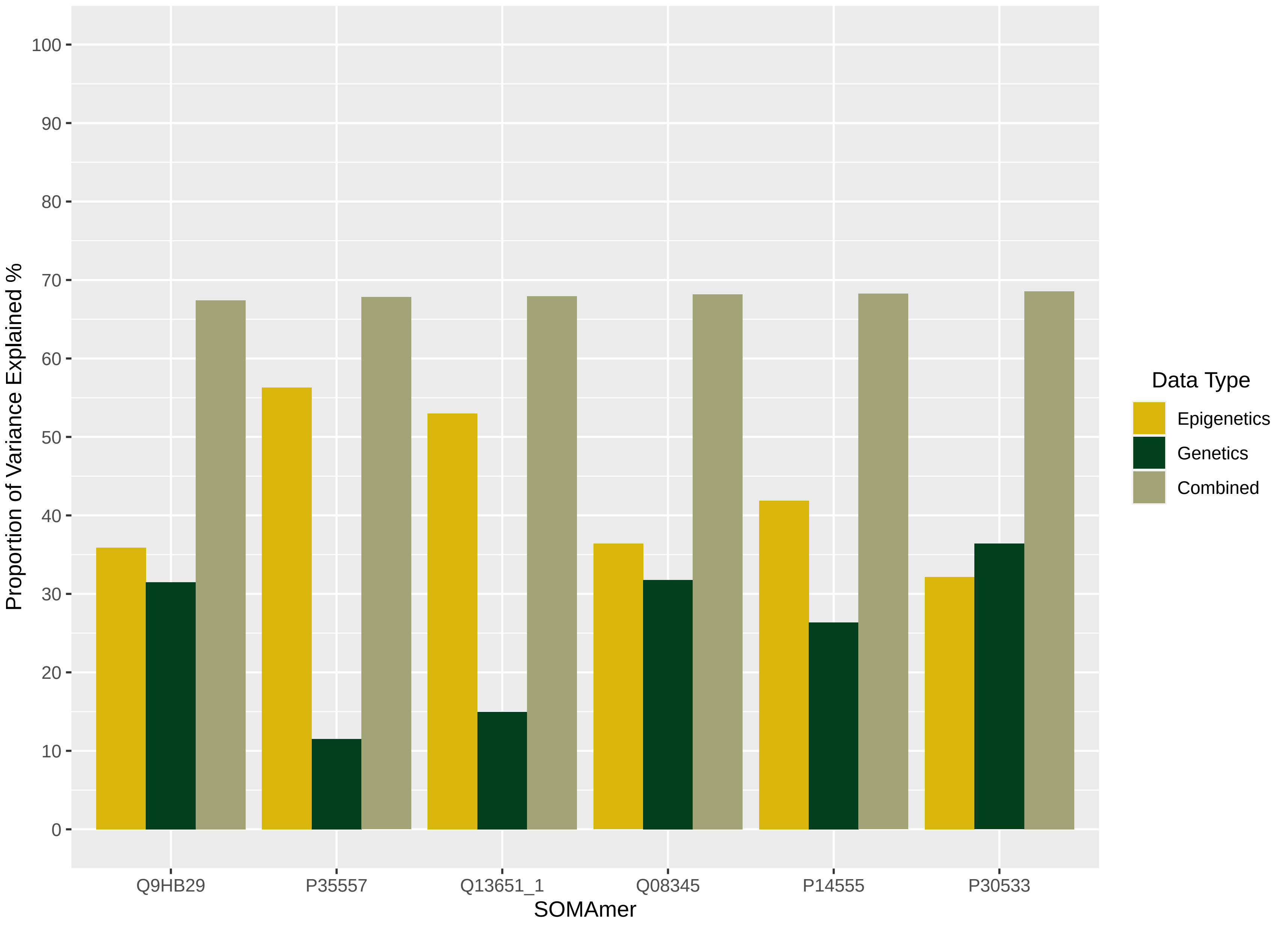

Proportion of Variance Explained %
100
90
80
70
60
50
40
30
20
10
0
Q9HB29
P35557
Q13651_1
Q08345
P14555
P30533
SOMAmer
Data Type
Epigenetics
Genetics
Combined

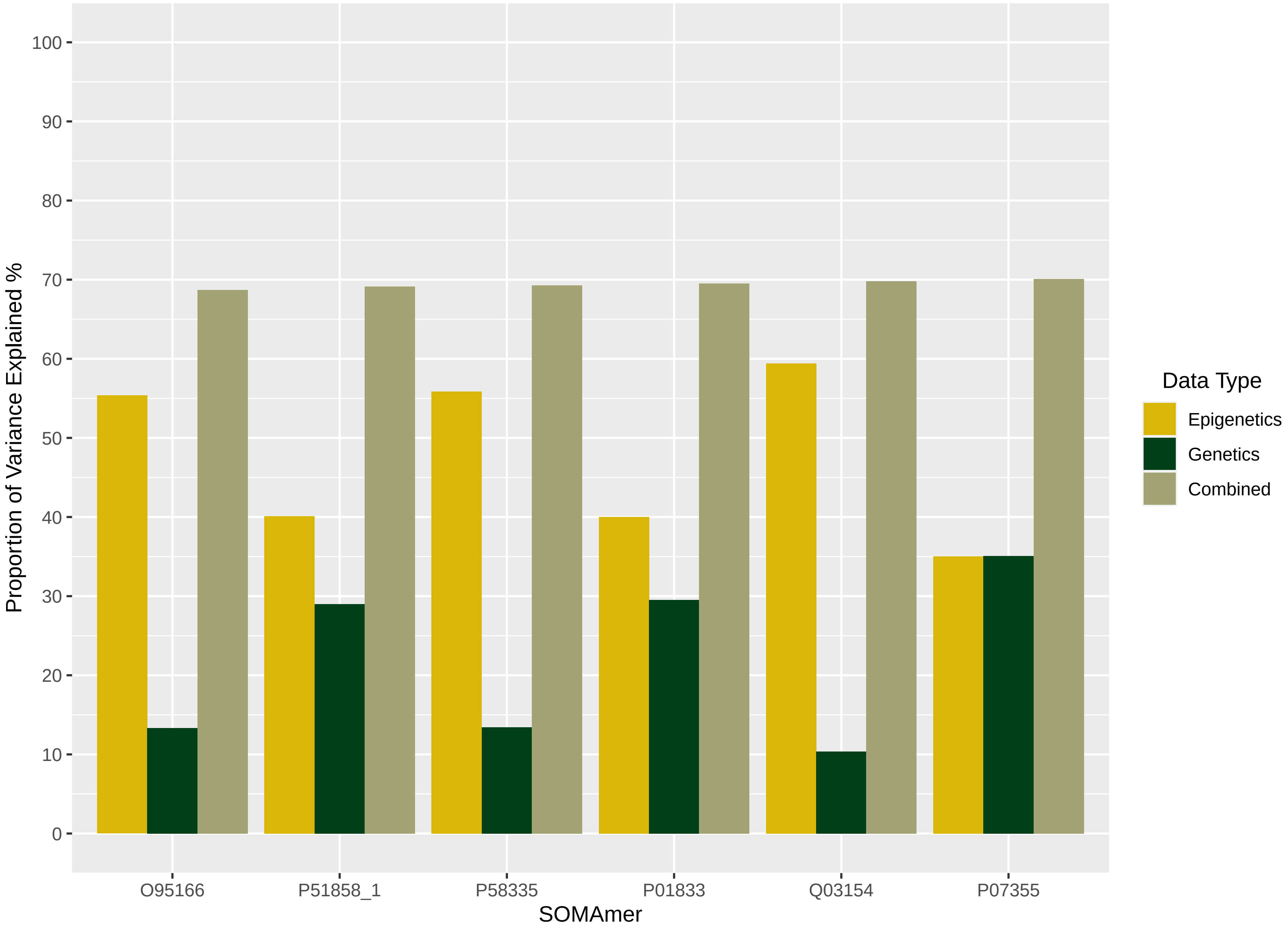

Proportion of Variance Explained %
100
90
80
70
60
50
40
30
20
10
0
O95166
P51858_1
P58335
P01833
Q03154
P07355
SOMAmer
Data Type
Epigenetics
Genetics
Combined

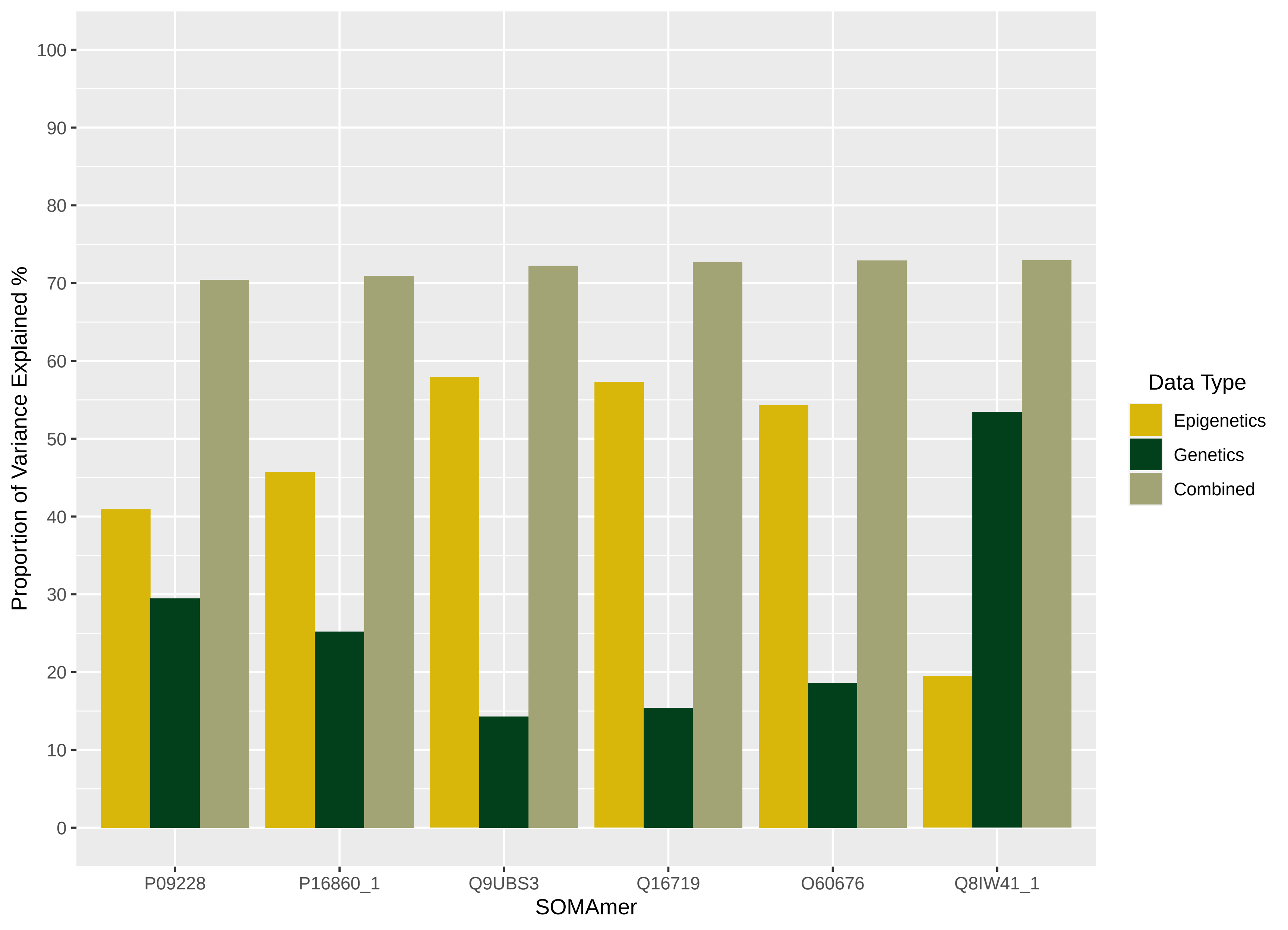

Proportion of Variance Explained %
100
90
80
70
60
50
40
30
20
10
0
P09228
P16860_1
Q9UBS3
Q16719
O60676
Q8IW41_1
SOMAmer
Data Type
Epigenetics
Genetics
Combined

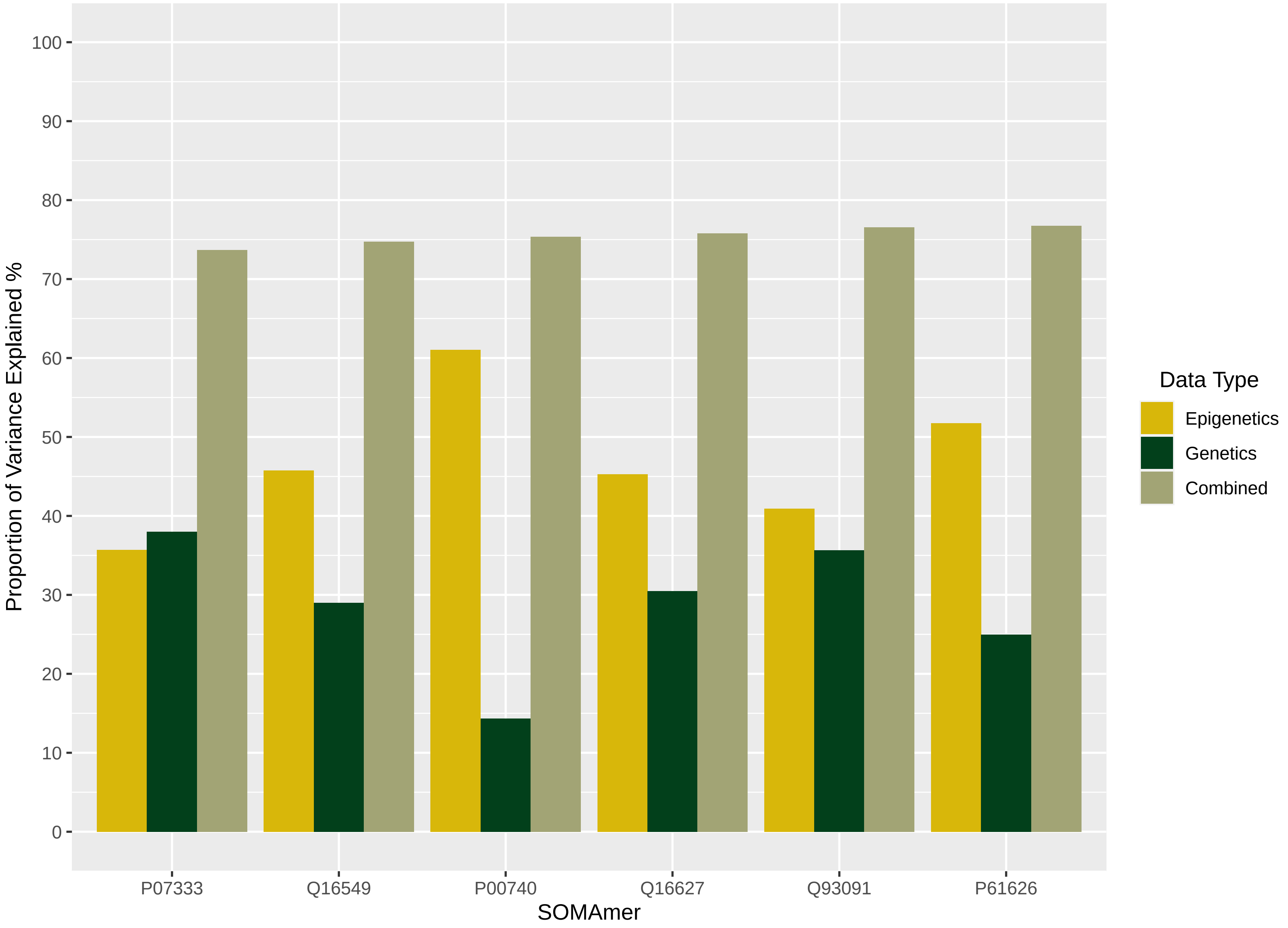
Proportion of Variance Explained %
0
10
20
30
40
50
60
70
80
90
100
P07333
Q16549
P00740
Q16627
Q93091
P61626
SOMAmer
Data Type
Epigenetics
Genetics
Combined

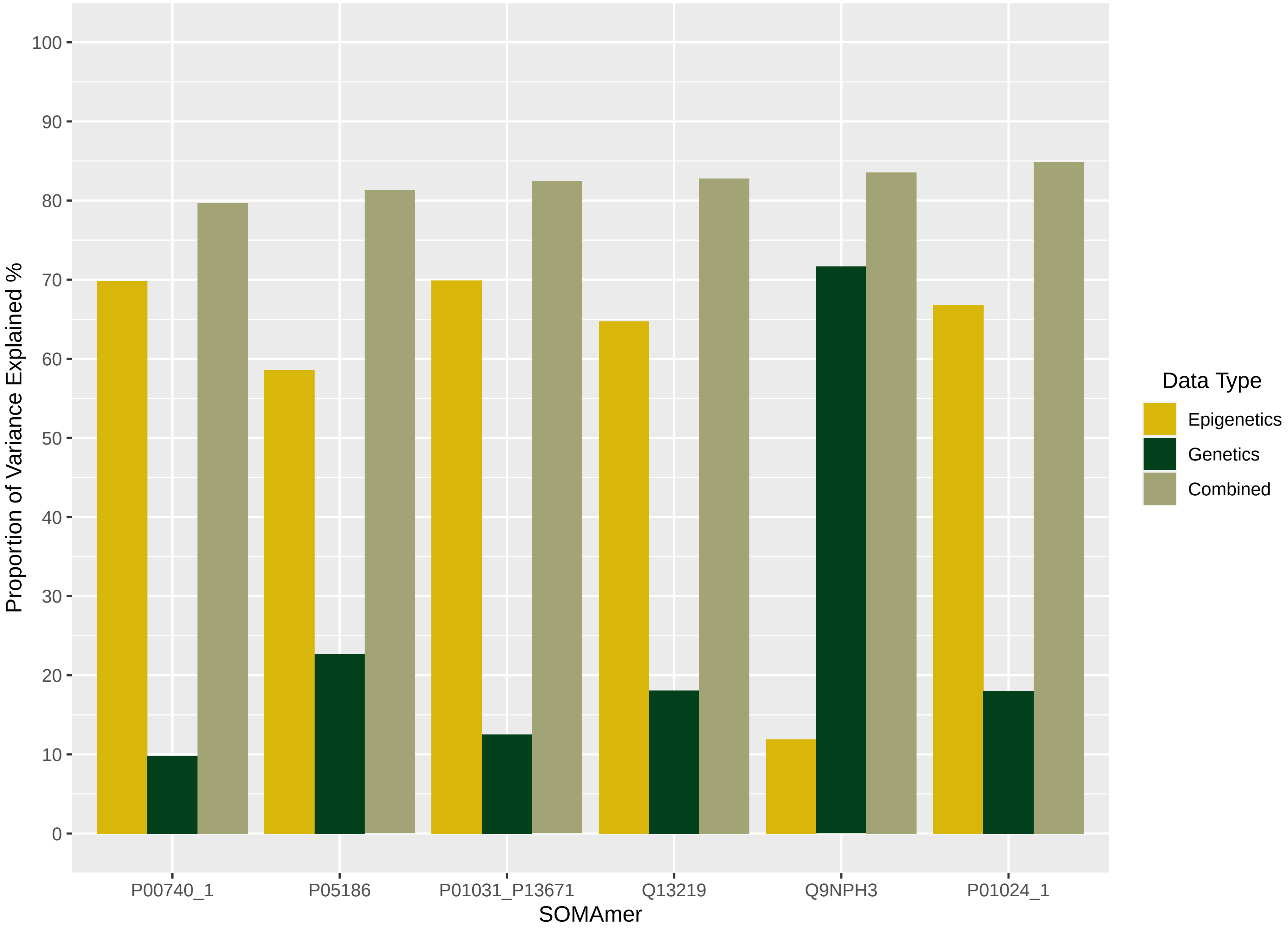

Proportion of Variance Explained %
100
90
80
70
60
50
40
30
20
10
0
P00740_1
P05186
P01031_P13671
Q13219
Q9NPH3
P01024_1
SOMAmer
Data Type
Epigenetics
Genetics
Combined

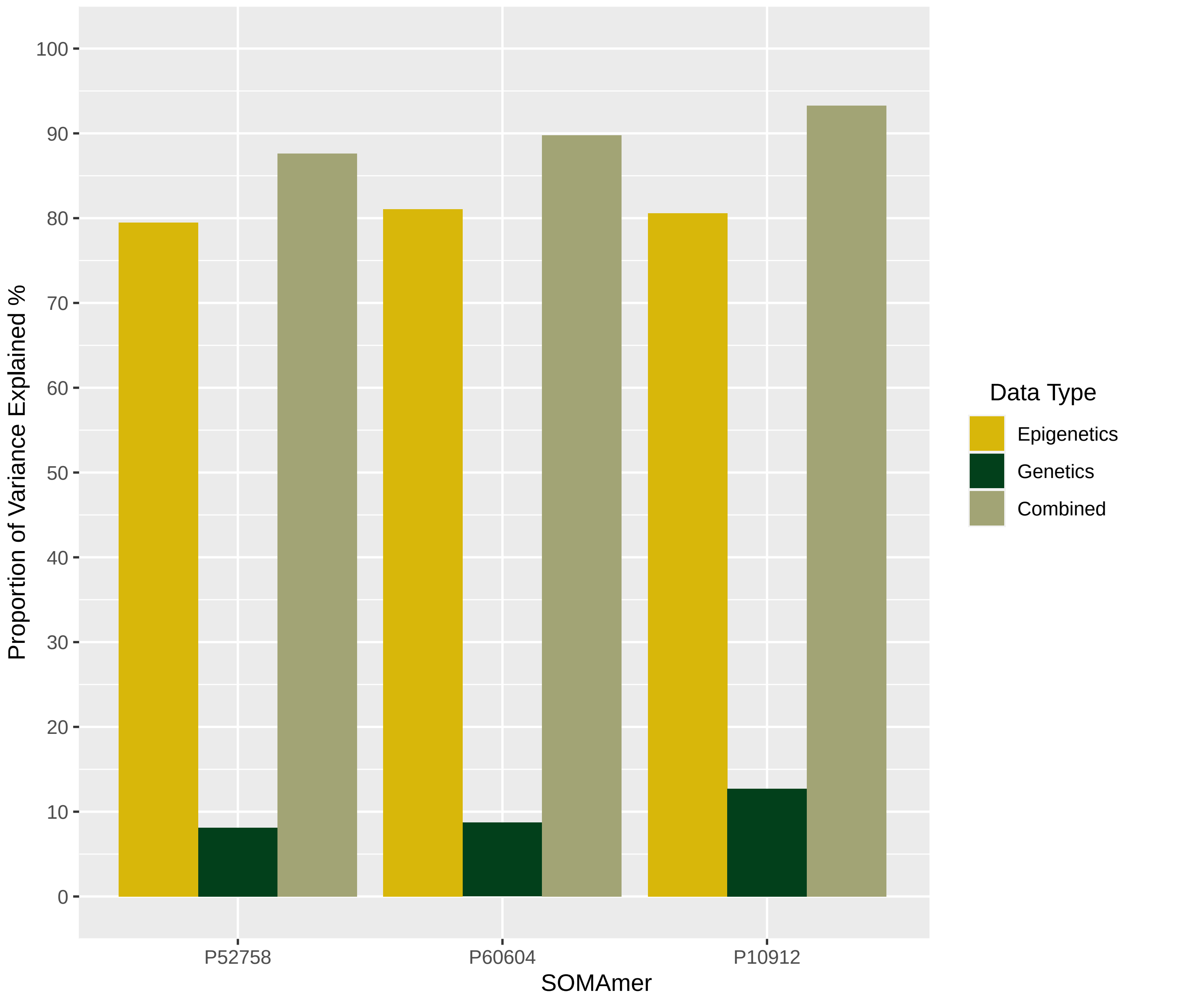

Proportion of Variance Explained %
100
90
80
70
60
50
40
30
20
10
0
P52758
P60604
P10912
SOMAmer
Data Type
Epigenetics
Genetics
Combined
